# The Diurnal Rhythmicity of Untargeted Metabolomic Profiles in Humans

**DOI:** 10.64898/2026.04.28.26347027

**Authors:** Pavithra Nagarajan, Tariq Faquih, Bing Yu, Hassan S. Dashti, Raymond Noordam, Qian Xiao, Robert Kaplan, Qibin Qi, Carmen R. Isasi, Richa Saxena, Frank A.J.L. Scheer, Susan Redline, Tamar Sofer, Heming Wang

## Abstract

It is well established that the circadian rhythm system plays a crucial role in metabolic regulation. Yet, the influence of time of sample collection in cross-sectional, untargeted metabolomics datasets remains underexplored. We therefore investigated the temporal oscillations of 853 serum metabolites from 5954 fasting samples in the Hispanic Community Health Study / Study of Latinos cohort, where blood samples were drawn within a short daytime window (7:45 am - 1:38 pm). In total, 84 serum metabolites were significantly associated (p<5.9e-05) with blood draw time. Time-varying metabolites exhibit enrichment in pathways of incretin hormones and light signaling, mechanistic connections to neurological and circadian control (dopamine response, melatonin activity), and implications for therapeutics (serotonergic drugs) and health disorder understanding (sleep, depression). Unsupervised analysis revealed latent, nonlinear patterns in five metabolite clusters according to their temporal profiles. Impact on false and missed discovery, and altered effect size estimates for metabolite-health disorder associations underscore the importance of accounting for time. Results fundamentally show that serum metabolites exhibit non-negligible temporal variation even in a restricted short daytime window. Accounting for this variance appears important for precise disease understanding from metabolomics data.

## INTRODUCTION

Metabolites are a captivating data modality for investigating the health state. Offering a granular view close to the phenome, these small molecules lie at the crossroads of lifestyle, environmental exposures, and internal physiology. Investigating the associations between metabolites and health disorders carries strength of interpretability due to their direct participation in biological pathways. With the advent of untargeted metabolomics technology, there has been a rapid rise of novel metabolite discoveries for health areas ranging from sleep timing, diabetes, to depression ^1-3^.

However, a key challenge metabolites present is their temporal fluctuations, which can be driven centrally by the endogenous circadian clock (orchestrated by the suprachiasmatic nucleus), or external factors including time of food intake, light exposure, sleep-wake patterns, and social and physical activities. For example, melatonin rises in the evening and peaks during the night, whereas cortisol peaks shortly after awakening and declines progressively throughout the day. Furthermore, recent small-scale experimental studies under tightly-controlled conditions have uncovered additional time-varying metabolites, critically broadening perspective on the human rhythmic metabolome^4^. However, such experimental studies may not reflect real-world conditions applicable to the clinical or epidemiological analysis contexts^5,6^. In contrast, large-scale cross-sectional studies enable powerful statistical analyses of metabolite associations with health disorders, yet often do not consider the influence of time-of-day variation on metabolite quantities^1,3,7,8^. Although blood sample collection in cohorts is typically confined to a specific timeframe (e.g. daytime-afternoon), time-of-day variation can still have a nonnegligible impact on drawn conclusions. For example, morning fasting glucose levels differ substantially from afternoon levels, which can lead to incorrectly missed diagnoses of diabetes if failing to account for time of measurement ^9^.

Taken together, variation in time of blood sample collection may influence metabolite biomarker discovery and obscure disease mechanism conclusions. This is important to systematically evaluate in a metabolome-wide manner, as untargeted metabolomics technology is driving the current trend of investigating hundreds of metabolites’ profiles for health outcomes in cross-sectional frameworks ^1-3,7,8,10-12^. Prior studies have examined the impact of blood sample collection time on metabolite profiles in population cohorts, but with considerably small sample sizes (N<500) resulting in varying conclusions ^13-15^. Thus, for cross-sectional data relevant for large-scale metabolome-wide analyses, the impact of time-of-day variation on metabolites appears underexplored.

We therefore sought to investigate in a population-based cohort study with close to 6,000 samples, whether serum metabolites measured from fasting blood samples drawn across a short six-hour daytime window, exhibit temporal trends important to account for. We envisioned this analysis will help reveal the impact of diurnal rhythmicity, informing future considerations in metabolomics methodological design.

## RESULTS

### Serum Metabolites Exhibit Associations with Time of Blood Draw

Sample draw times ranged across 7:45 am to 1:38 pm and were from 5954 fasting samples from the Hispanic Community Health Study/Study of Latinos (HCHS/SOL) cohort (Supplementary Figure 1, Supplementary Table 1). We conducted metabolome-wide association analyses of blood draw time (transformed into sine and cosine components), adjusting for age, sex, sleep and wake timing, as well as study population and design variables. Analyses were performed separately in two batches and then combined using random-effects meta-analysis (Methods). In total, 52 out of the 853 serum metabolites were revealed to exhibit significant (p<0.05/853) association with time of blood draw, in the 1 degree of freedom (df) sine test, 1df cosine test, or 2df joint test simultaneously assessing sine and cosine effects (Figure 1a, Table 1). 23 of these 52 metabolites have been previously reported to be rhythmic in blood (Table 1). The majority class of the identified 52 metabolites were lipids – all exhibiting a significant negative association with sine time of blood draw, or a daytime rise pattern (Supplementary Figure 2).

**Table 1.** 52 Blood Draw Time-Associated Serum Metabolites Identified in Primary Analysis. Effect size, Bonferroni-corrected p-value, and standard error estimates for 52 serum metabolites significantly associated (Bonferroni-corrected P < 0.05) with sine (P_SIN_, B_SIN_, SE_SIN_), cosine measures (P_COS_, B_COS_, SE_COS_), or the joint sine/cosine measure (P_JOINT_) of time of blood draw in the primary model analysis are displayed. Bolded p-values denote passing significance. Uncharacterized metabolites are reported via the chemical identifier reported by the Metabolon Discovery HD4 platform with a prefix of X-(e.g. X-18921). The superscript^**†**^ denotes that a given metabolite exhibits significant association with time of blood draw across all three sensitivity models (Supplementary Table 2).

| Metabolite | Prior Reported Evidence of Diurnal Rhythmicity in Blood (PMID(s)) | Metabolite Category | $P_{\text{sin}}$ | $\text{Effect}_{\text{sin}}$ | $SE_{\text{sin}}$ | $P_{\text{cos}}$ | $\text{Effect}_{\text{cos}}$ | $SE_{\text{cos}}$ | $P_{\text{joint}}$ |
| --- | --- | --- | --- | --- | --- | --- | --- | --- | --- |
| leucylglycine<sup>†</sup> |  | Peptide | 5.55E-01 | 0.78 | 0.23 | **1.02E-03** | -2.15 | 0.44 | **1.94E-05** |
| succinoyltaurine |  | Amino Acid | **4.86E-02** | -0.81 | 0.2 | 1 | 0.41 | 0.43 | 1.66E-01 |
| (R)-3-hydroxybutyrylcarnitine<sup>†</sup> |  | Lipid | **2.41E-02** | -0.88 | 0.21 | 1 | 0.35 | 0.45 | 9.79E-02 |
| hexanoylcarnitine (C6) | 22823870, 34069741 | Lipid | **3.60E-02** | -0.75 | 0.18 | 1 | 0.4 | 0.42 | 1.23E-01 |
| myristoleoylcarnitine (C14:1)<sup>†</sup> | 30293576, 34069741 | Lipid | **1.09E-02** | -0.86 | 0.2 | 1 | 0.28 | 0.43 | 5.03E-02 |
| hexanoylglutamine<sup>†</sup> | 30293576, 34069741 | Lipid | **1.19E-05** | -1.13 | 0.2 | 1 | 0.51 | 0.47 | **4.82E-05** |
| trans-2-hexenoylglycine<sup>†</sup> |  | Lipid | **2.02E-02** | -0.90 | 0.21 | 1 | 0.32 | 0.48 | 8.93E-02 |
| dodecanedioate (C12-DC)<sup>†</sup> | 30293576, 34069741 | Lipid | **1.71E-04** | -1.00 | 0.19 | 1 | 0.6 | 0.43 | **4.28E-04** |
| sebacate (C10-DC)<sup>†</sup> | 30293576, 34069741 | Lipid | **9.66E-03** | -0.93 | 0.21 | 1 | 0.8 | 0.43 | **9.55E-03** |
| tetradecanedioate (C14-DC)<sup>†</sup> |  | Lipid | **1.41E-02** | -0.83 | 0.19 | 1 | -0.25 | 0.4 | 6.53E-02 |
| hexadecanedioate (C16-DC)<sup>†</sup> | 30293576, 34069741 | Lipid | **2.26E-03** | -0.85 | 0.18 | 1 | -0.01 | 0.4 | **1.38E-02** |
| dodecenedioate (C12:1-DC)<sup>†</sup> |  | Lipid | **2.77E-11** | -1.37 | 0.18 | 1 | 0.97 | 0.42 | **1.84E-11** |
| dodecadienoate (12:2)<sup>†</sup> |  | Lipid | **9.85E-06** | -1.08 | 0.19 | 1 | 0.7 | 0.4 | **1.65E-05** |
| tetradecadienedioate (C14:2-DC)<sup>†</sup> |  | Lipid | **5.72E-07** | -1.08 | 0.17 | 1 | 0.48 | 0.41 | **2.32E-06** |
| decadienedioic acid (C10:2-DC)<sup>†</sup> |  | Lipid | **2.77E-05** | -0.98 | 0.18 | 1 | 1.19 | 0.39 | **2.01E-06** |
| 3-hydroxyoctanoate<sup>†</sup> | 30293576, 34069741 | Lipid | **2.88E-05** | -1.02 | 0.19 | 1 | 1.22 | 0.43 | **3.83E-06** |
| 3-hydroxydecanoate<sup>†</sup> | 22308371, 34069741 | Lipid | **1.60E-06** | -1.12 | 0.19 | 1 | 0.85 | 0.4 | **1.31E-06** |
| 3-hydroxylaurate<sup>†</sup> | 30293576, 34069741 | Lipid | **1.51E-04** | -0.97 | 0.19 | 1 | 0.38 | 0.4 | **6.52E-04** |
| 3-hydroxysebacate<sup>†</sup> |  | Lipid | **2.64E-06** | -1.23 | 0.21 | 1 | 1.35 | 0.46 | **2.83E-07** |
| 16-hydroxypalmitate<sup>†</sup> |  | Lipid | **2.14E-02** | -0.79 | 0.19 | 1 | -0.48 | 0.41 | 5.84E-02 |
| 3-hydroxyhexanoate<sup>†</sup> | 30293576, 34069741 | Lipid | **1.74E-04** | -1.01 | 0.19 | 1 | 0.87 | 0.43 | **1.48E-04** |
| glucose | 28744188, 24475090, 30293576,<br>31211770, 34069741 | Carbohydrate | **4.24E-02** | 0.76 | 0.19 | 1 | -0.56 | 0.42 | 9.48E-02 |
| 3-hydroxybutyrate (BHBA)<sup>†</sup> | 29522222, 34069741 | Lipid | **3.81E-06** | -1.21 | 0.21 | 1 | 0.39 | 0.45 | **1.97E-05** |
| 10-nonadecenoate (19:1n9)<sup>†</sup> |  | Lipid | **3.10E-05** | -0.90 | 0.16 | 1 | -0.24 | 0.36 | **1.77E-04** |
| 10-heptadecenoate (17:1n7)<sup>†</sup> | 22308371, 34069741 | Lipid | **1.43E-03** | -0.75 | 0.16 | 1 | -0.51 | 0.37 | **3.35E-03** |
| eicosenoate (20:1)<sup>†</sup> |  | Lipid | **8.07E-05** | -0.91 | 0.17 | 1 | -0.03 | 0.37 | **5.56E-04** |
| oleate/vaccenate (18:1)<sup>†</sup> |  | Lipid | **1.13E-05** | -0.86 | 0.15 | 1 | -0.28 | 0.35 | **5.98E-05** |
| erucate (22:1n9)<sup>†</sup> |  | Lipid | **1.60E-02** | -0.83 | 0.19 | 1 | 0.35 | 0.43 | 6.46E-02 |
| palmitoleate (16:1n7) | 22308371, 34069741 | Lipid | **1.42E-02** | -0.65 | 0.15 | 1 | -0.58 | 0.36 | **2.19E-02** |
| docosahexaenoate (DHA; 22:6n3) | 22308371, 34069741 | Lipid | **1.83E-03** | -0.81 | 0.17 | 1 | 0.26 | 0.36 | **8.71E-03** |
| docosapentaenoate (n3 DPA;<br>22:5n3)<sup>†</sup> | 22308371, 34069741 | Lipid | **4.23E-07** | -0.96 | 0.16 | 1 | 0.07 | 0.35 | **3.31E-06** |
| docosadienoate (22:2n6)<sup>†</sup> | 22308371, 34069741 | Lipid | **6.84E-05** | -0.87 | 0.16 | 1 | -0.2 | 0.35 | **4.03E-04** |
| stearidonate (18:4n3)<sup>†</sup> | 22308371 | Lipid | **2.38E-05** | -0.87 | 0.16 | 1 | 0.3 | 0.38 | **1.26E-04** |
| linolenate [alpha or gamma; (18:3n3<br>or 6)]<sup>†</sup> | 22308371, 34069741 | Lipid | **2.69E-05** | -0.97 | 0.17 | 1 | -0.03 | 0.37 | **1.92E-04** |
| dihomo-linolenate (20:3n3 or n6)<sup>†</sup> | 22308371, 34069741 | Lipid | **1.68E-03** | -0.82 | 0.17 | 1 | -0.06 | 0.37 | **1.03E-02** |
| hexadecadienoate (16:2n6)<sup>†</sup> |  | Lipid | **4.34E-05** | -0.92 | 0.17 | 1 | 0.06 | 0.37 | **3.02E-04** |
| tetradecadienoate (14:2)<sup>†</sup> |  | Lipid | **7.89E-07** | -1.13 | 0.18 | 1 | 0.6 | 0.39 | **1.92E-06** |
| dihomo-linoleate (20:2n6)<sup>†</sup> | 22308371, 34069741 | Lipid | **1.21E-05** | -1.07 | 0.19 | 1 | 0.13 | 0.39 | **8.37E-05** |
| linoleate (18:2n6)<sup>†</sup> | 22308371, 34069741 | Lipid | **5.06E-06** | -0.94 | 0.16 | 1 | -0.19 | 0.34 | **3.26E-05** |
| palmitate (16:0)<sup>†</sup> | 22308371, 34069741 | Lipid | **2.32E-05** | -0.82 | 0.15 | 1 | -0.44 | 0.34 | **7.27E-05** |
| stearate (18:0)<sup>†</sup> |  | Lipid | **1.33E-05** | -0.92 | 0.16 | 1 | -0.09 | 0.37 | **9.41E-05** |
| margarate (17:0)<sup>†</sup> |  | Lipid | **7.04E-06** | -0.96 | 0.17 | 1 | -0.1 | 0.37 | **5.06E-05** |
| 10-undecenoate (11:1n1) |  | Lipid | **1.06E-02** | -0.78 | 0.18 | 1 | -0.01 | 0.44 | 6.08E-02 |
| 5-dodecenoate (12:1n7)<sup>†</sup> |  | Lipid | **5.68E-05** | -0.91 | 0.17 | 1 | 0.24 | 0.37 | **3.23E-04** |
| cis-4-decenoate (10:1n6)<sup>†</sup> |  | Lipid | **3.83E-06** | -1.07 | 0.18 | 1 | 0.82 | 0.41 | **3.71E-06** |
| (2 or 3)-decenoate (10:1n7 or n8)<sup>†</sup> |  | Lipid | **3.27E-04** | -0.97 | 0.19 | 1 | 0.23 | 0.41 | **1.84E-03** |
| glutamine conjugate of C7H12O2<sup>†</sup> |  | Partially Characterized Molecules | **1.19E-02** | -0.89 | 0.21 | 1 | 0.16 | 0.48 | 6.39E-02 |
| glutamine conjugate of C6H10O2 (2)<sup>†</sup> |  | Partially Characterized Molecules | **6.26E-04** | -0.89 | 0.18 | 1 | 0.13 | 0.41 | **3.83E-03** |
| branched-chain, straight-chain, or cyclopropyl 12:1 fatty acid<sup>†</sup> |  | Partially Characterized Molecules | **6.99E-08** | -1.15 | 0.18 | 1 | 0.6 | 0.38 | **1.72E-07** |
| X-18921<sup>†</sup> |  | Uncharacterized Molecules | **2.62E-05** | -0.98 | 0.18 | 1 | 0.86 | 0.42 | **2.33E-05** |
| X-21353<sup>†</sup> |  | Uncharacterized Molecules | **7.51E-05** | -1.12 | 0.21 | 1 | 0.58 | 0.43 | **2.14E-04** |
| X-21607<sup>†</sup> |  | Uncharacterized Molecules | **6.77E-03** | -0.91 | 0.2 | 1 | 1.19 | 0.43 | **8.88E-04** |

**Figure 1.**
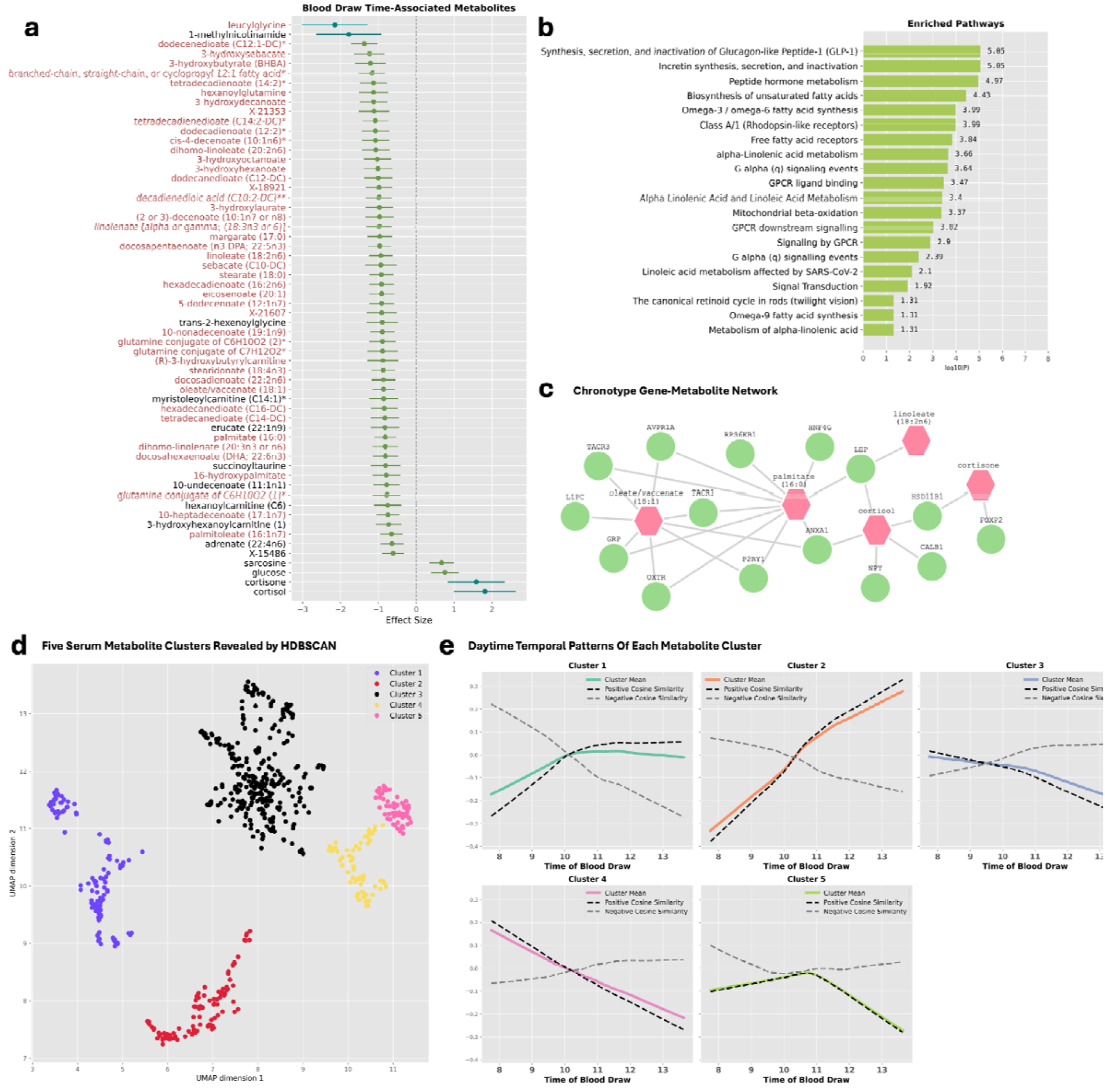
Serum Metabolites Exhibit Significant Temporal Variation. **a**. Forest plot of 60 blood-draw time associated serum metabolites identified in the primary combined sex models. In blue, is the cosine time of blood draw-associated metabolites and in green the sine time of blood-draw associated metabolites. Metabolites that exhibit significance in the 2df joint test (which jointly assesses both sine and cosine effects of time) are in red font. **b**. Pathway enrichment analysis results (false discovery rate < 0.05) for the complete set of 84 time of blood draw-associated serum metabolites discovered across combined samples, sex or age-stratified analyses, and/or primary and sensitivity models. **c**. Chronotype gene-metabolite connections identified by network analysis across all 84 blood draw-time associated metabolites. Pink hexagons depict serum metabolites, and green circles depict chronotype genes. **d**. Scatter plot of 672 serum metabolites in five clusters identified by HDBSCAN in UMAP dimensions. **e**. Cluster average LOWESS trends across time of blood draw. The overlaid dashed black line represents the average trajectory of metabolites with positive cosine similarity to the cluster mean and a dashed gray line for those with negative cosine similarity.

One modified amino acid (succinoyltaurine) and six uncharacterized molecules also exhibited a significant negative association with sine time of blood draw. The carbohydrate glucose uniquely showed a significant positive association with sine time of blood draw (a morning to afternoon dip; Supplementary Figure 2). Only one metabolite – the dipeptide leucylglycine – exhibited a significant negative association with cosine time of blood draw (a convex morning rise and afternoon dip; Supplementary Figure 2).

Three sensitivity analysis models sequentially adjusting for time of last food intake prior to fasting (capturing fasting duration), lifestyle behaviors (substance use, physical activity, diet quality), and medication use for underlying health disorders (Methods), identified 46 of the 52 metabolites to remain robustly associated (p<0.05/853) with time of blood draw across all three sensitivity models (Table 1, Supplementary Table 2). The fully-adjusted sensitivity model revealed known circadian metabolites cortisol, cortisone, and additionally six other metabolites to be significantly associated with time of blood draw (Supplementary Tables 2-3).

In the external validation analysis using independent samples from the SHIFT study (https://clinicaltrials.gov/study/NCT02997319; N=82 fasting samples; Methods, Supplementary Table 1), 52 out of the total identified 60 blood-draw time associated metabolites were available. Of these, 27 serum metabolites exhibited significant (p<0.05/52) differences in quantified levels between morning and evening sample collection times (Supplementary Table 4). Amongst these 27 validated metabolites, all metabolites with significantly higher evening levels in SHIFT (n=24), consistently exhibited a significant negative sine effect (morning to afternoon rise) in the discovery (HCHS/SOL) dataset. Similarly, all metabolites identified to have significantly lower evening levels in SHIFT (n=3), were consistently significantly associated with cosine time of blood draw (convex rise-fall, or concave fall-rise daytime pattern) in the discovery dataset.

### Age and Sex-Stratified Analyses Reveal Additional Time-Associated Metabolites

Female-specific analysis identified nine additional time-varying serum metabolites: one bile acid, three acyl carnitines, one vitamin B3 derivative, one branched fatty acid, one sterol, one dicarboxylic acid, and an uncharacterized molecule (Table 2, Supplementary Table 5). Male-specific analysis uniquely identified nine time-varying serum metabolites: cotinine, five diacylglycerols, two phosphatidylinositols, and one modified amino acid (Table 2, Supplementary Table 6). In younger adults (age<50 years) (S)-3-hydroxybutyrylcarnitine was uniquely revealed to be time-varying (Table 2, Supplementary Table 7). In older adults (aged >=50 years) five time-varying metabolites were uniquely revealed: two acylcholines, o-cresol sulfate, and two modified amino acids (Table 2, Supplementary Table 8).

**Table 2.**
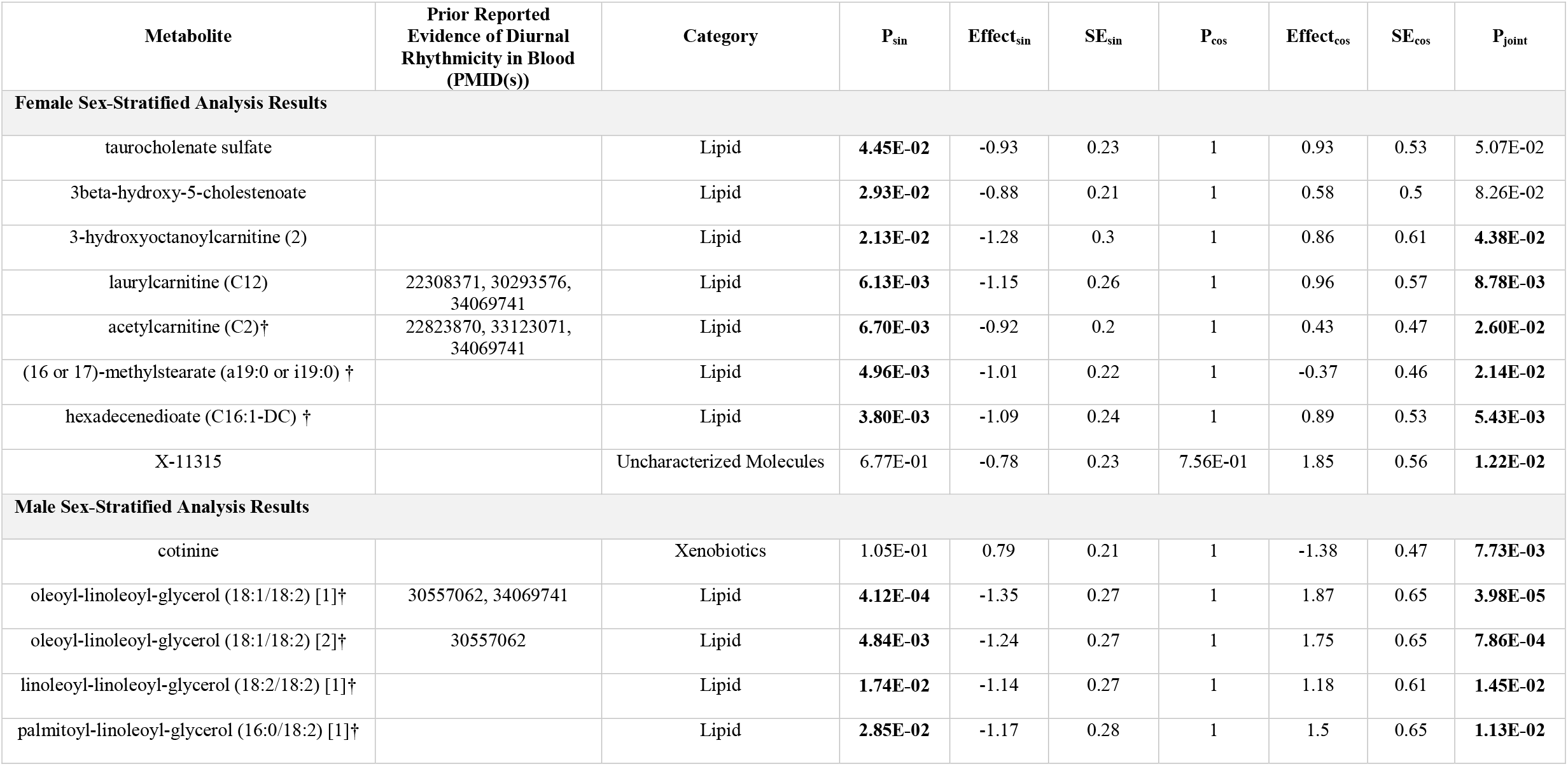
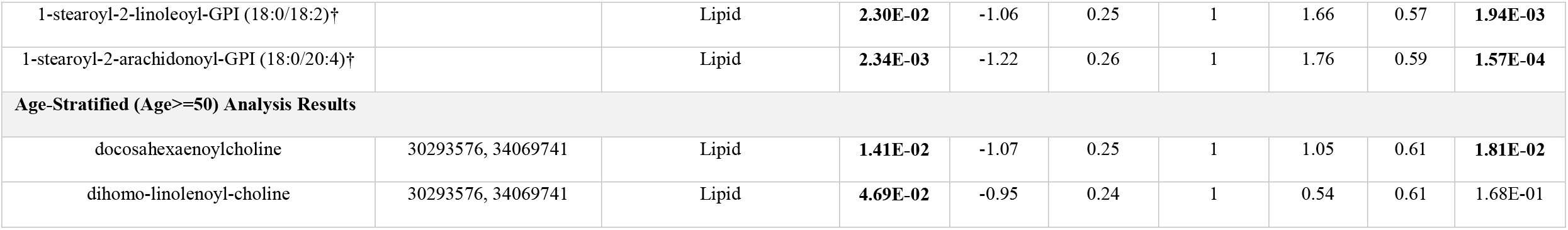
Sex-Specific or Age-Specific Blood-Draw Time-Associated Serum Metabolites. Effect size, Bonferroni-corrected p-value, and standard error estimates for 17 serum metabolites significantly (Bonferroni-corrected P < 0.05) associated with sine (P_SIN_, B_SIN_, SE_SIN_), cosine measures (P_COS_, B_COS_, SE_COS_), or the joint sine/cosine measure (P_JOINT_) of time of blood draw in primary model analyses of particular sex or age-specific groups are displayed. Bolded p-values denote passing significance. All metabolites shown here are those uniquely identified in these stratified analysis (not identified by non-stratified analyses). Uncharacterized metabolites are reported via the chemical identifier reported by the Metabolon Discovery HD4 platform with a prefix of X-(e.g. X-11315). The superscript† denotes that a given metabolite exhibits significant association with time of blood draw across all three sensitivity models (Supplementary Tables 5-8). Heterogeneity p-values by sex and age are provided in Supplementary Table 10.

| Metabolite | Prior Reported Evidence of Diurnal Rhythmicity in Blood (PMID(s)) | Category | $P_{\text{sin}}$ | $\text{Effect}_{\text{sin}}$ | $SE_{\text{sin}}$ | $P_{\text{cos}}$ | $\text{Effect}_{\text{cos}}$ | $SE_{\text{cos}}$ | $P_{\text{joint}}$ |
| --- | --- | --- | --- | --- | --- | --- | --- | --- | --- |
| **Female Sex-Stratified Analysis Results** |  |  |  |  |  |  |  |  |  |
| taurochenolate sulfate |  | Lipid | **4.45E-02** | -0.93 | 0.23 | 1 | 0.93 | 0.53 | 5.07E-02 |
| 3beta-hydroxy-5-cholestenoate |  | Lipid | **2.93E-02** | -0.88 | 0.21 | 1 | 0.58 | 0.5 | 8.26E-02 |
| 3-hydroxyoctanoylcarnitine (2) |  | Lipid | **2.13E-02** | -1.28 | 0.3 | 1 | 0.86 | 0.61 | **4.38E-02** |
| laurylcarnitine (C12) | 22308371, 30293576, 34069741 | Lipid | **6.13E-03** | -1.15 | 0.26 | 1 | 0.96 | 0.57 | **8.78E-03** |
| acetylcarnitine (C2)† | 22823870, 33123071, 34069741 | Lipid | **6.70E-03** | -0.92 | 0.2 | 1 | 0.43 | 0.47 | **2.60E-02** |
| (16 or 17)-methylstearate (a19:0 or i19:0) † |  | Lipid | **4.96E-03** | -1.01 | 0.22 | 1 | -0.37 | 0.46 | **2.14E-02** |
| hexadecenedioate (C16:1-DC) † |  | Lipid | **3.80E-03** | -1.09 | 0.24 | 1 | 0.89 | 0.53 | **5.43E-03** |
| X-11315 |  | Uncharacterized Molecules | 6.77E-01 | -0.78 | 0.23 | 7.56E-01 | 1.85 | 0.56 | **1.22E-02** |
| **Male Sex-Stratified Analysis Results** |  |  |  |  |  |  |  |  |  |
| cotinine |  | Xenobiotics | 1.05E-01 | 0.79 | 0.21 | 1 | -1.38 | 0.47 | **7.73E-03** |
| oleoyl-linoleoyl-glycerol (18:1/18:2) [1]† | 30557062, 34069741 | Lipid | **4.12E-04** | -1.35 | 0.27 | 1 | 1.87 | 0.65 | **3.98E-05** |
| oleoyl-linoleoyl-glycerol (18:1/18:2) [2]† | 30557062 | Lipid | **4.84E-03** | -1.24 | 0.27 | 1 | 1.75 | 0.65 | **7.86E-04** |
| linoleoyl-linoleoyl-glycerol (18:2/18:2) [1]† |  | Lipid | **1.74E-02** | -1.14 | 0.27 | 1 | 1.18 | 0.61 | **1.45E-02** |
| palmitoyl-linoleoyl-glycerol (16:0/18:2) [1]† |  | Lipid | **2.85E-02** | -1.17 | 0.28 | 1 | 1.5 | 0.65 | **1.13E-02** |
| 1-stearoyl-2-linoleoyl-GPI (18:0/18:2)† |  | Lipid | **2.30E-02** | -1.06 | 0.25 | 1 | 1.66 | 0.57 | **1.94E-03** |
| 1-stearoyl-2-arachidonoyl-GPI (18:0/20:4)† |  | Lipid | **2.34E-03** | -1.22 | 0.26 | 1 | 1.76 | 0.59 | **1.57E-04** |
| **Age-Stratified (Age>=50) Analysis Results** |  |  |  |  |  |  |  |  |  |
| docosahexaenoylcholine | 30293576, 34069741 | Lipid | **1.41E-02** | -1.07 | 0.25 | 1 | 1.05 | 0.61 | **1.81E-02** |
| dihomo-linolenoyl-choline | 30293576, 34069741 | Lipid | **4.69E-02** | -0.95 | 0.24 | 1 | 0.54 | 0.61 | 1.68E-01 |

This amounted to across all pooled and stratified analyses, a total of 84 serum metabolites identified to be significantly associated with time of blood draw, showing consistent directions of effect across metabolite profiling batches (Supplementary Table 9, Supplementary Figure 3). Of these, seven metabolites appear to have strong evidence (P<0.05/84) of sex or age-based heterogeneous effects: 3-hydroxylaurate, eicosenoate, hexanoylglutamine, oleoyl-linoleoyl-glycerol, leucylglycine, (R)-3-hydroxybutyrylcarnitine, and glutamine conjugate of C7H12O2 (Supplementary Table 10).

### Pathway Enrichment and Network Analyses of Time-Associated Metabolites

For the total set of 84 time-varying metabolites, pathway enrichment analysis revealed 20 significantly enriched (false discovery rate < 0.05) pathways – the top two enriched being *Synthesis, secretion, and inactivation of Glucagon-like Peptide-1 (GLP-1)* (FDR: 9.0e-06), and *Incretin synthesis, secretion, and inactivation* (FDR: 9.0e-06) (Figure 1b, Supplementary Table 11). One enriched pathway was also noted in visual light cue processing: *The canonical retinoid cycle in rods* (twilight vision) (FDR: 4.9e-02).

60 metabolites exhibited absolute partial correlation coefficient values >=0.3 (Supplementary Tables 12-13). Joint integrative network analysis between chronotype genes (Supplementary Table 14) and the 84 time-varying metabolites revealed cortisol, cortisone, oleic acid, palmitic acid, and linoleic acid to be connected to one or multiple of 15 chronotype genes (Figure 1c, Supplementary Table 15) ^17^. The KEGG pathway *Biosynthesis of unsaturated fatty acids* was enriched jointly by chronotype gene *HSD17B12* and eight serum lipids (FDR: 5.3e-06; Supplementary Table 16). Network analysis results are visualized at https://ggm-v0fm.onrender.com/.

### Latent Temporal Patterns Identified by Unsupervised Clustering Analysis

To investigate whether latent, nonlinear temporal patterns of serum metabolites exist, HDBSCAN clustering analysis was performed, providing as input to the algorithm the 853 serum metabolite profiles across 5954 fasting samples, with no other input features (e.g. metabolite annotations) ^16^. HDBSCAN identified five metabolite clusters (Cluster 1: 131, Cluster 2: 115, Cluster 3: 297, Cluster 4: 64, Cluster 5: 65) with a density-based clustering validation score of 0.55, indicating good separation (Figure 1d). As a validation check for algorithm choice, testing a K-means approach confirmed that a linear algorithm was not appropriate for this complex multi-dimensional data (Supplementary Figure 4). Clusters were broadly segregated according to metabolite class: Cluster 1 (phospholipids), Cluster 2 (fatty acids), Cluster 3 (food and drug-derived xenobiotics), and Clusters 4 and 5 (amino acids, peptides) (Supplementary Figure 5, Supplementary Table 17). Graphing average temporal profiles revealed trends of a rise-plateau (Cluster 1), strong rise (Cluster 2), weak-strong fall (Cluster 3), strong fall (Cluster 4), and weak rise-strong fall (Cluster 5) (Figure 1e). Investigating cosine similarity values between metabolite profiles and each cluster average revealed that each cluster also harbored metabolites with diametrically opposed (similar temporal shape, opposite orientation) trajectories with respect to the cluster average (Figure 1e, Supplementary Table 17).

### Impact of Temporal Variation on Health Trait/Disorder Biomarker Discovery

We tested associations between the 52 primary time-varying metabolites and 91 health trait/disorders, adjusting for blood draw time, to assess the clinical impact of these rhythmic metabolites (Methods). Associations were widespread, with a total of 673 significant (P<1.06e-05) associations for 58 traits across 51 serum metabolites (Figure 2a, Supplementary Tables 18-19, Supplementary Figures 6-7). We also compared effect sizes from models with blood draw time adjustment to models without, and identified the effect size ratios comparing both models for the 673 significant associations to range between 0.8 to 1.4 (Supplementary Figure 8).

**Figure 2.**
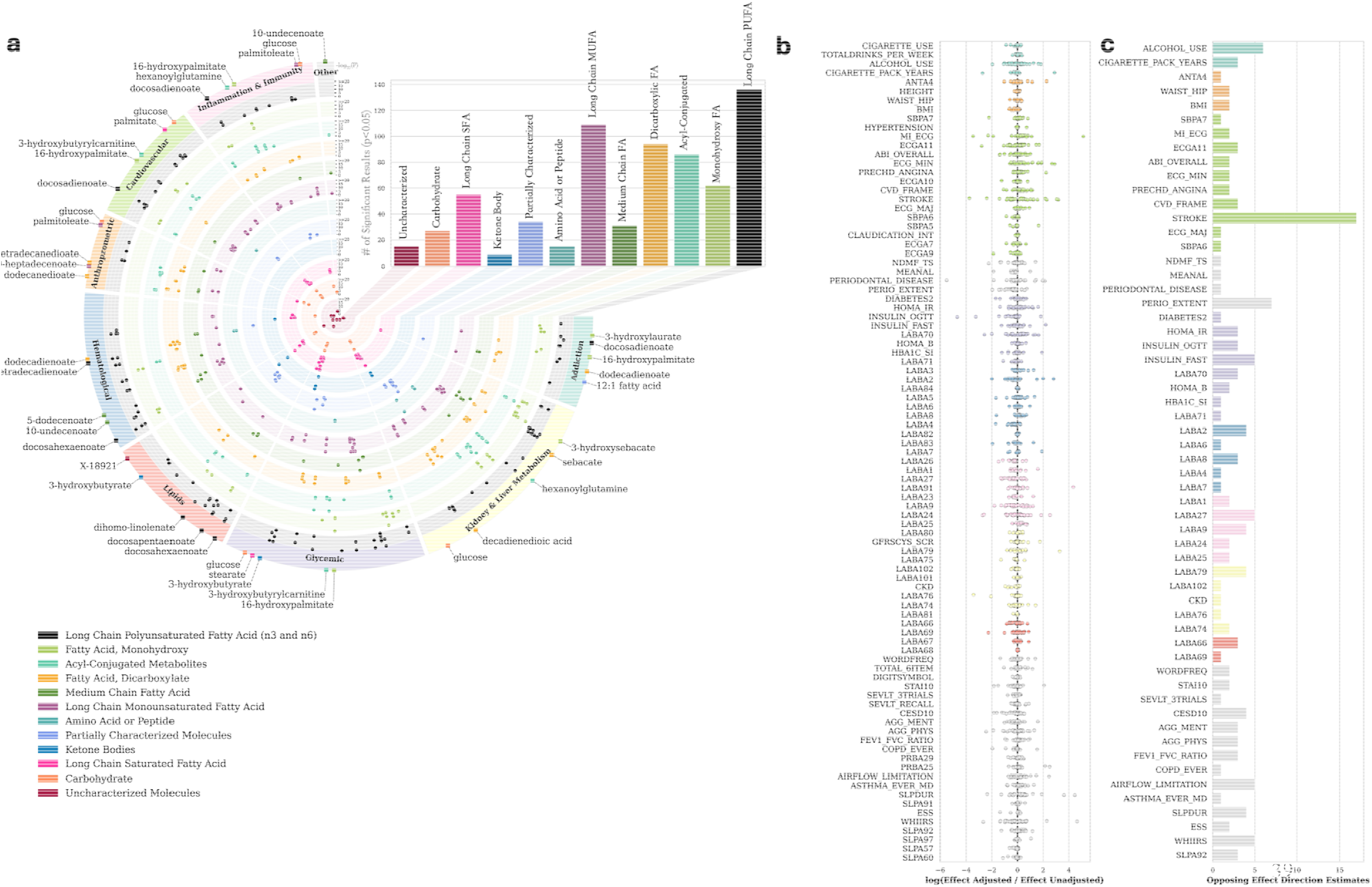
Serum Metabolite-Trait Associations and the Impact of Blood Draw Time. **a**. Fuji plot of 673 significant metabolite-trait associations (51 metabolites across 58 health traits) with adjustment of blood draw time. The outermost track displays health trait categories, and their corresponding legend color. The ‘Other’ trait category comprises of Dental, Sleep, Respiratory, Neuropsychological, and Quality of Life traits. The top five or top one most significant metabolite(s) (according to p-value significance) are annotated in the outermost track with their name, with the colored square indicating metabolite type. Metabolite types are separated each into a designated separate scatter plot track, with the tracks colored according to the legend in the bottom left. The bar plot in the upper right quadrant showcases total metabolite-trait associations identified, according to metabolite type. The following abbreviations are used: PUFA: polyunsaturated fatty acid, MUFA: monounsaturated fatty acid, SFA: saturated fatty acid, and FA: fatty acid. **b**. Natural log-scaled ratios comparing metabolite-trait association effect sizes from models adjusting for blood draw time, to models not adjusting for blood draw time. **c**. Non-zero counts of metabolite-trait associations which exhibit directional disagreement in effect size estimates between models.

Natural log-scaled effect size ratios across all 4732 association results ranged from -5.5 to 5.1, with 154 results showing opposite effect directions between models (Figure 2b, Figure 2c). 57 significant metabolite-trait associations in models unadjusted for blood draw time, were shown to lose their statistical significance when adjusting for blood draw time (Supplementary Table 20). On the contrary, 47 significant metabolite-trait associations were revealed to have been missed, if models did not adjust for blood draw time (Supplementary Table 21). Assessing 626 serum metabolite-trait associations that exhibited significant trait association regardless of model adjustment for blood draw time, 241 metabolite-disease associations demonstrated a stronger effect size, and 385 a weaker effect size, when accounting for blood draw time (Supplementary Table 22). Filtering metabolite-trait associations to those that exhibit >10-fold p-value difference and a change in statistical significance when comparing results from models with blood draw time adjustment to models without such adjustment highlighted 57 metabolite-trait associations (Supplementary Table 23, Supplementary Figures 9-10).

We next performed simulation analyses to assess potential bias in metabolite–trait associations arising from circadian oscillations of metabolites (Methods). For time-varying traits, naïve models that ignored blood draw sample collection time introduced positive bias when the traits oscillated in-phase with a given metabolite (Supplementary Figure 11). For traits oscillating out-of-phase with a given metabolite, phase differences between 0 and π/2 produced positive bias, whereas phase differences between π/2 and π led to negative bias, with variations in this trend according to the amplitude of oscillation of the metabolite (Supplementary Figure 11). For both scenarios (in-phase, out-of-phase), the blood-draw time adjusted model outperformed the naïve model, yielding effect size estimates closer to the true effect size (Supplementary Figure 11). In metabolome-wide simulations, adjusting for blood draw time reduced both type 1 and type 2 error in in all settings where the outcome and a proportion of the causal metabolites are rhythmic (Figure 3a, Figure 3b). Type 2 error was zero for the naïve model in the scenario of all causal metabolites being rhythmic, due to substantial inflation in effect estimates. These results highlighted the critical importance of accounting for blood draw timing in metabolic association analyses.

**Figure 3.**
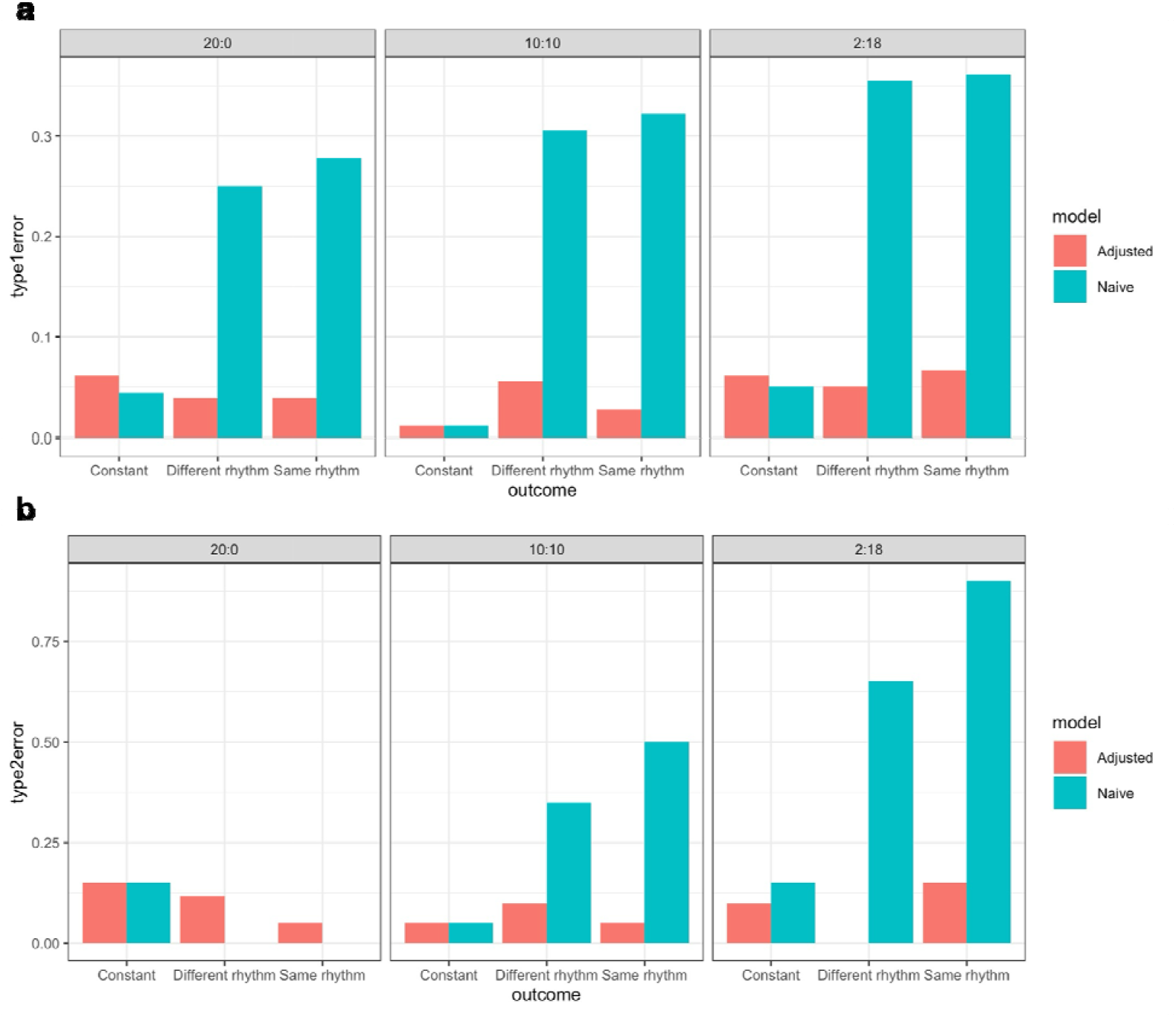
Type 1 and Type 2 Errors of Metabolome-Wide Trait Association Analysis for Metabolites Simulated from a Restricted Blood Draw Time Window. **a**. Type 1 error under α=0.05 across three outcome scenarios (constant, in-phase with metabolite, and π/2 phase-shifted) and three causal metabolite compositions (circadian:non-circadian = 20:0, 10:10, 2:18) from the simulation analyses. **b**. Type 2 error under α=10^-4^ across the same outcome scenarios and metabolite compositions.

## DISCUSSION

Fundamentally, the results of these analyses emphasize that non-negligible temporal patterns underly serum metabolites. Even when blood draw sample collection times span a restricted, short daytime window (7:45 am – 1:38 pm) 84 serum metabolites were identified to be significantly time-varying and 672 metabolites clustered according to potential latent daytime patterns. Time of sample collection appears to be an important factor influencing quantified metabolite measures.

Amongst regression analyses assessing serum metabolite associations with time of blood draw, as a validation check it was confirmed that the well-known circadian rhythmic metabolite cortisol and its precursor cortisone both exhibited a significant daytime decrease trend, matching prior reports of blood cortisol exhibiting a morning peak (∼08:30) followed by gradual decrease ^18^ (Supplementary Figure 2). The fully-adjusted sensitivity model (accounting for food intake time, sleep/wake time, physical activity, diet, substance use, and medication use for underlying health disorders) revealed stronger statistical significance and effect size for cortisol’s association with cosine time of blood draw (p=1.4e-05, *β*=1.8), compared to the primary model (p=1.7e-04, *β*=1.6; Supplementary Table 3). This may reflect the fact that cortisol rhythms are often disrupted in individuals with chronic pain, immune/inflammatory disorders, and depression ^19-21^. In addition to cortisol, the observed daytime trend of glucose (significant positive association with sine time of blood draw, i.e. a morning to afternoon dip) aligned with prior reports of daytime fasting plasma glucose fluctuations ^9^ (Supplementary Figure 2). All 27 metabolites validated in the independent clinical trial (SHIFT) exhibited consistent trends of variation (higher levels in evening, lower levels in evening) according to their observed pattern in the discovery data (sine effect, cosine effect).

Results of the health/disease trait association analyses identified that covariate adjustment for sample collection time can indeed impact effect size estimates as well as strength of association evidence, resulting in both type 1 and type 2 errors. This suggests that for biomarker discovery, model design may benefit from adjusting for sample collection time, potentially improving precision of effect estimates. This was confirmed by the simulation analyses, which showed that models for time-varying outcomes that do not adjust for blood draw time, result in over-estimated or under-estimated effect sizes for circadian metabolite-trait associations. Furthermore, simulations of metabolomics data harboring a subset of circadian metabolites showed that blood draw time-adjusted models have both reduced type 1 and type 2 errors compared to models that do not adjust for blood draw time for temporally varying outcomes. This impact on biased effect size estimates is particularly important to highlight, as many traits and the biomarkers used to define disease outcomes exhibit rhythmic oscillations (e.g. blood pressure, lipids, and glycemia).

Some of the metabolites revealed to be time-varying in this study, have not been prior reported to be rhythmic in literature (Table 1). Of note, both succinoyltaurine and leucylglycine uniquely were the only peptides/amino acids identified to be significantly time-varying in the primary analysis, were clustered together by HDBSCAN, and have not been prior reported to be time-varying. Interestingly, both metabolites appear to carry pathway connections relevant for mood, sleep, and cognitive health. For instance, leucylglycine can be generated from a fragment of the breakdown product of oxytocin, which is a neuropeptide involved in addiction, stress, and sleep-wake patterns ^22-24^. Leucylglycine can cross the blood-brain-barrier, and in the nucleus accumbens induce dopamine rise, suggesting it may protect against proclivity to addiction disorders ^25^. In fact, its cyclic form has been proposed to counter opioid addiction and dopamine agonist drug-induced dyskinesia ^26,27^. Succinoyltaurine is also linked to addiction disorders as it is implicated in the endocannabinoid system by its strong association with rs324420, a missense variant (P: 8.0e-101; Open Targets Platform) for *FAAH. FAAH* is implicated in substance use behavior, sleep duration, sleep quality, and depression risk ^28-30^. Unmodified taurine is an antidepressive molecule, sensitive to sleep deprivation, and enhances activity of serotonin N-acetyltransferase (rate-limiting enzyme in melatonin synthesis) ^31-33^.

The secondary age-stratified and sex-stratified linear regression analyses also identified unique insights, that may be important for screening or prediction models. Of note, diacylglycerols were significantly associated with time of blood draw only in the male-specific analysis. Some of these diacylglycerols (oleoyl-linoleoyl-glycerol, linoleoyl-linoleoyl-glycerol, palmitoyl-linoleoyl glycerol) have been previously utilized in obstructive sleep apnea risk prediction models or exhibit sensitivity to sleep duration or disrupted sleep-wake schedules ^34-36^. Similarly, acylcholines (docosahexaenoylcholine, dihomo-linolenoyl-choline) were revealed only in the analysis restricted to older adults. Long chain unsaturated acylcholines are proposed to endogenously inhibit the acetylcholine signaling system^37^. Given that reduced acetylcholine signaling is associated with aging, and may link to worsened inhibitory control, if utilizing these metabolites for cognitive impairment risk screening, it may be of interest to understand their temporal patterns alongside their age-specific trajectories ^38,39^.

When considering the results from our pooled and stratified regression models, it is important to note we transformed blood draw time into its sine and cosine components, as even in a short daytime window, cosine captures a changing curvature whereas sine captures a linear decrease – trends that a decimal 24-hour representation may not adequately capture (Supplementary Figure 1). Illustrating this, the unsupervised clustering analysis confirmed metabolites can exhibit both nonlinear and linear trajectories (Figure 1e). In addition, the overall mean squared error was identified to be lower in primary regression models encoding time of blood draw as sine/cosine terms versus models encoding time of blood draw as a decimal (Supplementary Table 24).

Pathway enrichment analyses interestingly identified significant enrichment in both incretin hormone and light pathways – pertinent especially for sleep-wake patterns and glycemic regulation. This is supported by the fact that polyunsaturated fatty acids can act as ligands for *GPR40* or *GPR120* receptors, promoting the intestinal release of glucagon-like peptide-1 (GLP-1) and gastric inhibitory polypeptide – incretin hormones known to have important roles in satiety and pancreatic insulin release (Reactome R-HSA-381771). In addition, long-chain fatty acids can act as substrates for retinol esterification, enabling transport and storage of vitamin A, critical for light perception (Reactome R-HSA-2187338) ^40^. Vitamin A is important for circadian rhythm maintenance, exemplified by its deficiency in animal models shown to alter cyclic patterns (amplitude and phase shifts) of clock genes (i.e. *BMAL1*) in the hippocampus ^41^.

Fatty acid metabolites highlighted to be significantly associated with time of blood draw in this work, are also implicated in other important circadian clock-related pathways. For example, erythrocyte levels of oleic acid are linked to *CLOCK* genetic variation ^42^. Melatonin acts against oleic-acid induced triglyceride accumulation, stearic acid-induced β-cell senescence, and palmitic acid-induced disruption of glucose uptake and hypothalamus *BMAL1* and *CLOCK* gene expression ^43-47^. Alpha-linolenic acid and linoleic acid intake help protect against cortisol rise in rodents exposed to stress, and an inverse relationship between red blood cell membrane docosahexaenoic acid (DHA) and cortisol levels have been observed in major depressive disorder and schizophrenia patients ^48-50^. DHA can protect against palmitate’s circadian disruption of *BMAL1* gene expression in the hypothalamus, and as a *SIRT1* activator may revert palmitate-induced *PER2* suppression and *BMAL1*-*CLOCK* destabilization in hepatocytes ^51-53^.

Network and partial correlation analyses of the time-associated metabolites also revealed pertinent insights. Both tetradecanedioate and hexadecanedioate reflected a partial correlation value of 0.47 – interestingly aligning with them both being potential suggested biomarkers for OATP1B1 inhibition (e.g. by rifampicin, cyclosporin) to assess drug-drug interactions ^54,55^. Another notable result was the identified putative connection between palmitate and *TACR1* in the integrated gene-metabolite network analysis. *TACR1* is a gene linked to morning chronotype by rs12464387 (P: 1.0e-09; Open Targets Platform), and encodes the receptor for substance P, a mood regulating neuropeptide. Palmitate appears to be relevant to its activity, as the absence or addition of palmitate (palmitoylation) of CaVβ2a directs the fate of the *TACR1* activation-triggered calcium signaling, determining whether the N-type calcium current is enhanced, or suppressed ^56^. This may be important for sleep behavior and serotonergic modulation because *TACR1* activity is implicated in regulating wake after sleep onset and sleep latency in insomnia, and effects of citalopram, a selective serotonin reuptake inhibitor (SSRI) ^56-58^. SSRIs may also impact palmitoylation, as escitalopram has shown to reduce palmitoylation of the serotonin transporter SERT, a key regulator of serotonin tone, and in turn mood and sleep^59^.

Limitations of this study include the following to be considered. First, the internal clock was approximated using self-reported sleep and wake time. Assessing alignment between endogenous rhythms and external cues (e.g., sleep, exercise, and food intake timing) is important to more granularly identify the impact of time on metabolite measures. Second, a restricted time window does not reveal circadian rhythm. Third, as the analyzed metabolomics dataset is from overnight fasting individuals, the observed increases in some metabolites may reflect compensatory mechanisms in response to declining glucose or other metabolite levels.

In conclusion, this study reveals that even in a short daytime window, nonnegligible temporal variation exists in serum metabolites. Pathway connections of time-varying metabolites were identified in key areas ranging from depression to acetylcholine signaling. Findings from our study fundamentally suggest that in a cross-sectional analysis framework using untargeted metabolomics data – relevant to the rapidly evolving metabolomics field in trait association analyses, prediction models, and disease biomarker discovery – time of sample collection appears to be a source of variation that should not be ignored. Through covariate adjustment, accounting for this technical variable may improve discovery, protecting against signal obscurity and bias in biological findings.

## METHODS

### Data

Hispanic Community Health Study/Study of Latinos (HCHS/SOL) is a prospective, community-based cohort comprising of individuals who were aged 18-74 years at baseline (2008-2011), self-identified as Hispanic/Latino, and recruited in a multi-stage stratified random sampling manner across Bronx NY, Chicago IL, Miami FL, and San Diego CA ^60^. Serum metabolite concentrations from baseline fasting blood samples retrieved during the day were quantified by the Metabolon, Inc. Discovery HD4 platform (untargeted liquid chromatography-mass spectrometry) in two batches in 2017 and 2021 after storage at -70°C in the HCHS/SOL Core Laboratory. Preprocessing of this retrieved serum metabolite data was as follows. Metabolites were restricted to those with <=75% sample-wise missingness, rank-based inverse normal transformed, and set to missing if an outlier (>5 standard deviations from mean). Following this, missing xenobiotic values were set to 0 and other missing metabolite values imputed ^61^. Multiple imputation was utilized via the R mice package across 5 iterations, with covariables being top five correlated metabolites, age, sex, body mass index, waist-hip ratio, total cholesterol, high-density lipoprotein cholesterol, low-density lipoprotein cholesterol, triglycerides, fasting glucose, fasting insulin, hypertension status, statin use, and diabetes status ^7,61^. This imputed serum metabolite dataset was merged with weekly-average self-reported wake times, weekly-average self-reported bed times, and blood sample draw time. Time measures were radian-transformed (2π x time/24), and encoded as sine and cosine terms (*sin*[radians], *cos*[radians]), with 0 radians corresponding to midnight. Lastly age, study center, sex, self-reported Hispanic/Latino background (Dominican, Central American, Cuban, Mexican, Puerto Rican, South American, Mixed/Other), and HCHS/SOL sampling variables (stratification ID, primary sampling unit ID, weights) were retrieved. The final resulting dataset with both non-missing covariate data and metabolomics data for discovery analyses comprised of 853 fasting serum metabolites across 5954 samples, with blood draw sample times ranging across 7:45 am to 1:38 pm.

Shift Work, Heredity, Insulin, and Food Timing Study (SHIFT) is a clinical trial to determine whether nighttime eating that coincides with elevated endogenous melatonin impairs glucose tolerance (https://clinicaltrials.gov/study/NCT02997319). This cohort comprised of 82 non-Hispanic white individuals who did not work night shifts and reported no medical conditions or medication use (except for oral contraceptives). All participants underwent an 8-hour fasting protocol. For each participant, samples from T0 and T120 from both morning and evening OGTTs were analyzed. Metabolomics profiling was conducted using ultrahigh-performance liquid chromatography-tandem mass spectrometry at Metabolon (Metabolon Inc., Morrisville, NC, United States) followed by standard quality control procedures and comparison to Metabolon’s well-validated metabolomics compound library. A total of 1,533 metabolites were identified and quantified, of which 1,194 were named (339 unnamed) and considered. Metabolites with <20% missing data (i.e., including at least 940 named metabolites) were retained for analysis, with missing metabolite values imputed with a value equal to half the minimum of each metabolite measure in the remaining samples. Among these, the set of metabolites overlapping with metabolites identified in the discovery HCHS/SOL dataset to be significantly associated with time of blood draw, were assessed for significant change in levels between morning and evening times in the SHIFT study, in order to further assess temporal patterns in a separate, external dataset.

### Regression Analysis

The primary regression model of interest was designated as Y_i_ = B_0i_ + B_1i *_ sin_time_of_blood_drawi_ + B_2i *_ cos_time_of_blood_drawi_ + B_Ci *_ C_i_ where Y_i_ refers to sample serum metabolite measure, and C_i_ refers to the covariate vector consisting of age_i_, sex_i_, study center_i_, Hispanic/Latino background_i_, sin_bedtimei_, cos_bedtimei_, sin_waketimei_, and cos_waketimei_. To appropriately account for HCHS/SOL’s cohort sampling design the survey R package’s svyglm() (with provided sampling unit ID, stratification variable ID, and survey weights) was used. Utilizing survey design weights modeled from the entire HCHS/SOL cohort via svydesign(id=∼PSU_ID, strata=∼STRAT, weights=∼WEIGHT_FINAL_NORM_OVERALL)), regression models were fit in each batch separately (2017, 2021), and imputation datasets pooled via mice R package pool.scalar(). P-value, effect size, and standard error estimates for the one degree of freedom (1_df_) tests H_0_: B_1i_ = 0 and H_0_: B_2i_ = 0 were retrieved. Following this, random effects meta-analysis across both batches for the 853 shared metabolites was conducted using rma.mv() from the metafor R package. Batch was modeled as a random effect, and indicator variables distinguishing sine and cosine effects provided to the *mods* argument in rma.mv(), to enable their joint estimated effect in multivariate meta-analysis accounting for their covariance. To assess the 2_df_ test of H_0_: B_1i_ = 0 and B_2i_ = 0 linearHypothesis() from the car R package was utilized. Resultant meta-analysis p-values were Bonferroni corrected for multiple testing. Significant metabolite for the cos, sin, or joint sin/cos effect of blood draw time (Bonferroni-adjusted p< 0.05) – denoted herein as T* – were annotated for prior reports of rhythmicity ^4^.

Sex-stratified (male, female), and age-stratified (<50, >=50) analyses were conducted in addition to the primary model. For unstratified and stratified analysis three sensitivity analyses were performed, adjusting sequentially for additional covariates: A. time of last meal before fasting (24-hour decimal encoded as sine and cosine terms), B. time of last meal and lifestyle variables (current alcohol use (1/0), current cigarette use (1/0), daily physical activity (MET/min, diet score (HEI-2010)), and C. a fully-adjusted model incorporating time of last meal, lifestyle variables, and medication use (1/0: hormone, sleep, lipid-lowering, cardiovascular, diabetic, anti-inflammatory, psychiatric, and/or respiratory).

### Validation Analysis

In SHIFT, differential levels of metabolites repeatedly measured in morning and evening were evaluated as external evidence of a time-varying nature for a given metabolite. Mean log□ fold changes in metabolite were analyzed using linear mixed-effects models adjusted for age, sex, with a random effect for participant. Significant evidence was determined to be p<0.05/X* where X* is the total number of metabolites (Metabolon IDs) overlapping the significant metabolites in combined sex primary model and sensitivity model analyses.

### Metabolite Clustering Analysis

To test whether an unsupervised approach may capture latent temporal patterns in serum metabolite expression, all 853 serum metabolites were clustered by the density-based clustering algorithm HDBSCAN using Python package hdbscan. The input data comprised of 853 rows across 5954 samples. Imputed values across MICE iterations were averaged, data standardized by scikit-learn Python package StandardScaler(), and dimension reduction conducted by Python package umap. For hyperparameter selection, Python package Optuna was used with 500 trials and the objective function maximizing HDBSCAN relative_validity score (Density Based Cluster Validity score or DBCV score). Hyperparameter search spaces were the following: UMAP n_dimensions [2,3], UMAP n_neighbors: [30, 853/4], HDBSCAN min_cluster_size: [15,100], and HDBSCAN min_samples: [1,100]. Set parameters were UMAP min_dist=0.0, and HDBSCAN cluster_selection_method=‘leaf’.

### Bioinformatics Analysis

Metabolites exhibiting significant association with time of blood draw across group-specific, primary model, and sensitivity models (denoted T**) were used to conduct set overrepresentation analysis on the MetaboAnalyst (v6.0) platform. Enrichment (FDR<0.05) in pathways was assessed for pathways curated in The Small Molecule Pathway Database (SMPDB), Kyoto Encyclopedia of Genes and Genomes (KEGG), or Relational database of Metabolomic Pathways (RaMP-DB). Partial correlations, forming a gaussian graphical model, between the T** serum metabolites were calculated using R function EBICglasso(threshold=TRUE, gamma = 0.5), and any connections with chronotype genes (Supplementary Table 14), or joint pathway enrichment with chronotype genes noted via the MetaboAnalyst Network Analysis tool.

### Time-varying metabolite-Disease Associations

Each metabolite in T* identified by the primary model was analyzed for associations with a 91 traits across 13 categories: addiction (4), anthropometric (4), cardiovascular (16), dental (4), glycemic (8), hematological (10), immune-inflammatory (8), lipids (4), neuropsychology (7), quality of life (2), kidney and liver metabolism (10), respiratory (6), and sleep (8), to characterize the potential clinical relevance (Supplementary Table 25). Associations were tested using the same regression model design as in the primary analysis, with the metabolite as the outcome, the trait as the additional exposure of interest, and adjusting for time of blood draw along with other primary model covariates. Effect sizes and p-value significance were further compared between models with and without adjustment for blood draw time.

### Simulation Analyses

Simulation analyses were conducted to evaluate the impact of rhythmicity on metabolite–trait associations when blood samples are collected at random times. In each simulation, data was generated for 3,000 individuals with a true baseline metabolite level *M*_*true,i*_ ∼ *N*(0,1). Circadian variation was incorporated into the observed metabolite as *M*_*obs,i*_ *M*_*true,i*_ + *Asin* (2*πt*_*i*_ /24 +*ϕ*) + *ϵ*,where *t*_*i*_ *is the o*bservation time (representing blood draw time) within a 24-hour period, *A* is the amplitude of circadian oscillation, *ϕ* is the circadian phase shift, and *ϵ* is the random noise. Outcomes were simulated depending on the true metabolite level: *Y*_*i*_ = *βM*_*true,i*_ + *C* + *ϵ* under three scenarios: 1. Constant overtime C=0; 2. Outcome oscillates in-phase with the metabolite *C* = *A’*sin (2*πt*_*i*_ /24); 3. Outcome oscillates out-of-phase with the metabolite *C* = *A’*sin (2*πt*_*i*_ /24 +*ϕ*). *β* was set to 0.8, representing the true effect association between the metabolite and the outcome. *A* was set to 1.5, *A*^*’*^ (outcome oscillation amplitude) set to 2, and *ϕ* (phase shift between metabolite and outcome rhythms) set to *π*/2. Across the 1000 simulations, for each of the three scenarios, estimated metabolite-trait effect sizes 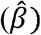 and 2-sided p-values from the naïve model: 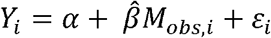 were compared to the blood draw time-adjusted model: 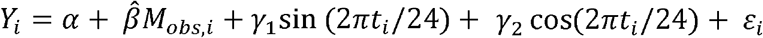.

To assess impact of phase difference between metabolite and outcome oscillations on estimated metabolite-trait association effect sizes, the simulation experiment was repeated across varying levels of *A* between 0 to 2 in increments of 0.25, and levels of *ϕ* between 0 and *π* in increments of *π*/12. Additionally, 500 metabolome-wide simulations were conducted for 200 metabolites measured across 5,000 individuals, of which 30% had circadian variation. Observation times (*t*_*i*_) were restricted to 7:00–14:00. For each outcome scenario, 20 causal metabolites were simulated under three cases: (1) all causal metabolites are circadian, (2) half are circadian, and (3) 10% are circadian. Results from this simulated metabolome-wide association analysis were used to compare type 1 and type 2 errors between naïve and blood draw time-adjusted models.

## Data Availability

HCHS/SOL pseudonymized data are available via controlled-access application to the database of genotypes and phenotypes (dbGaP) under accession phs000810, or via approved data use agreement with the HCHS/SOL Data Coordinating Center (DCC) at the University of North Carolina at Chapel Hill (https://sites.cscc.unc.edu/hchs/).

## Code Availability

Open-source software packages and bioinformatics web servers utilized for this project are detailed in Methods, and can be found here: *mice* (https://github.com/cran/mice), *survey* (https://cloud.r-project.org/web/packages/survey/index.html), *metafor* (https://github.com/wviechtb/metafor), *car* (https://cran.r-project.org/web/packages/car/index.html), *hdbscan* (https://github.com/scikit-learn-contrib/hdbscan), *scikit-learn* (https://github.com/scikit-learn/scikit-learn), *umap* (https://github.com/lmcinnes/umap), *optuna* (https://github.com/optuna/optuna), *MetaboAnalyst* (https://www.metaboanalyst.ca/).

## Acknowledgements

This work was supported by the National Institute of Health (NIH) grants R01HL161012 (to T.S.) and R01HL153814 (to H.W.). Support for metabolomics data was graciously provided by the JLH Foundation (Houston, Texas). The Hispanic Community Health Study/Study of Latinos is a collaborative study supported by contracts from the National Heart, Lung, and Blood Institute (NHLBI) to the University of North Carolina (HHSN268201300001I /N01-HC-65233), University of Miami (HHSN268201300004I /N01-HC-65234), Albert Einstein College of Medicine (HHSN268201300002I /N01-HC-65235), University of Illinois at Chicago (HHSN268201300003I /N01-HC-65236 Northwestern Univ), and San Diego State University (HHSN268201300005I /N01-HC-65237). The following Institutes/Centers/Offices have contributed to the HCHS/SOL through a transfer of funds to the NHLBI: National Institute on Minority Health and Health Disparities, National Institute on Deafness and Other Communication Disorders, National Institute of Dental and Craniofacial Research, National Institute of Diabetes and Digestive and Kidney Diseases, National Institute of Neurological Disorders and Stroke, NIH Institution-Office of Dietary Supplements. The authors thank the staff and participants of HCHS/SOL for their important contributions. The HCHS/SOL Publications Committee has reviewed and approved the content of this manuscript.

## Author Contributions

P.N. and H.W. conducted data analyses, contextual interpretation, and manuscript writing. B.Y, Q.Q., T.S. and H.W. were responsible for data and funding acquisition. T.S. and H.W. provided project supervision. All co-authors participated in study design and methodology, analyses feedback, interpretation of results, and manuscript editing.

