## Supplementary Figures for "The Diurnal Rhythmicity of Untargeted Metabolomic Profiles in Humans"

Pavithra Nagarajan, et al.

### Supplementary Figure 1. Distribution of Blood Draw Timings

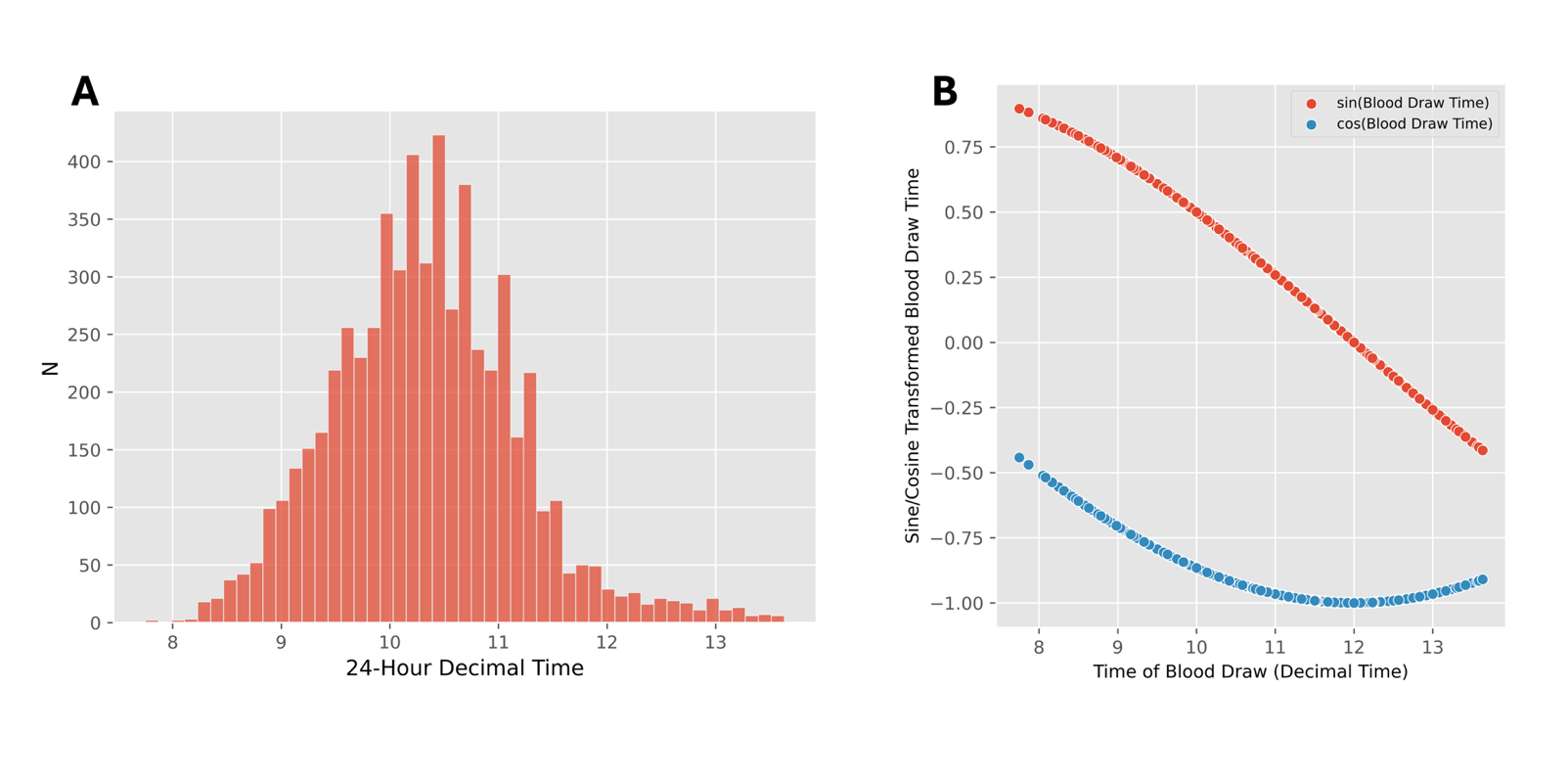
(A) This figure depicts a histogram of the distribution of blood draw timings of 5,954 samples from the HCHS/SOL cohort. The x-axis is in 24-hour decimal time and y-axis total number of samples. Blood draw timings range between 7:45 am and 1:38 pm (mean: 10:20 am). (B) This figure displays the transformation of blood draw time from 24-hour decimal encoding to its sine and cosine counterparts.

### Supplementary Figure 2. Trends of Blood-Draw Time Associated Metabolites

Lowess-smoothed residuals of observed data (ranging between 7:45 am - 1:38 pm) for each of the 52 metabolites significantly associated with time of blood draw in the primary model (Table 1), are plotted here in black. Cortisol is also plotted here, corresponding to the fully-adjusted sensitivity model (Supplementary Table 2). Rolling averages for the observed data are overlaid in the background (light gray) using a 200-sample window with a minimum of 50 non-missing values per window. In pink is the predicted trend curve of the metabolite between 6 am to 6 pm. The orange dashed line is the fitted sine component, and the blue dashed line the fitted cosine component. The x-axis is time of blood draw (24 hour decimal time).

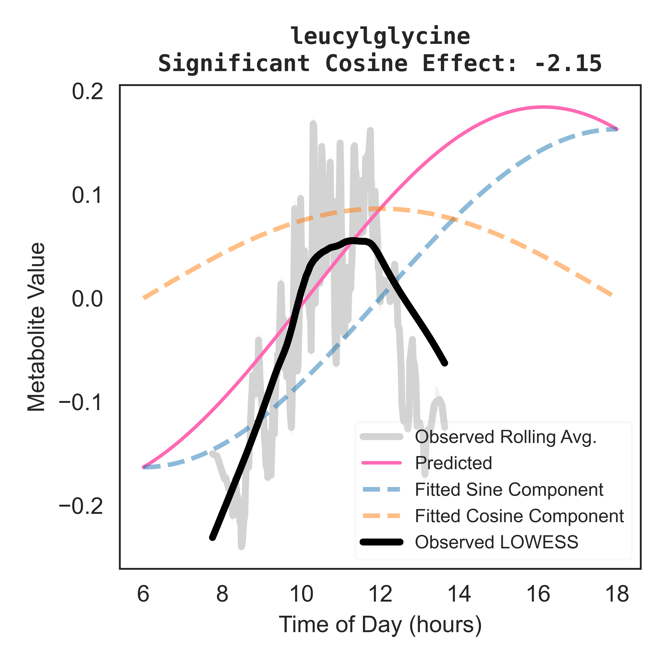

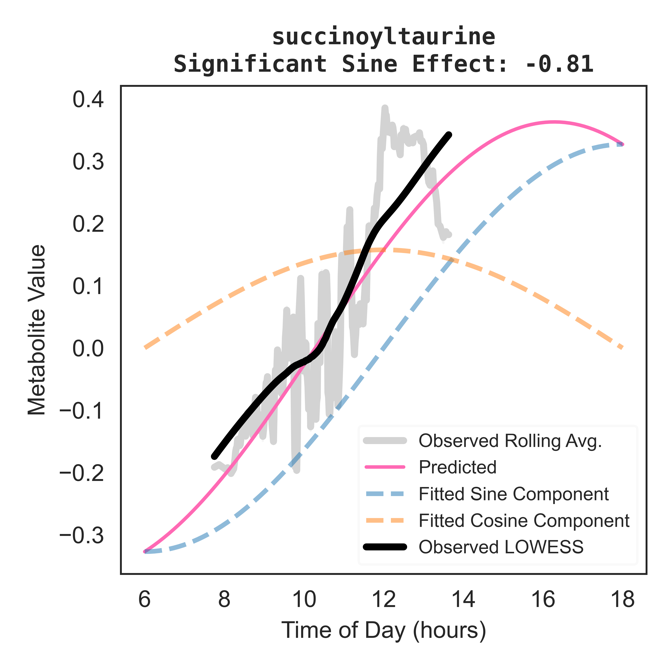

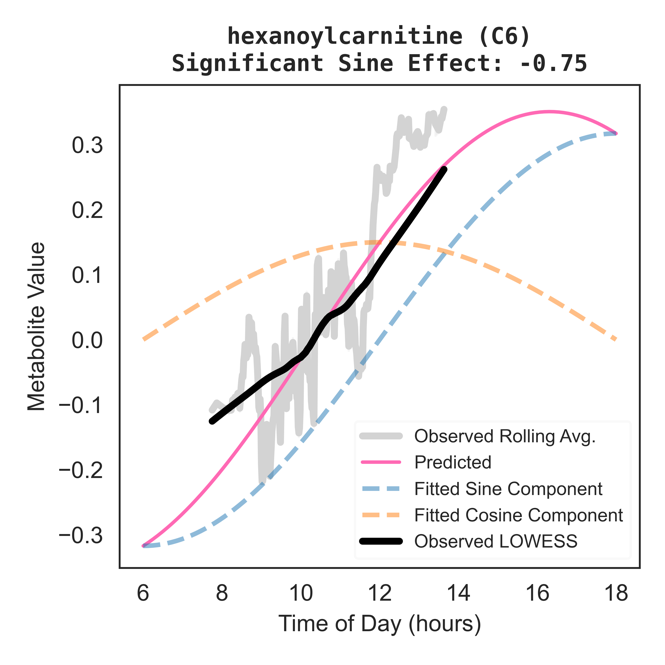

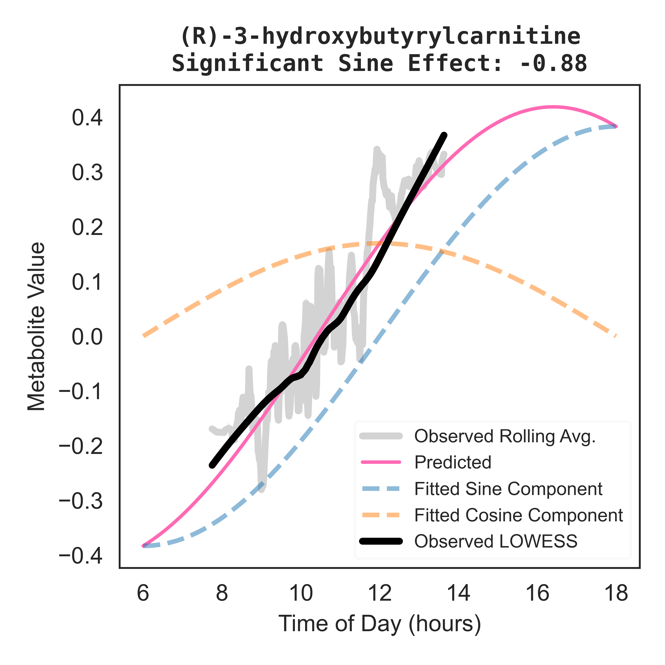

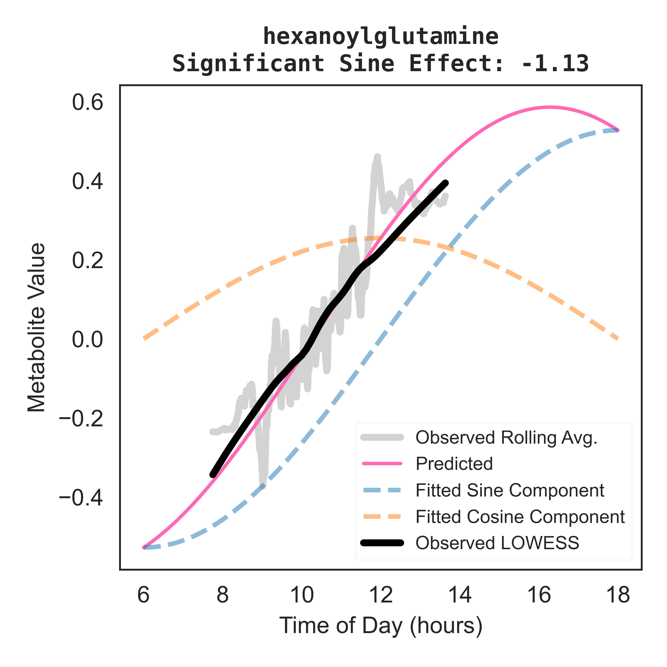

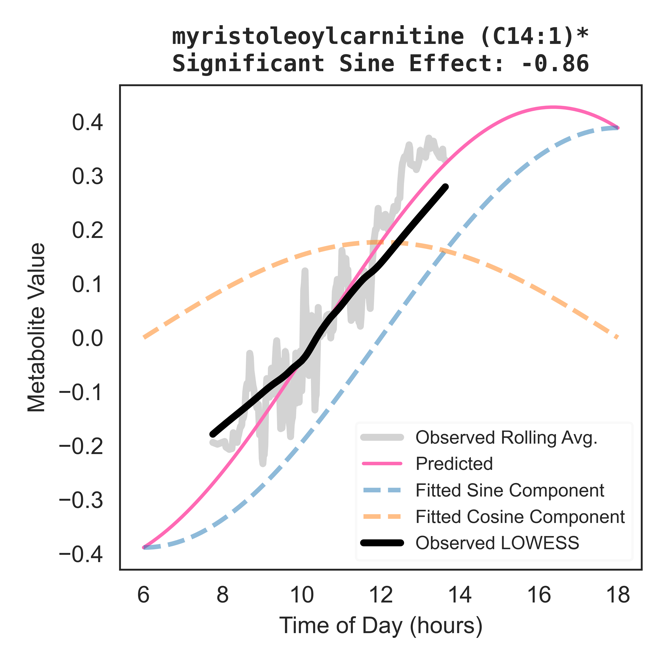

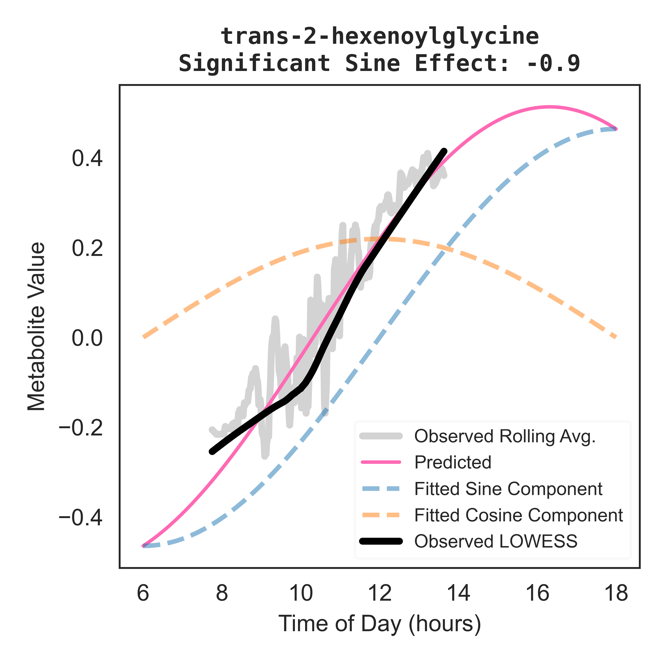

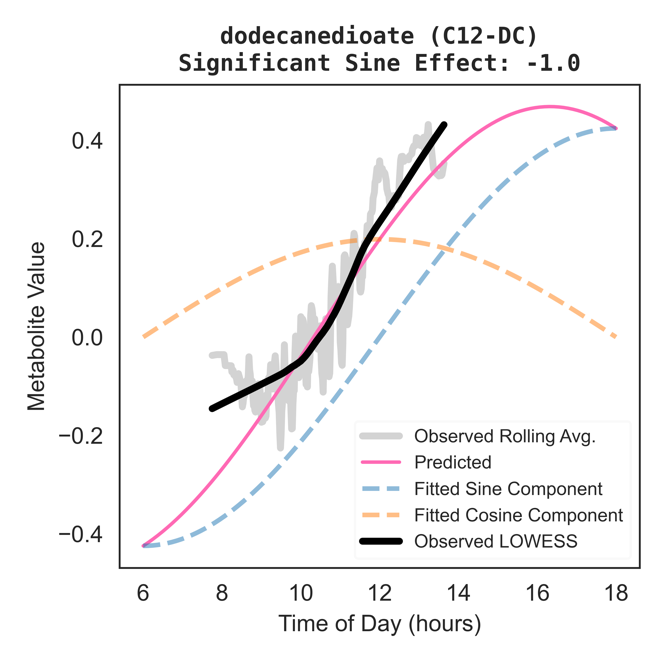

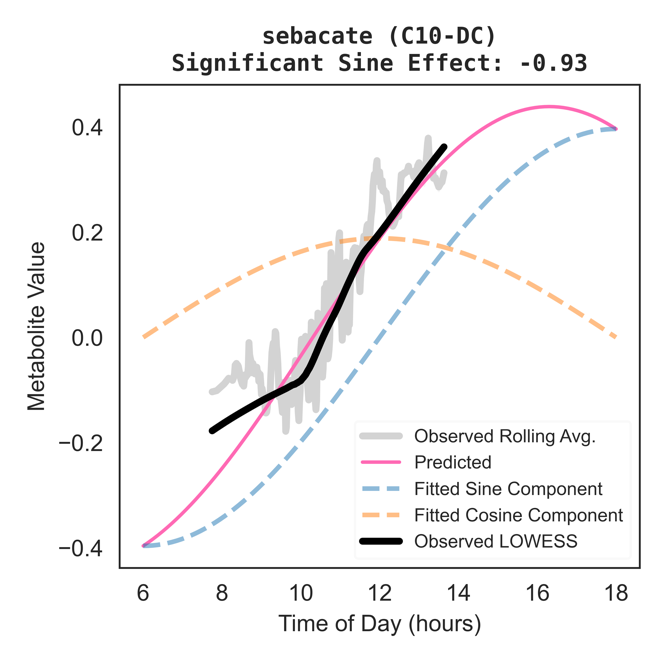

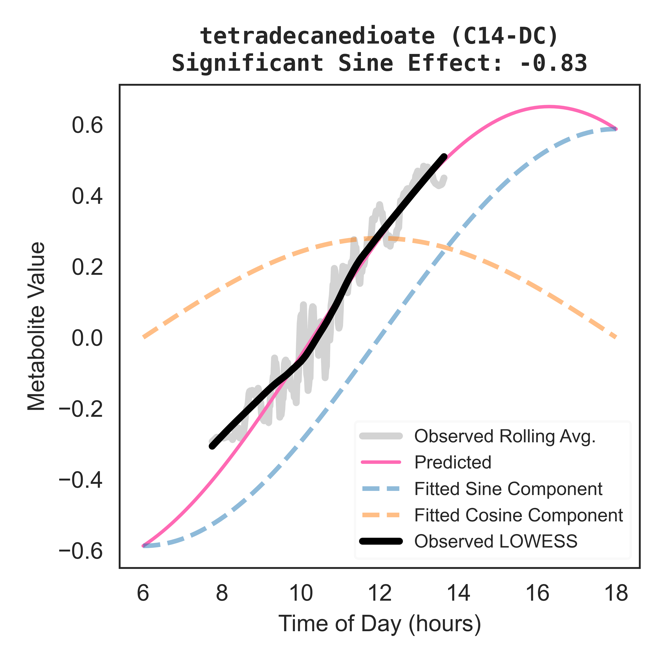

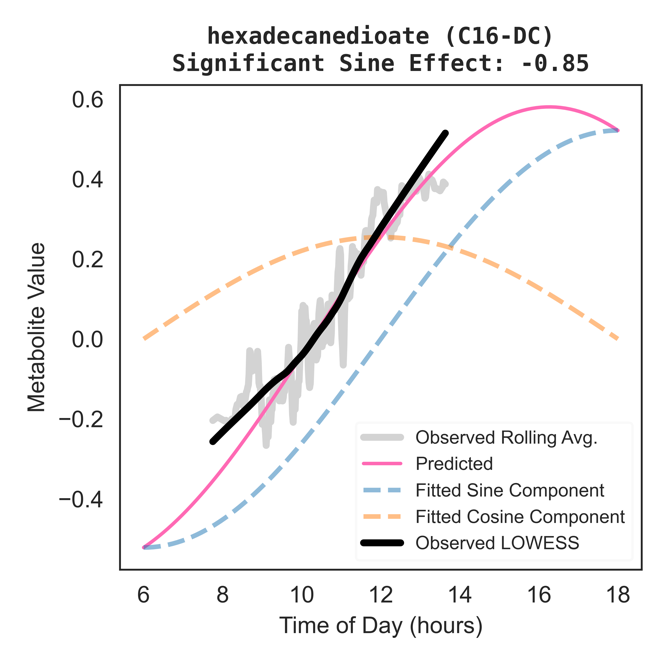

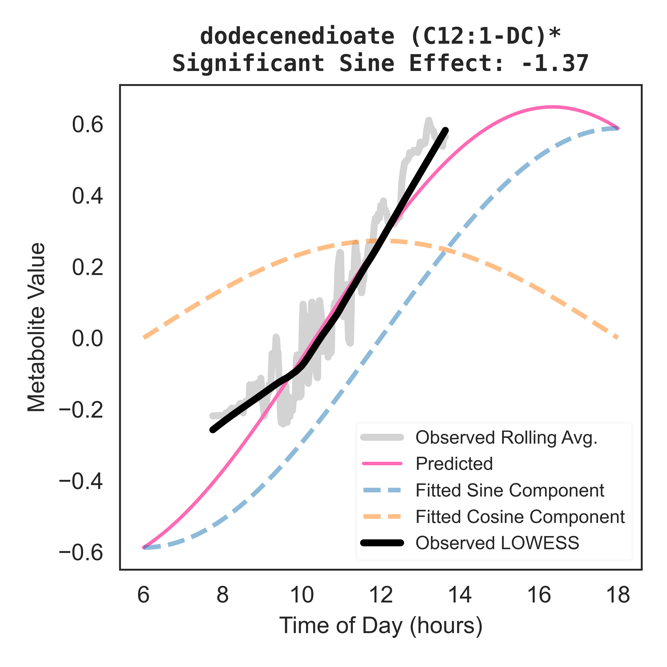

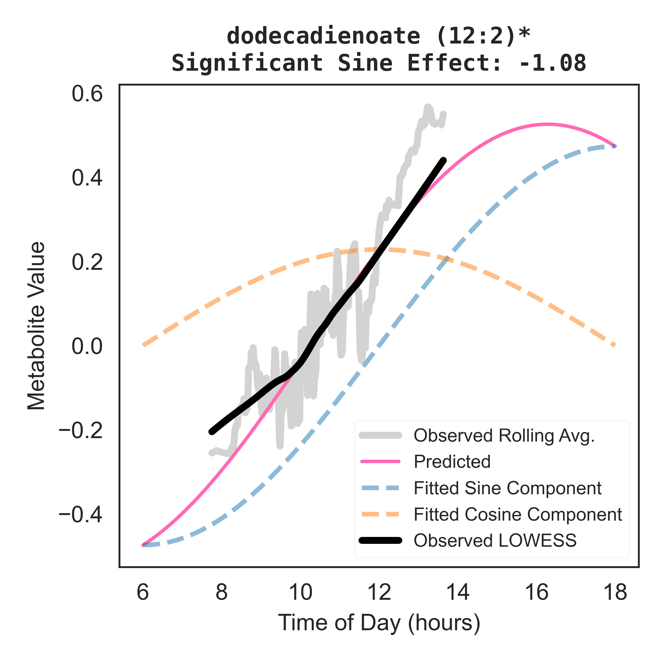

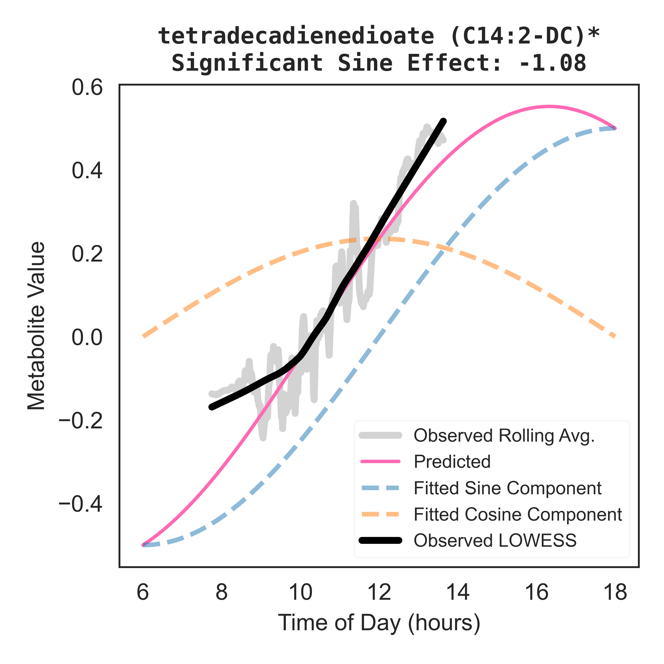

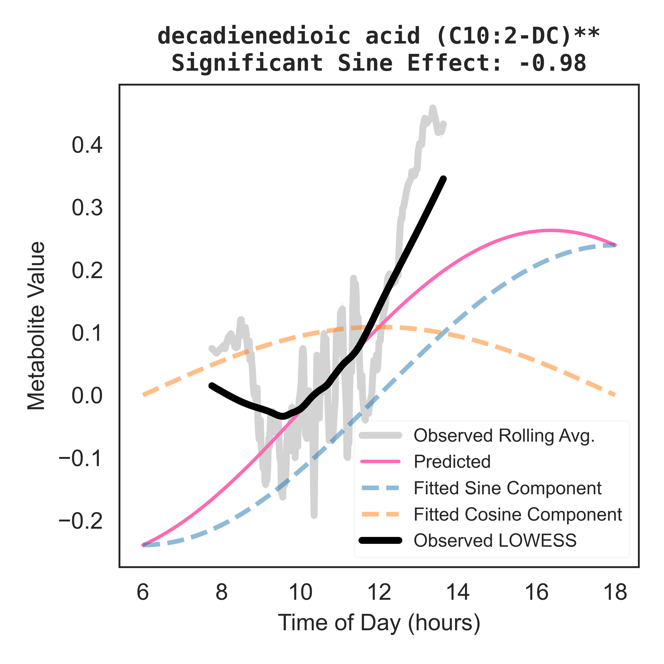

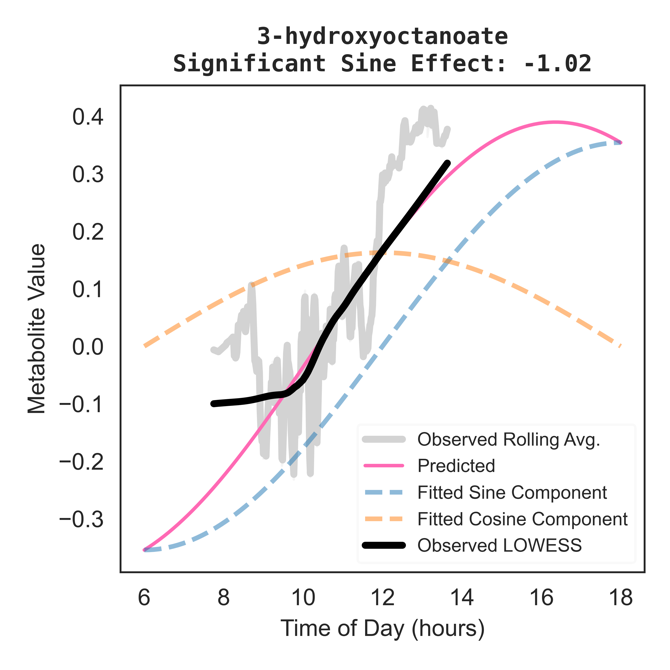

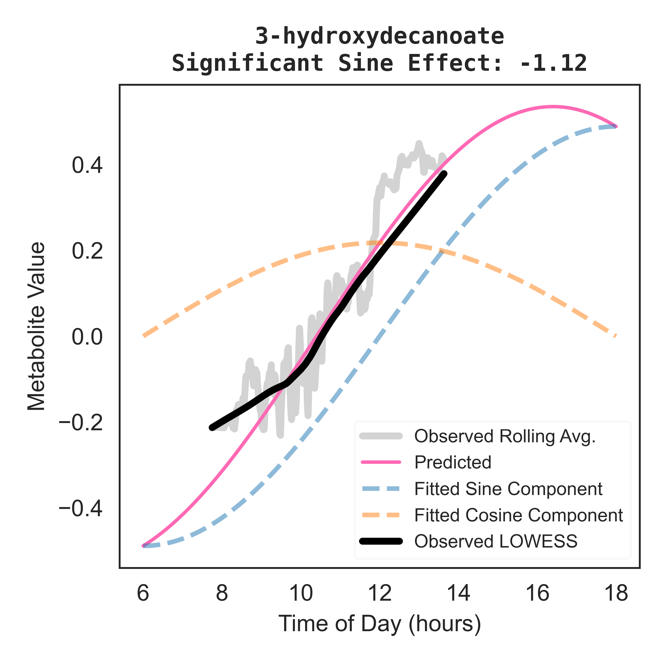

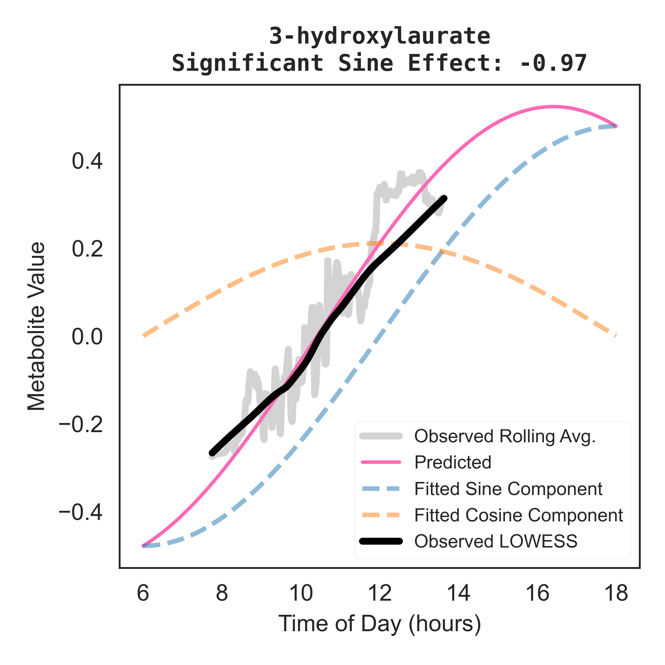

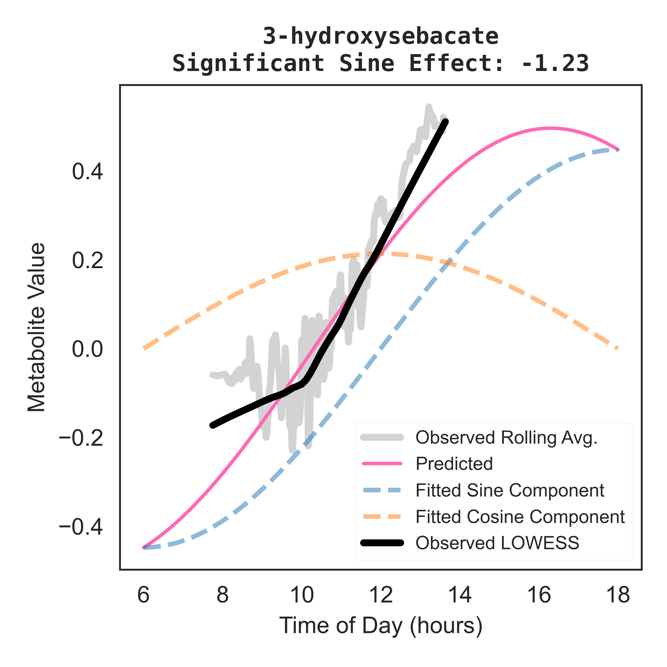

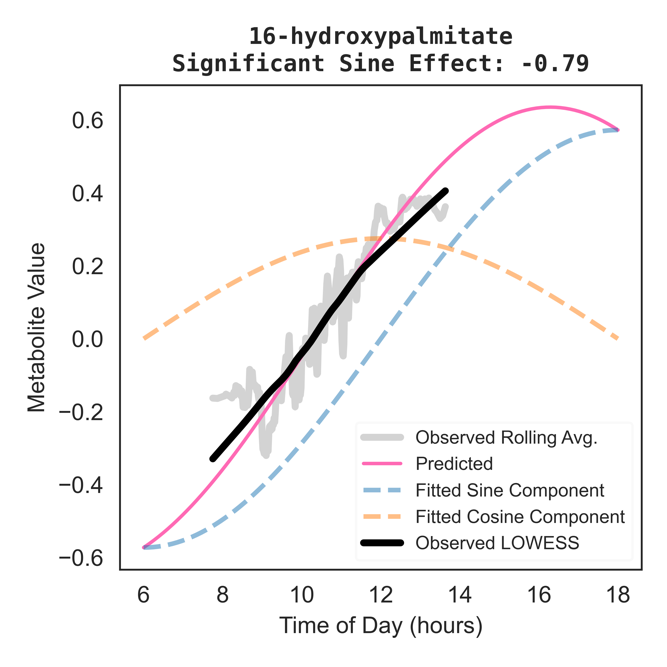

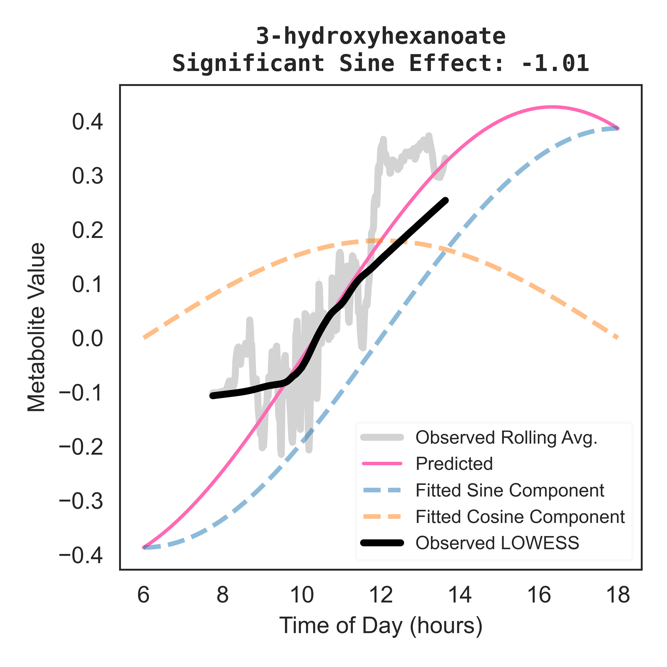

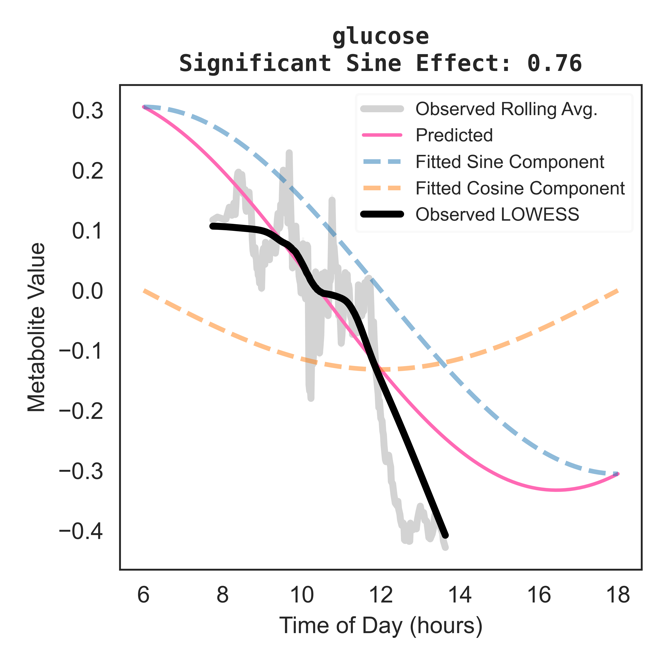

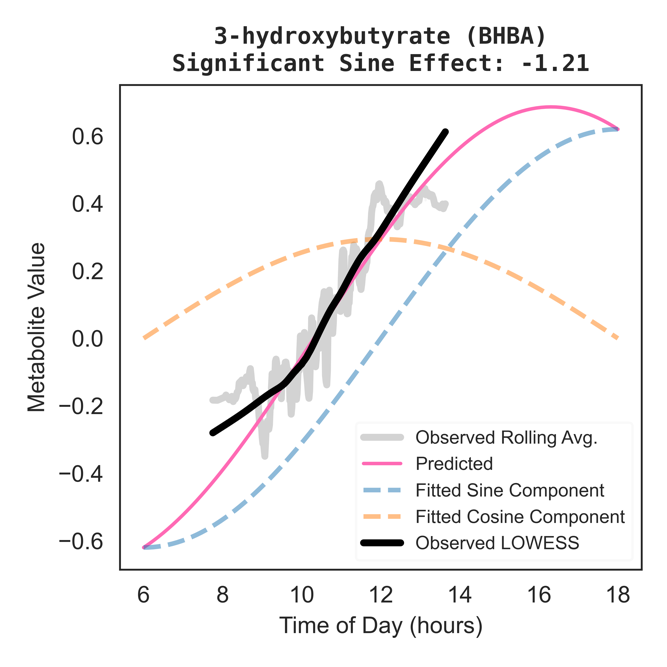

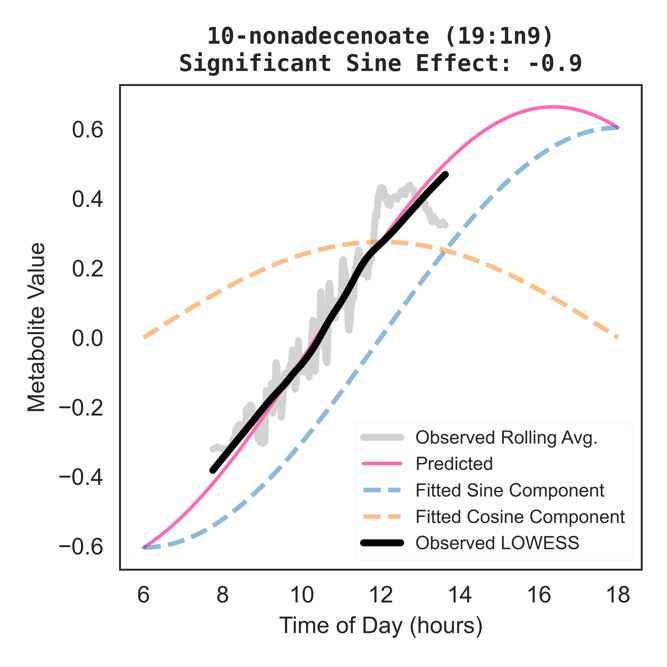

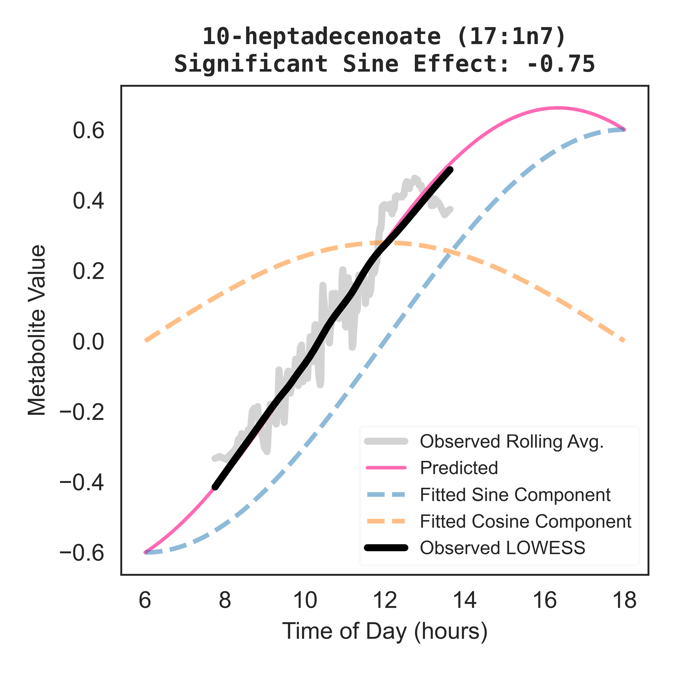

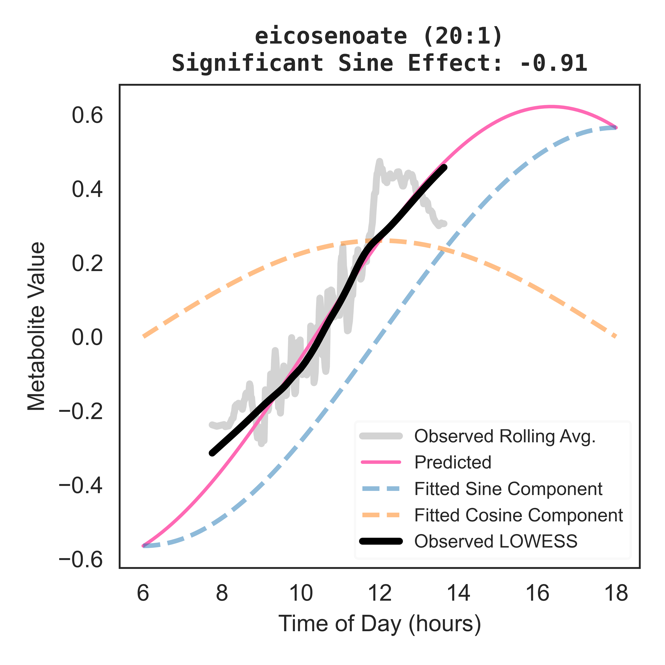

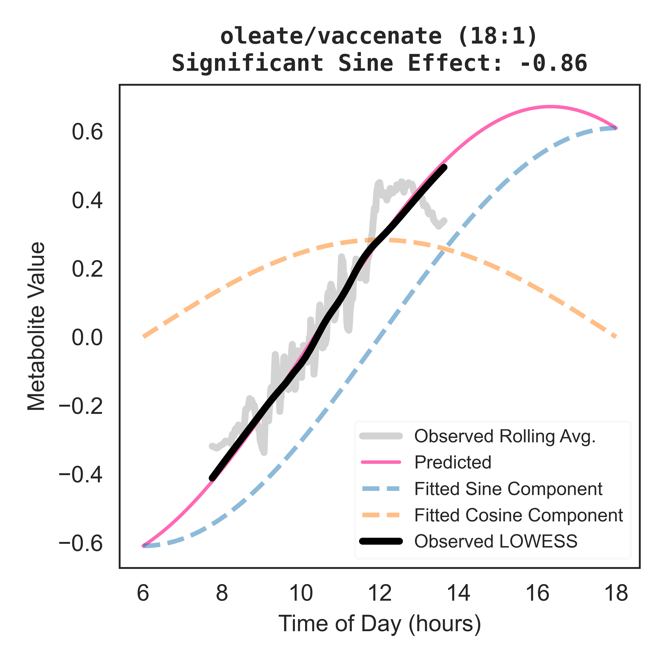

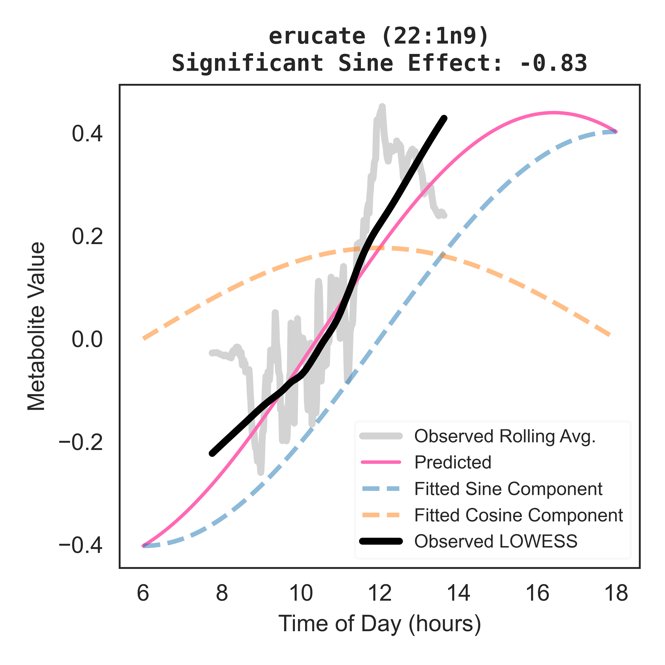

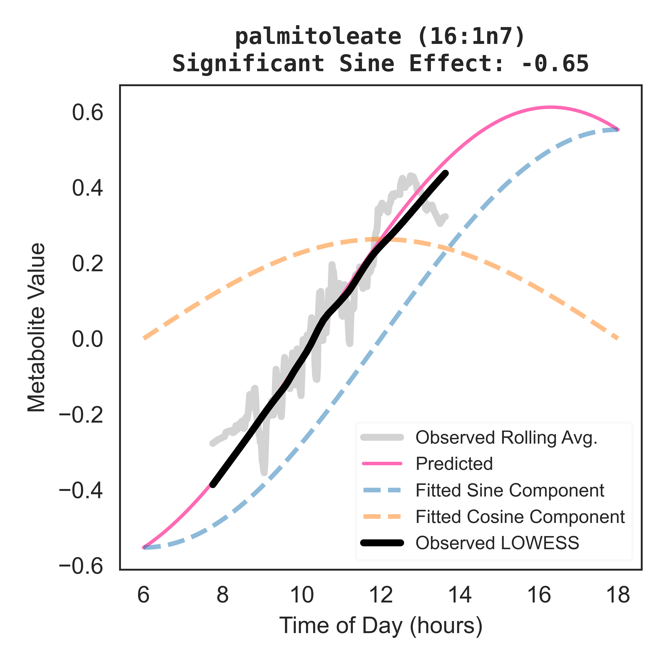

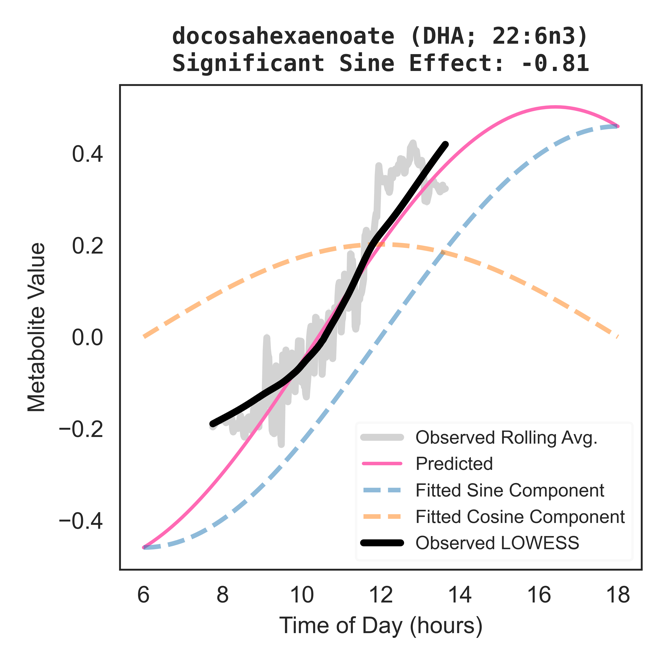

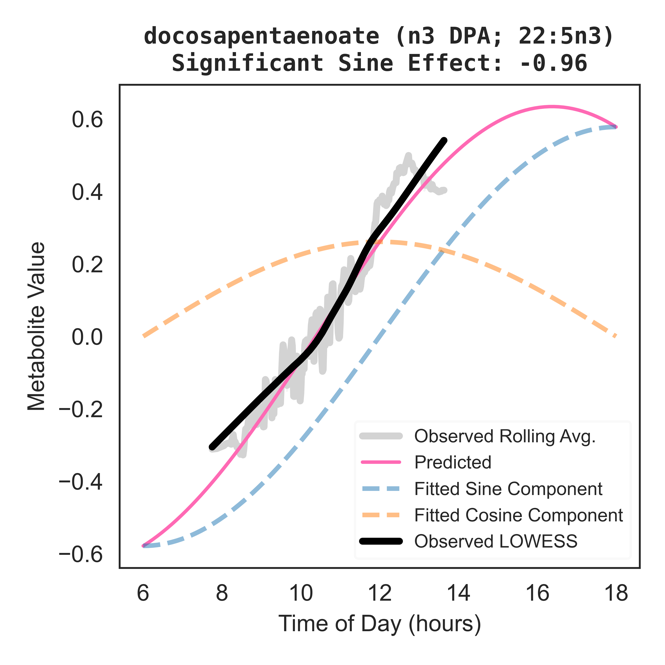

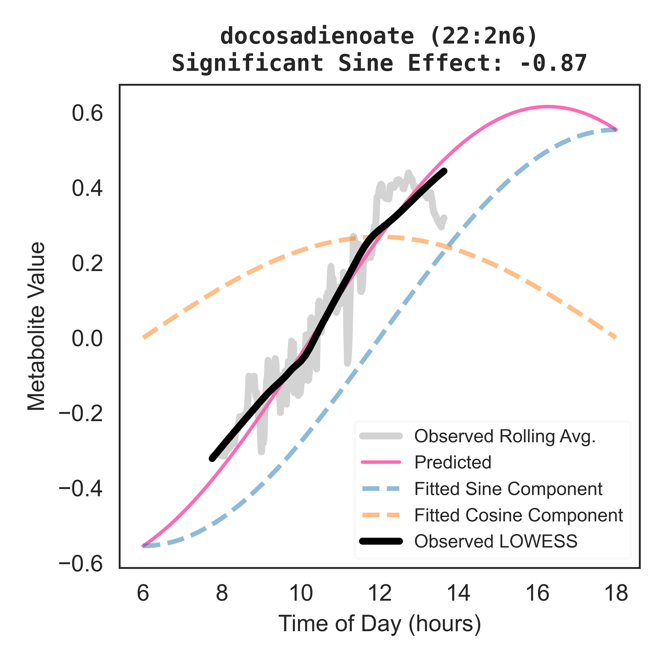

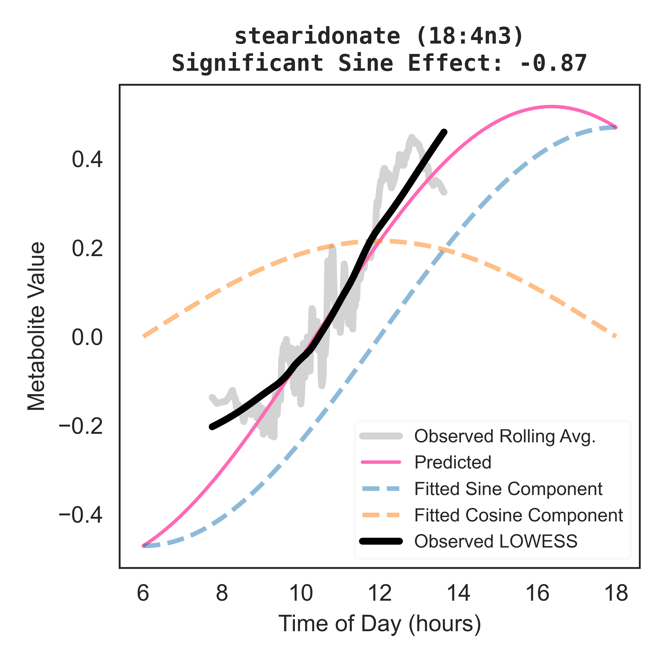

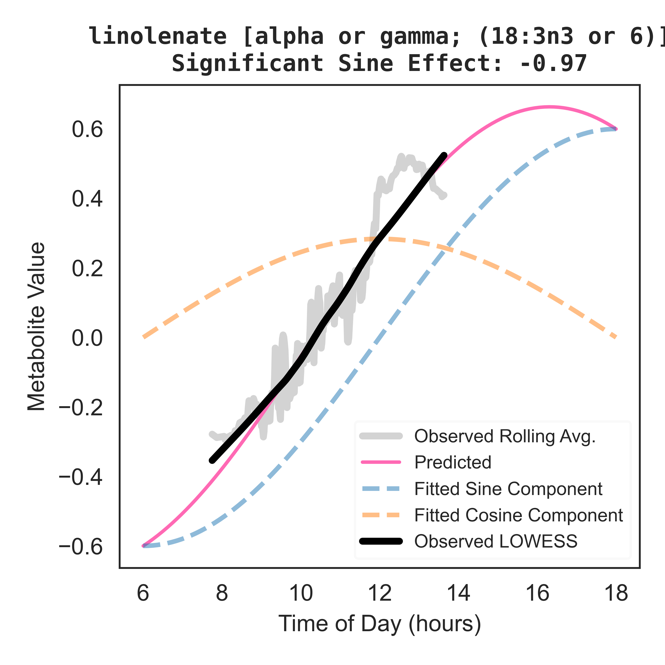

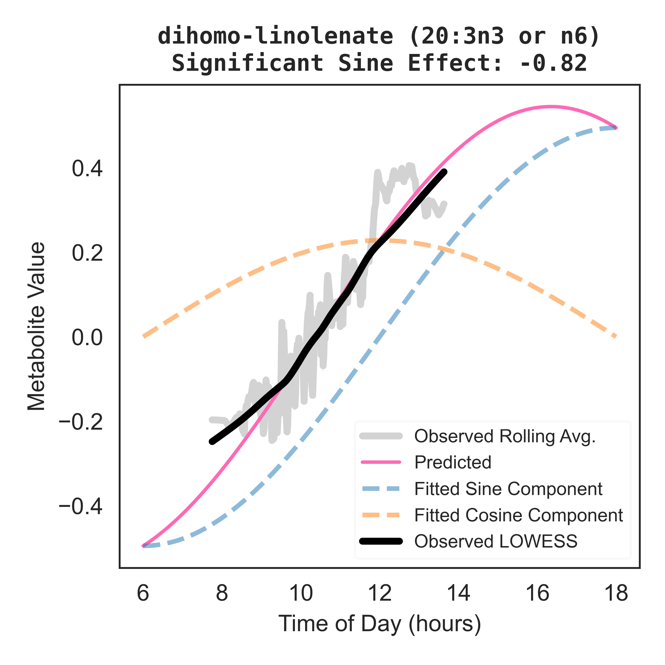

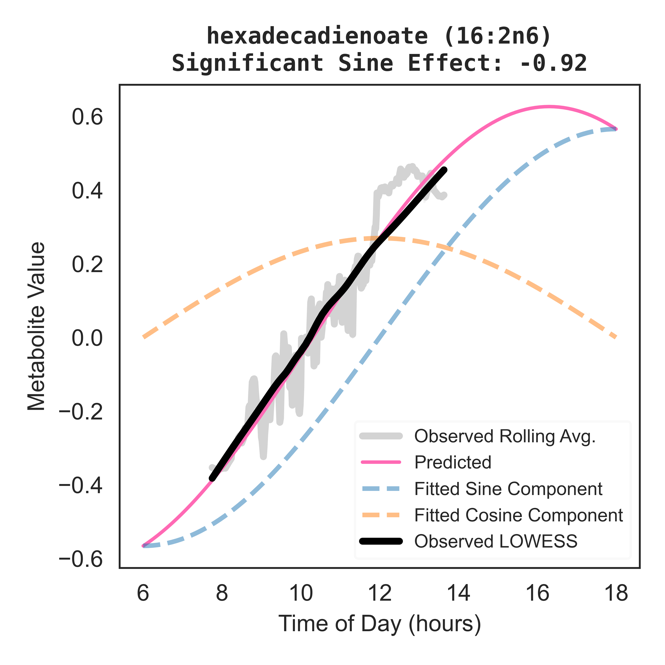

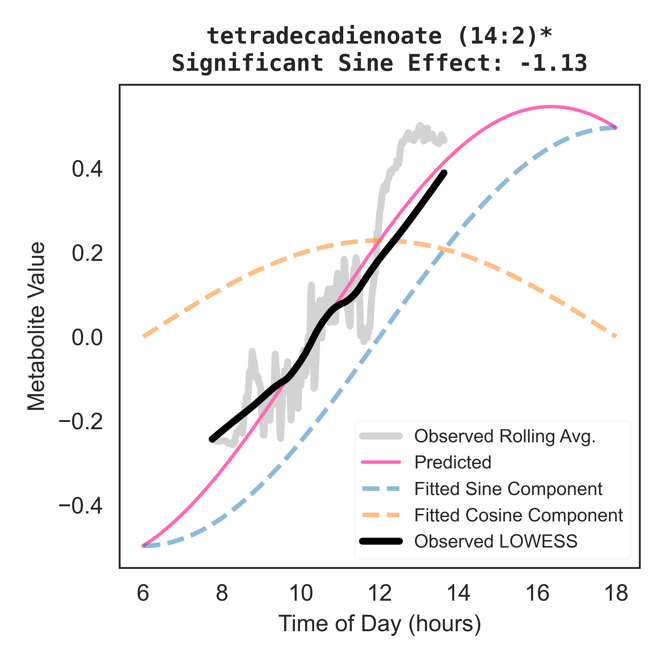

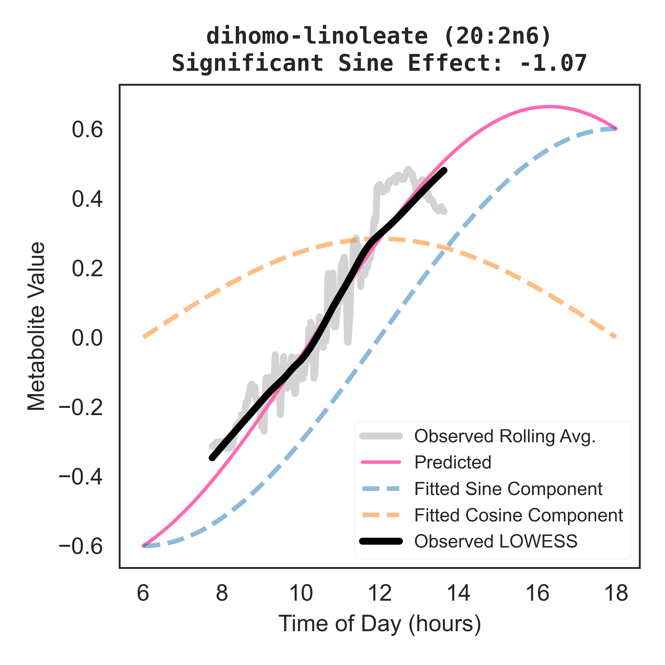

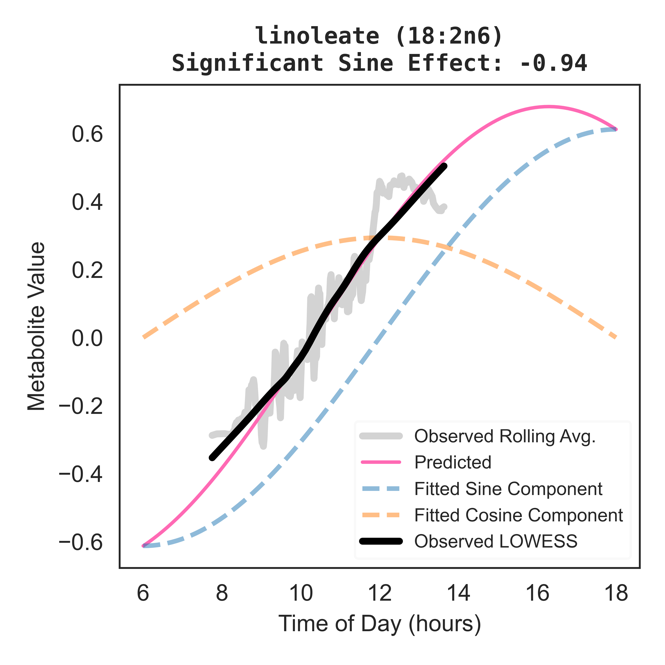

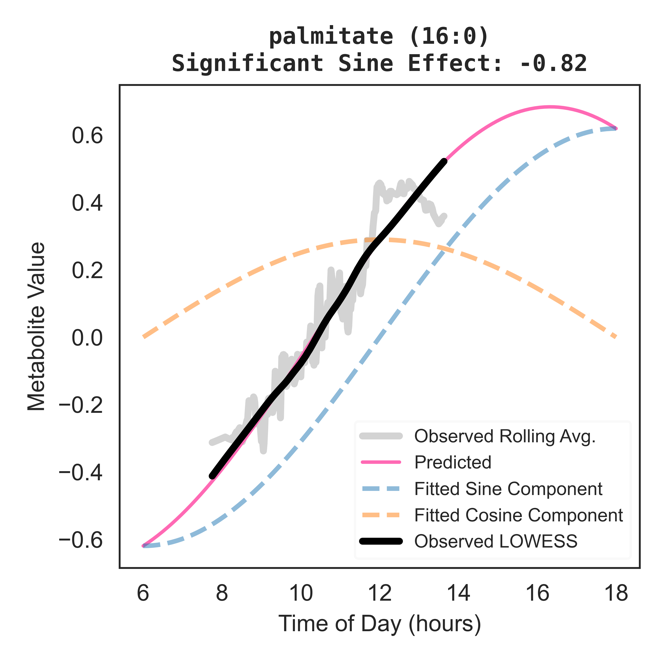

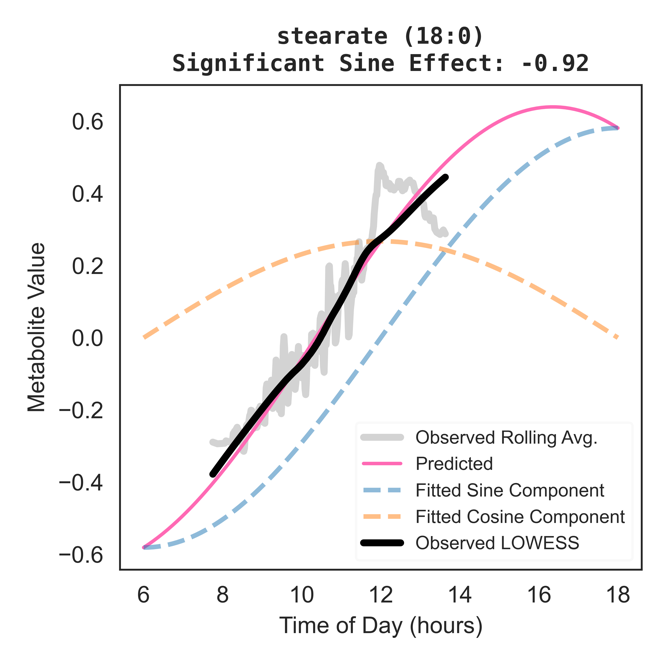

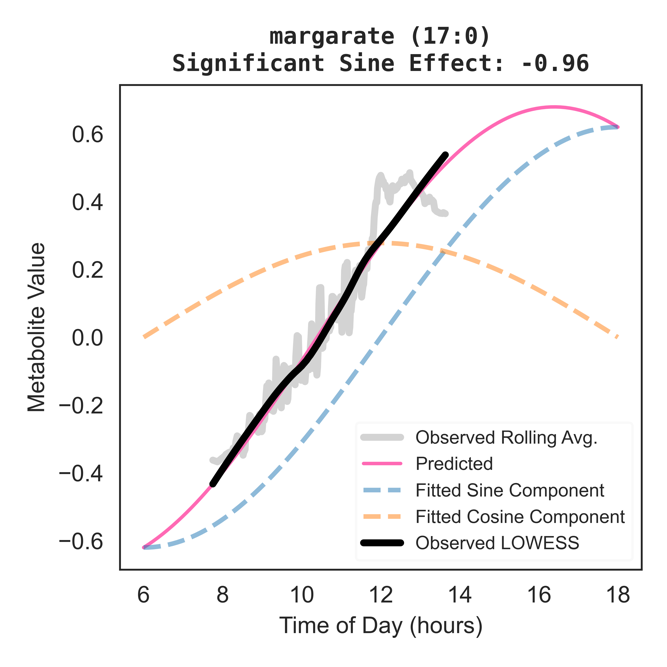

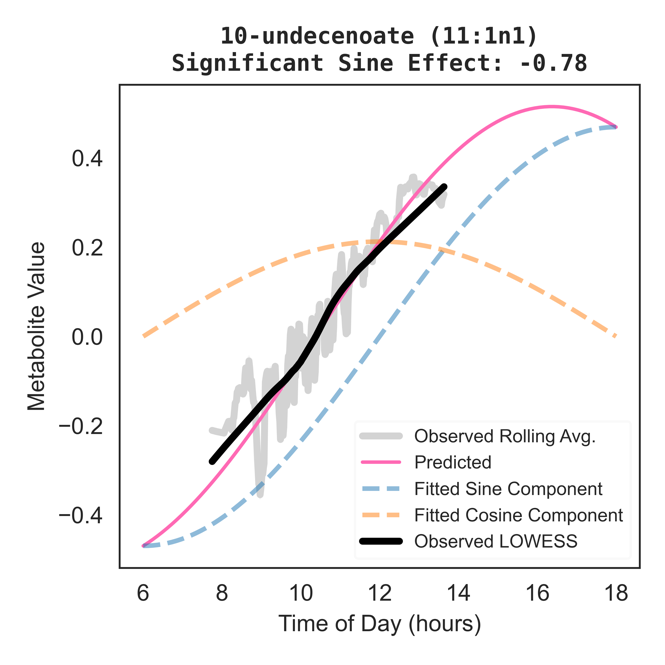

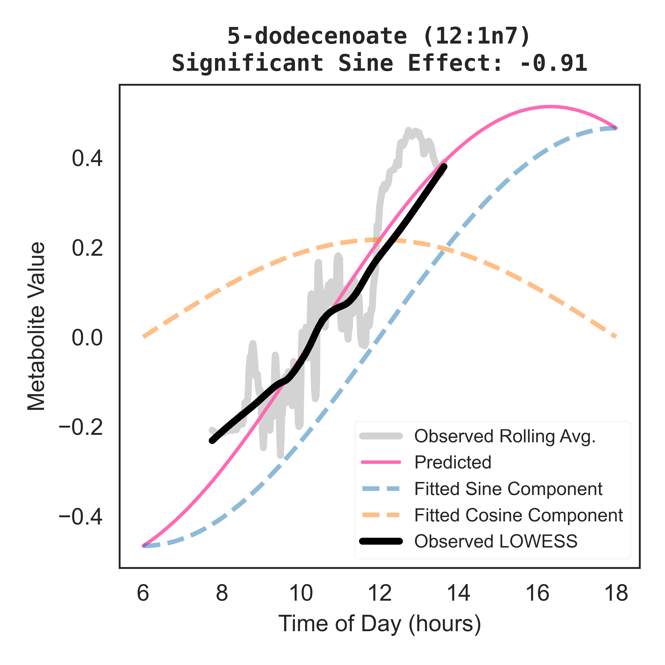

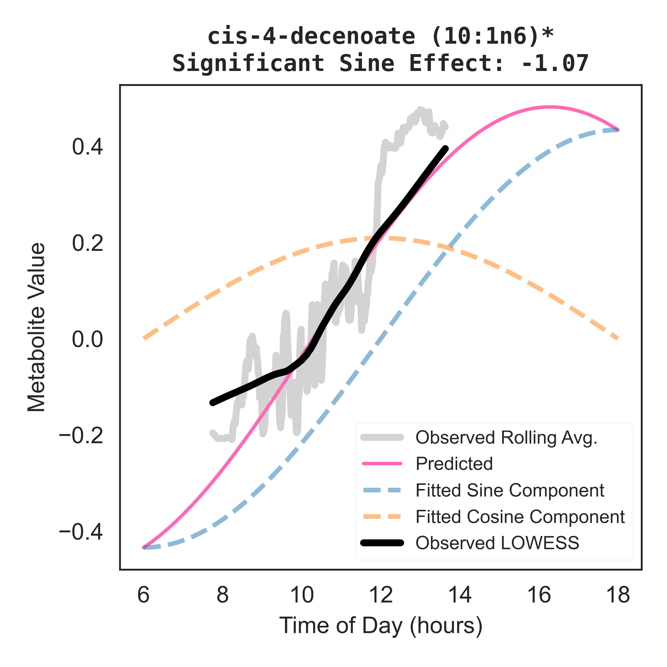

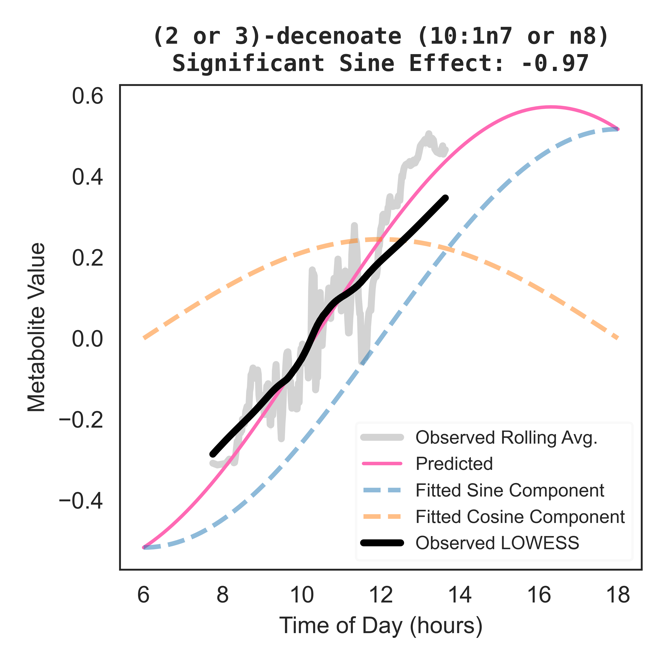

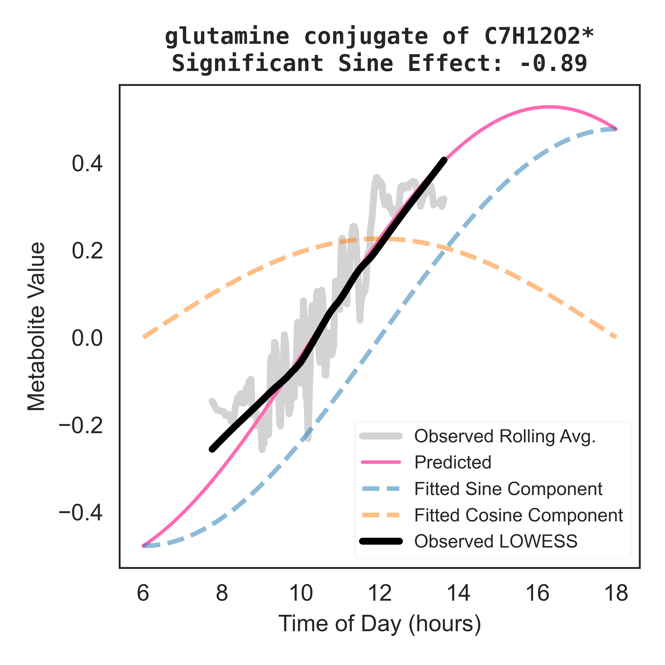

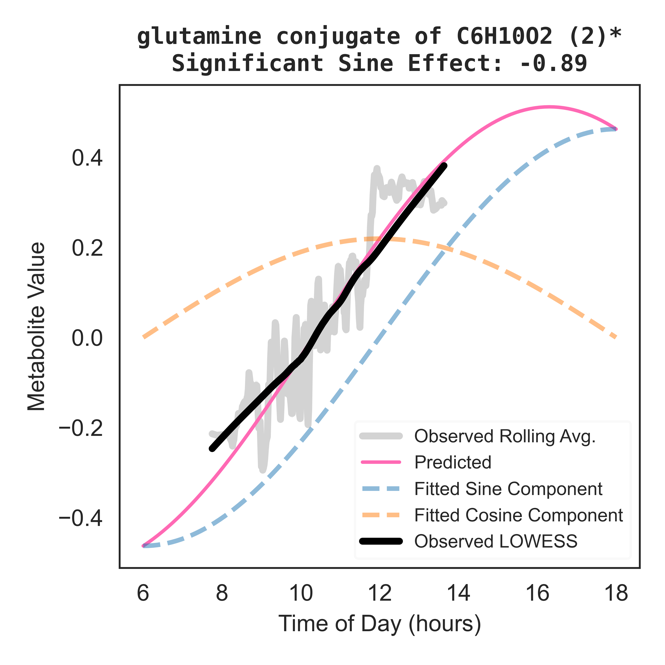

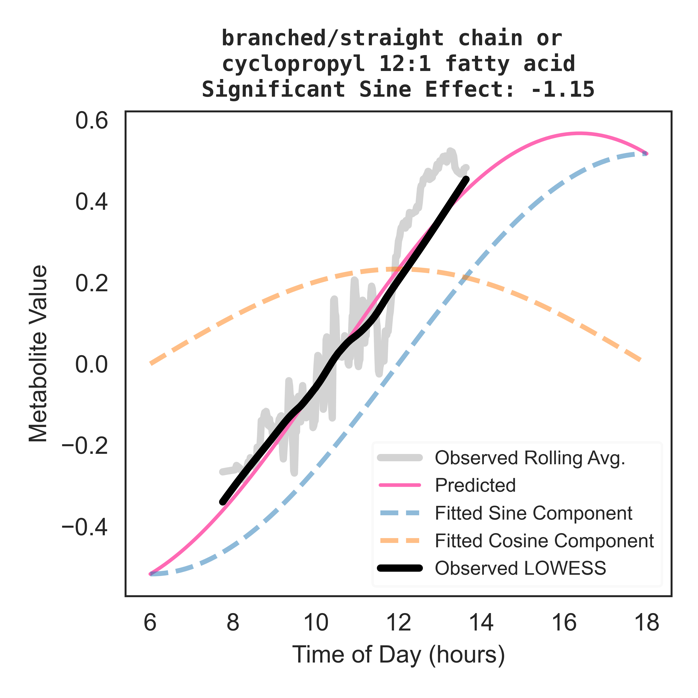

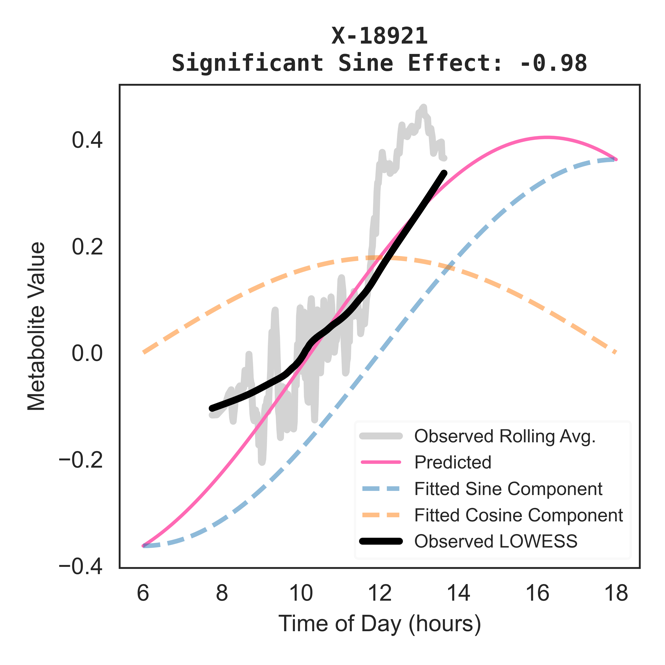

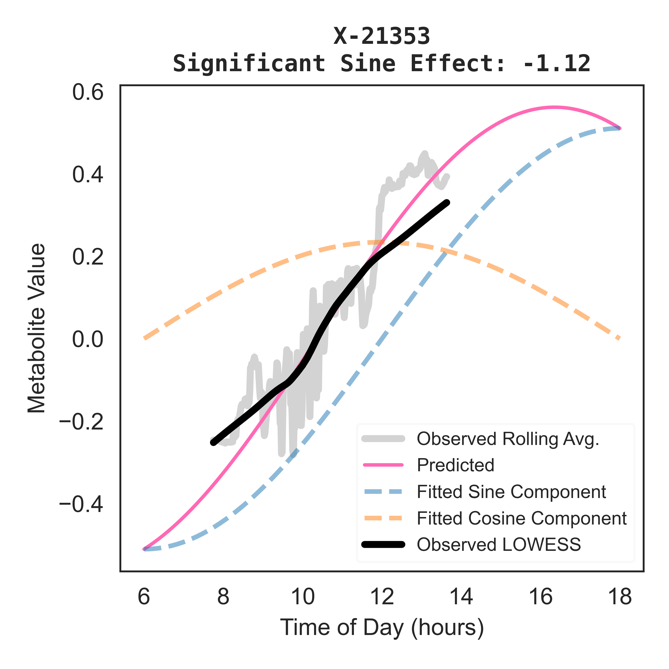

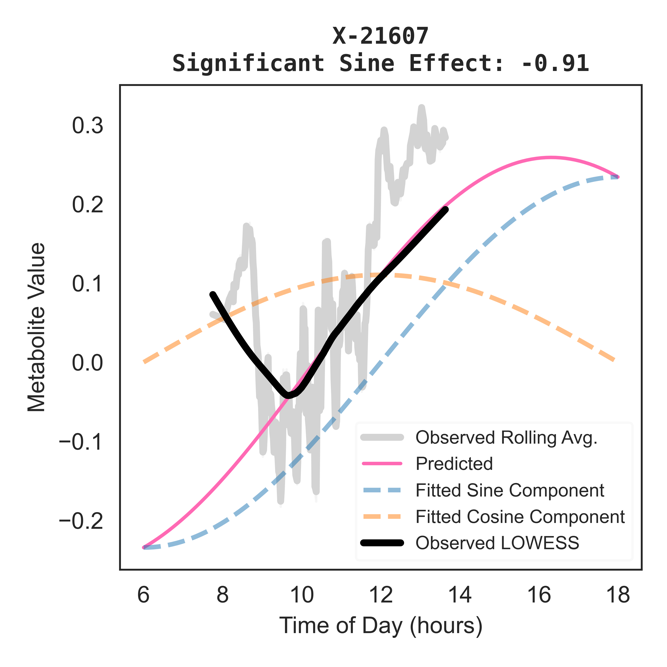

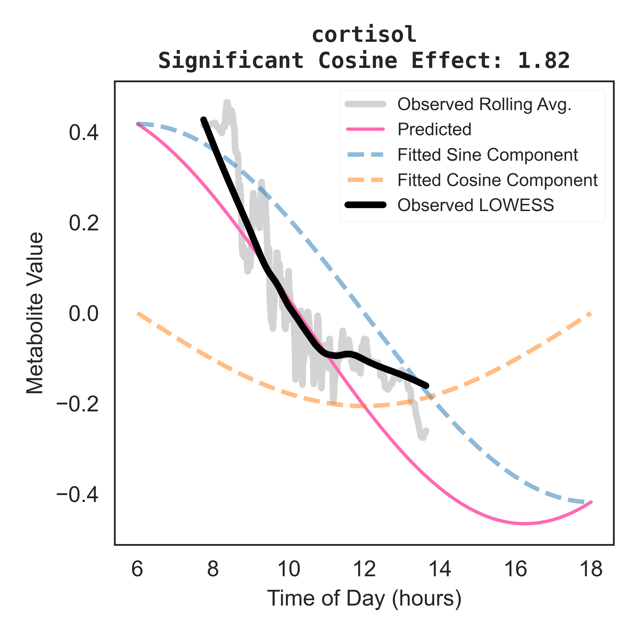

### Supplementary Figure 3. Comparing Batch-Level and Meta-Analysis Level Summary Statistics

These forest plots depict effect size and 95% confidence intervals for all metabolites identified across pooled and stratified primary and sensitivity models to be significantly associated with either sine or cosine time of blood draw (Supplementary Table 9). Black error bars and diamond-shape effect estimates are results from the final meta-analysis across the two HCHS/SOL batches, and colored (according to the legend) circle-shape effect estimates, are results for a specific batch (Batch 1, or Batch 2).

A. **Significant Sine Effect Estimates**

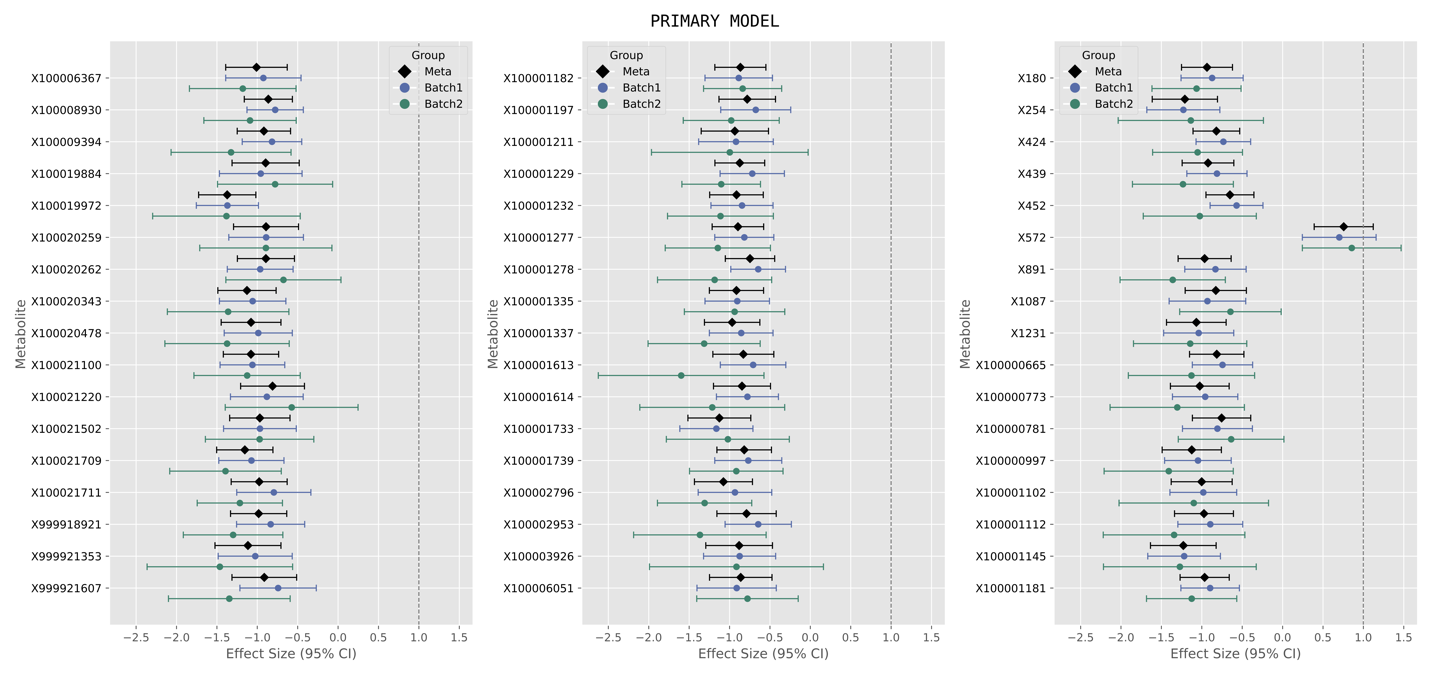

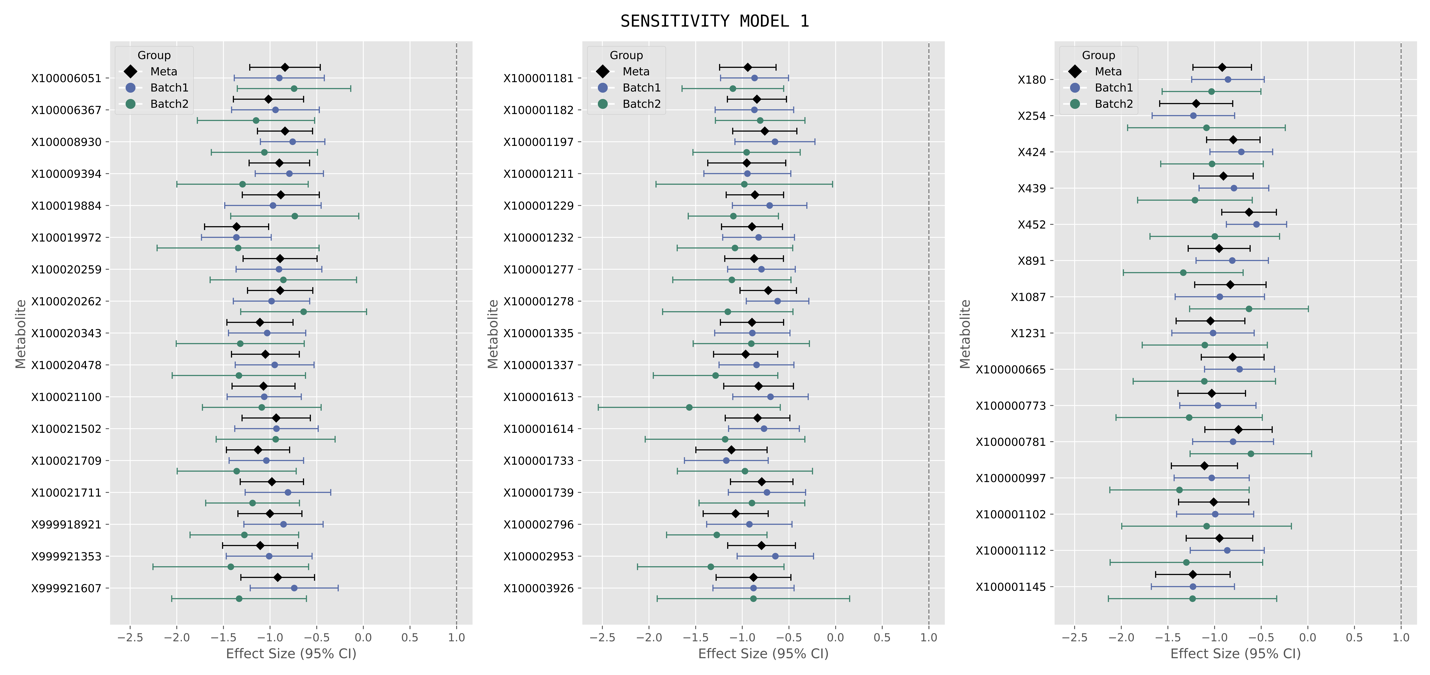

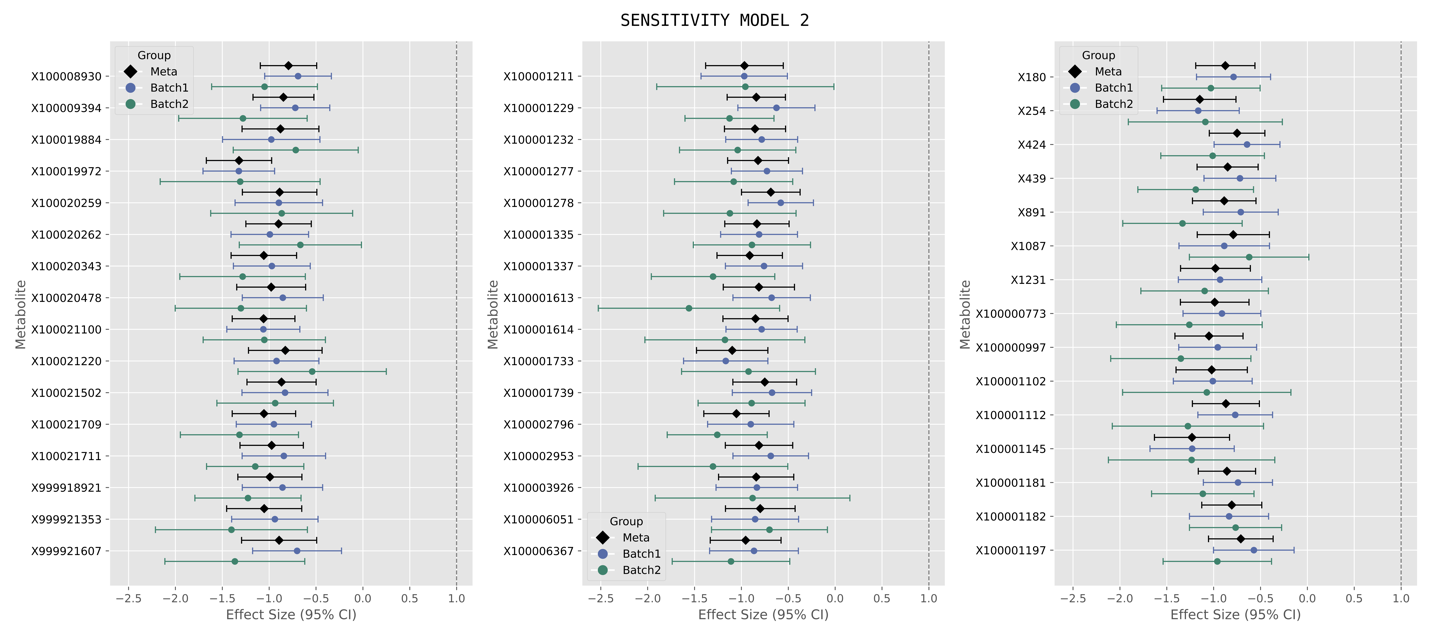

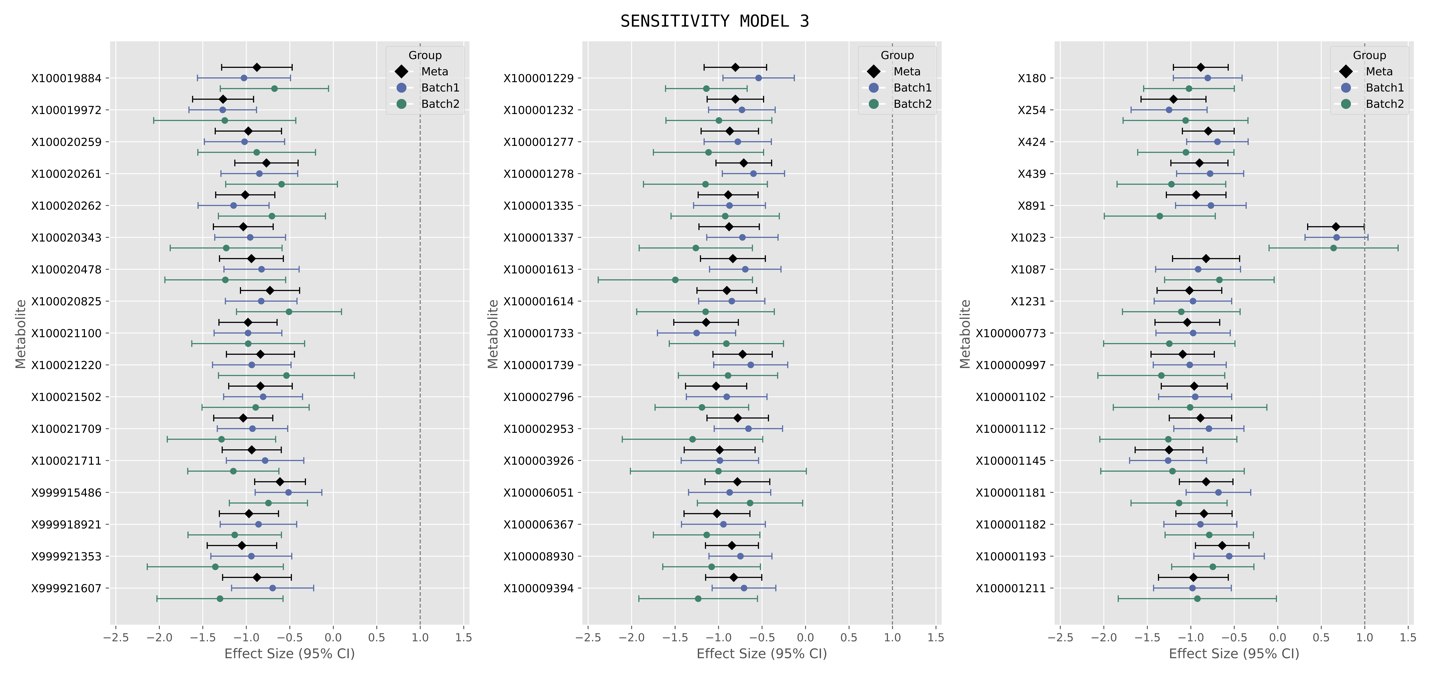

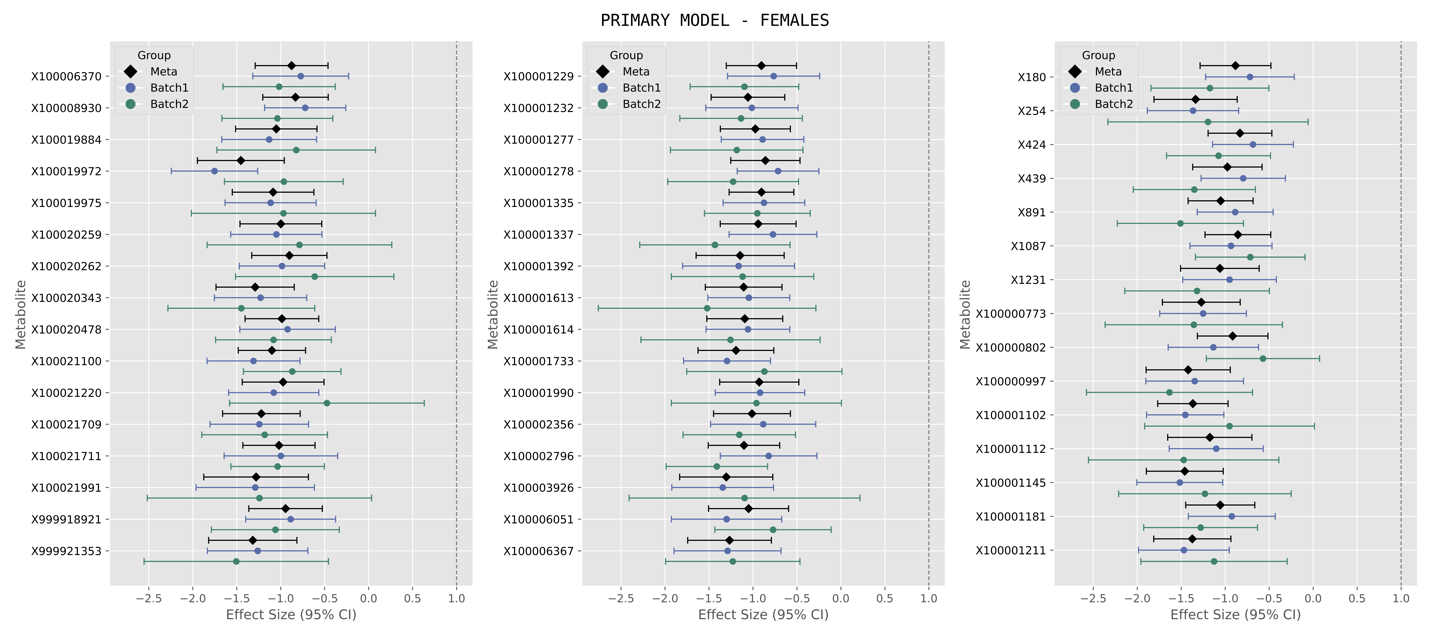

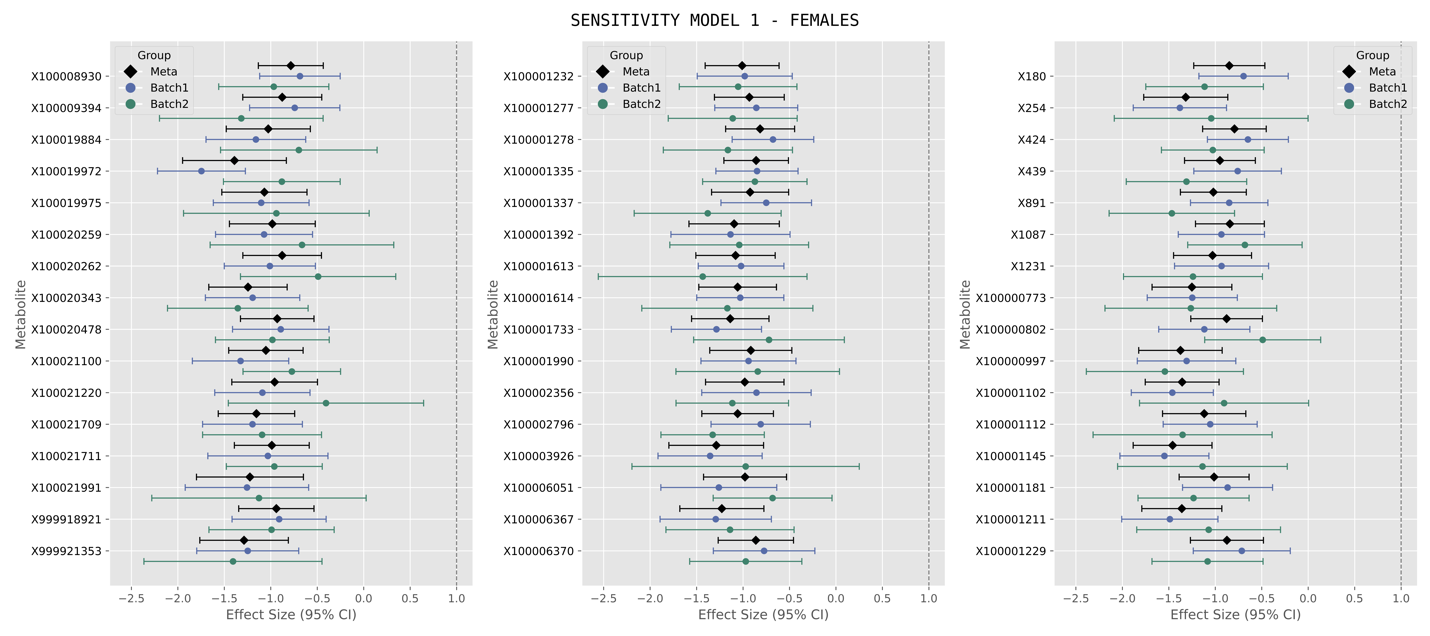

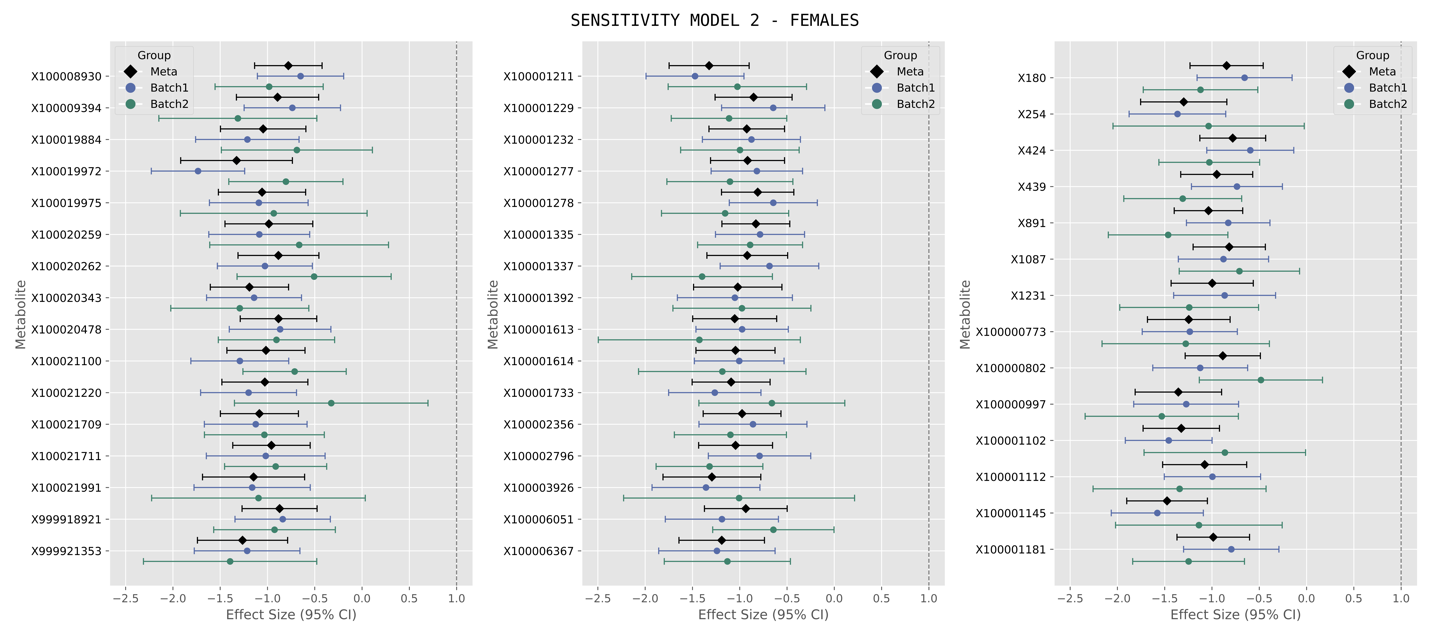

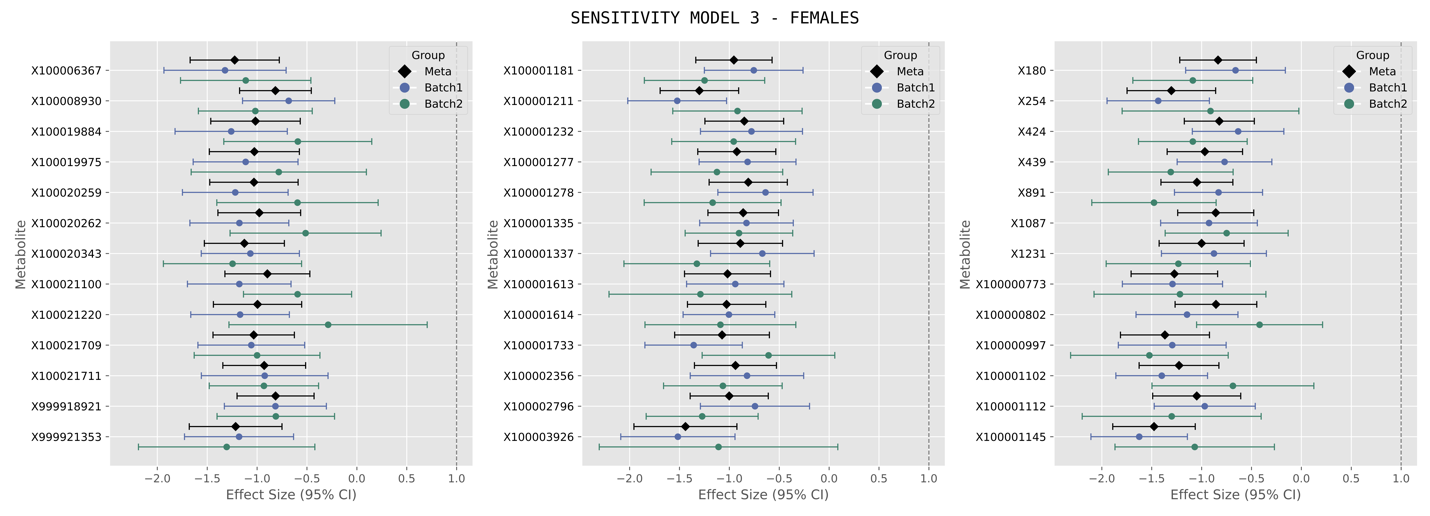

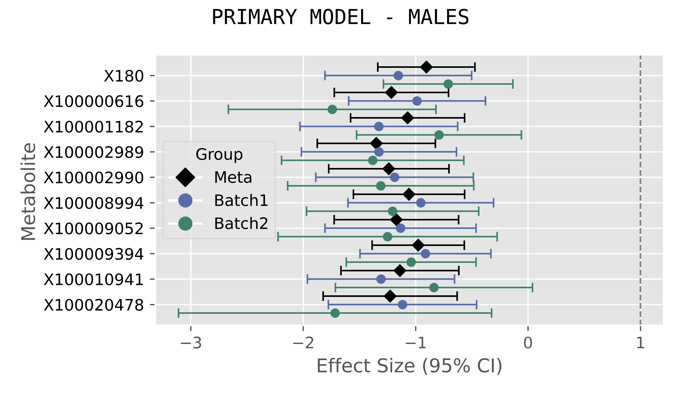

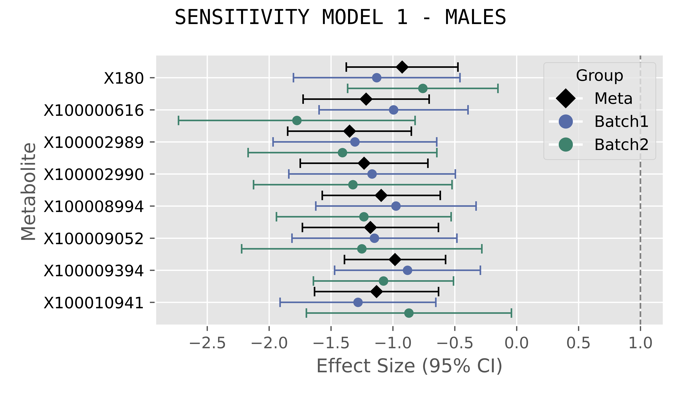

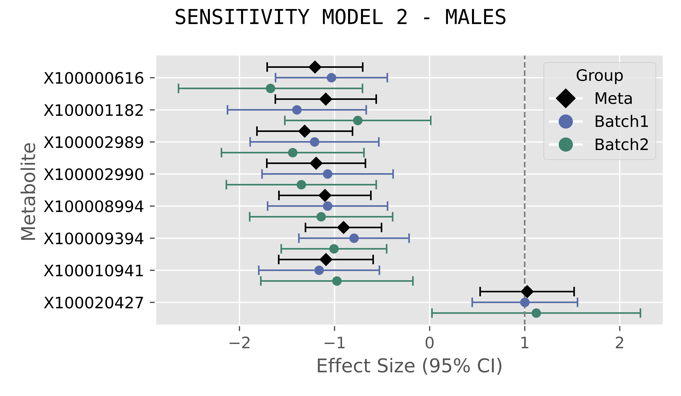

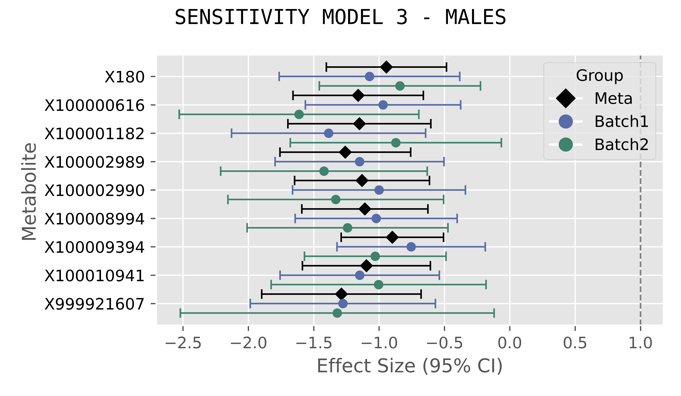

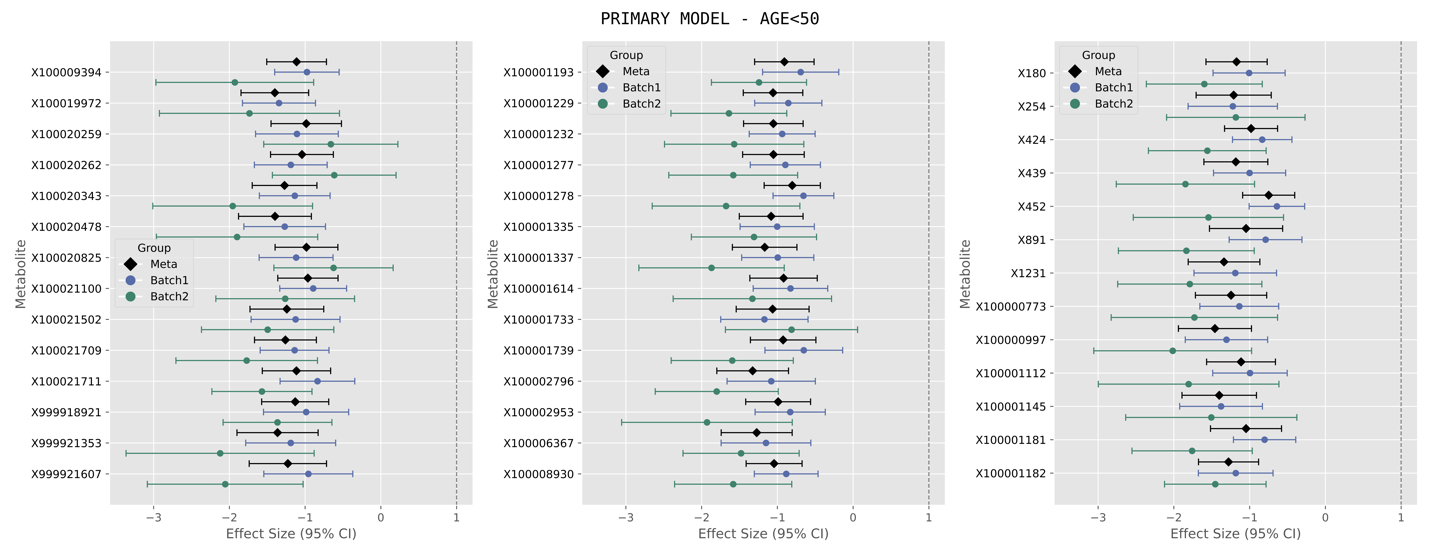

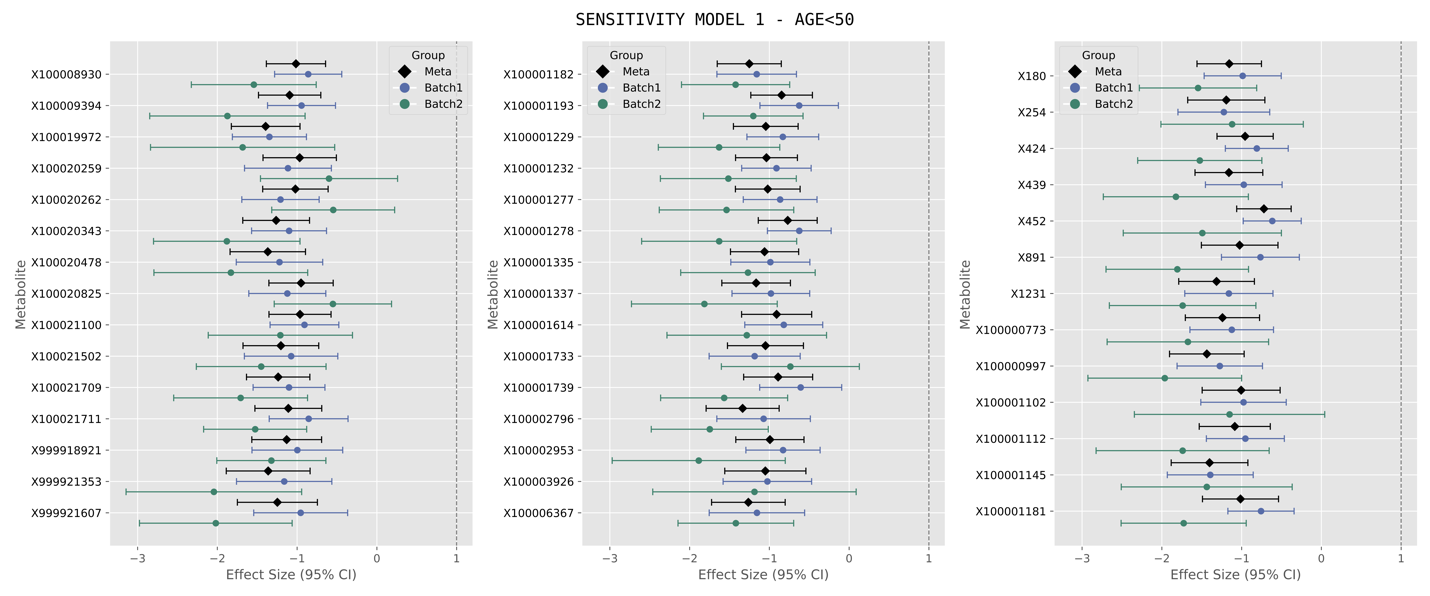

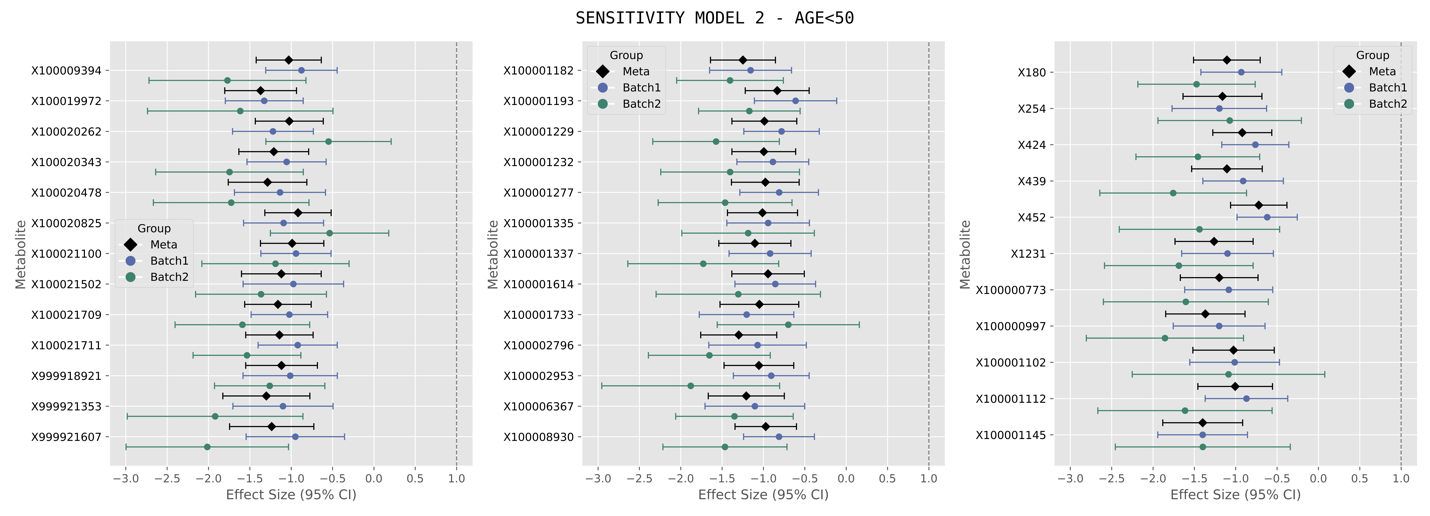

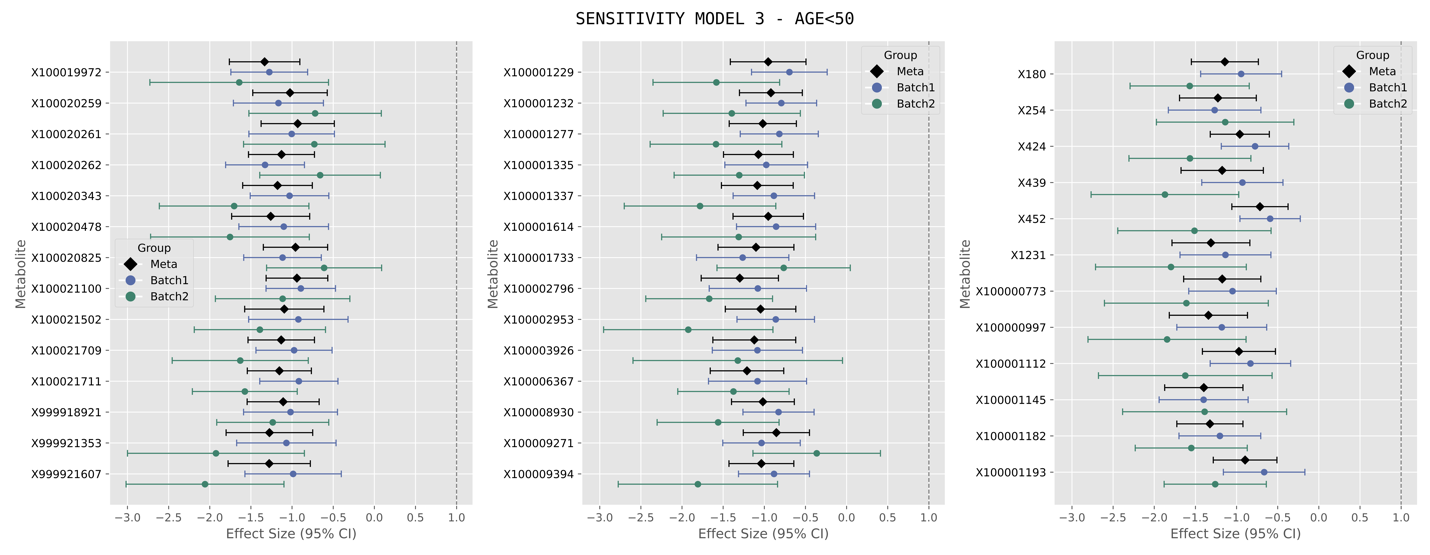

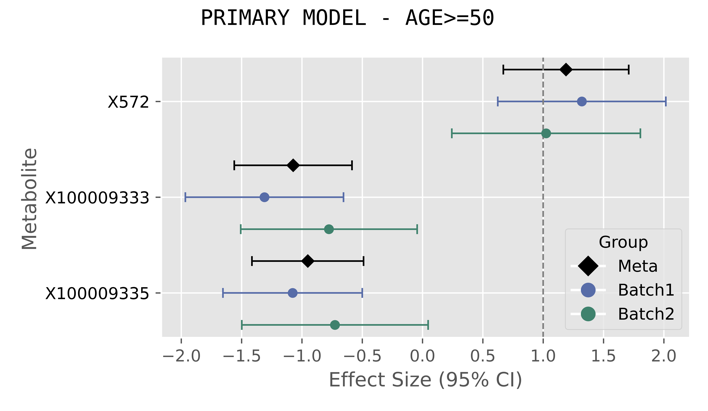

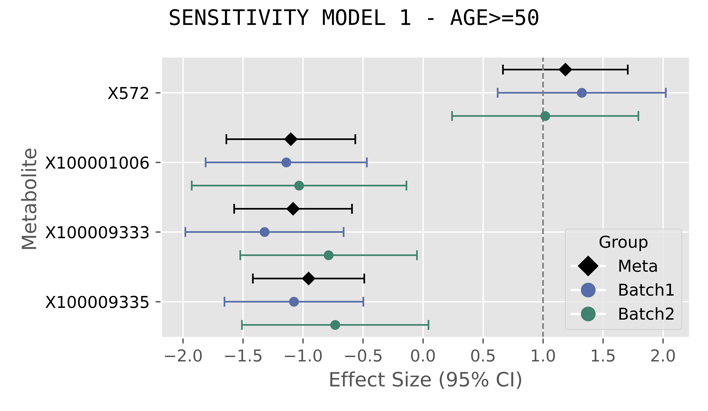

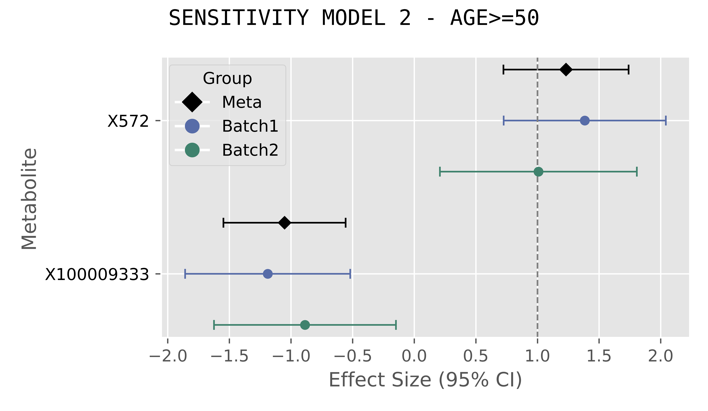

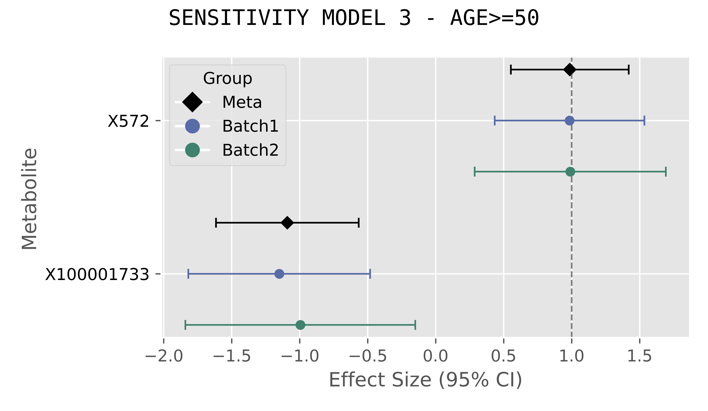

B. **Significant Cosine Effect Estimates**

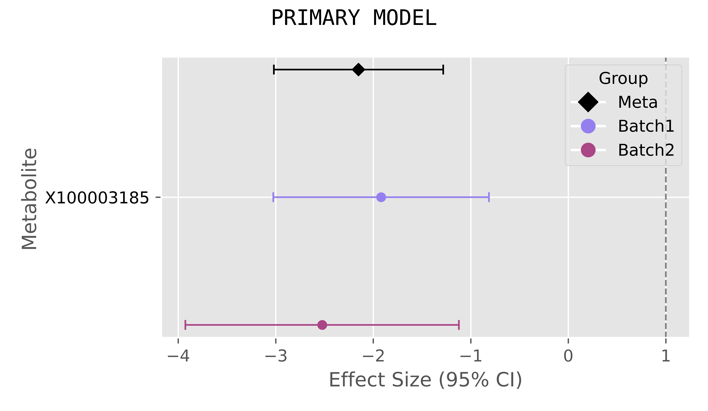

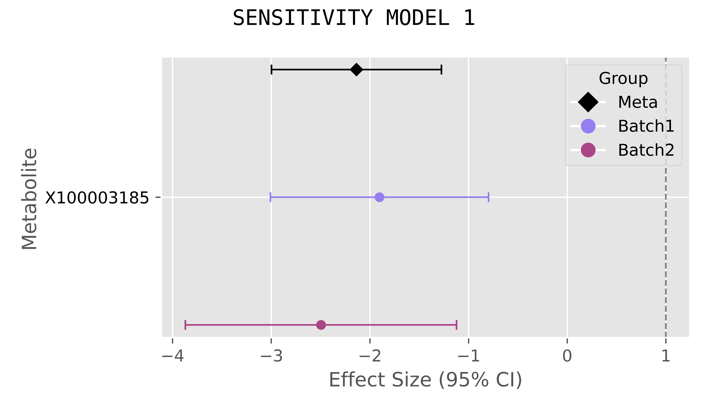

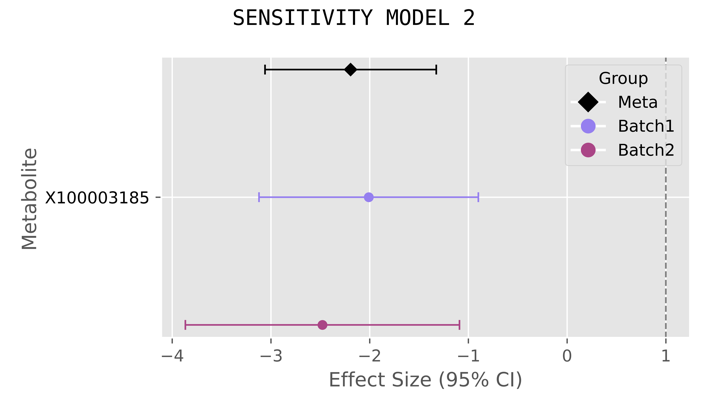

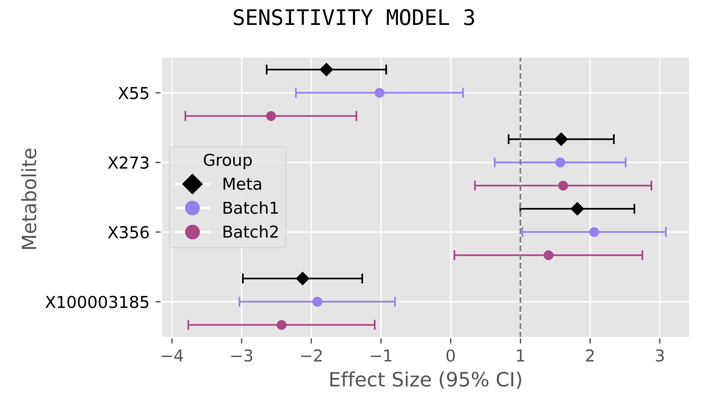

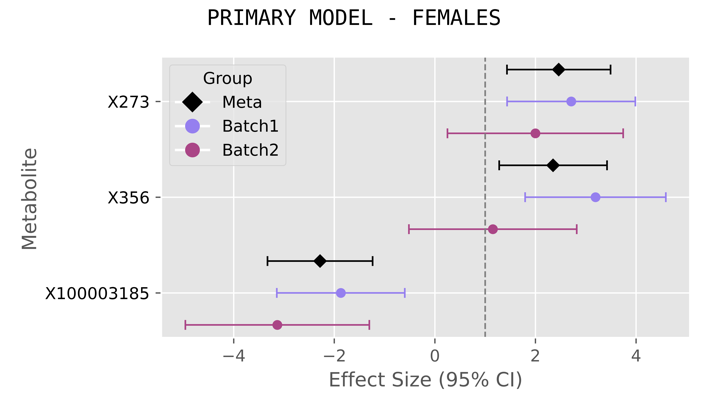

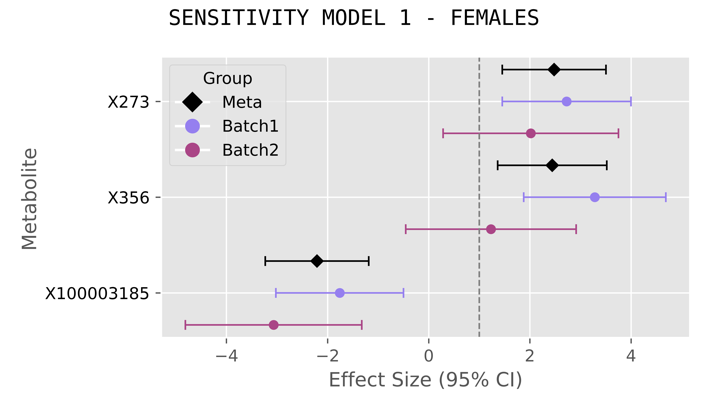

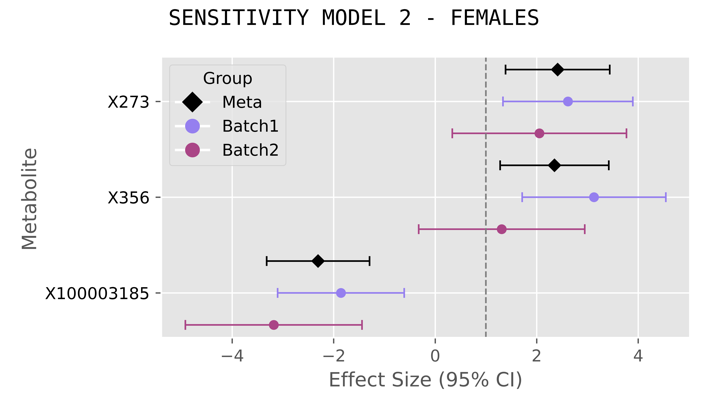

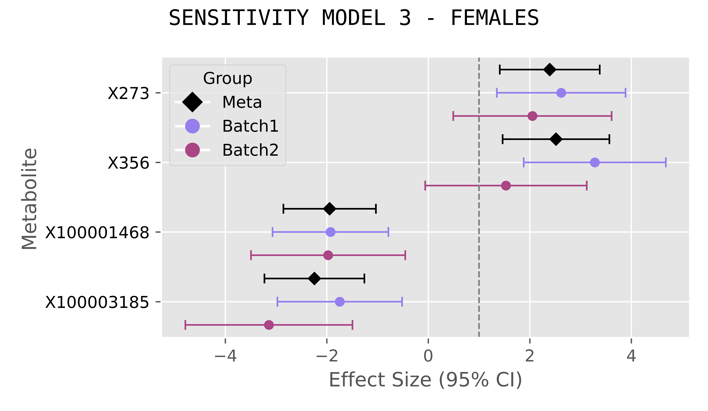

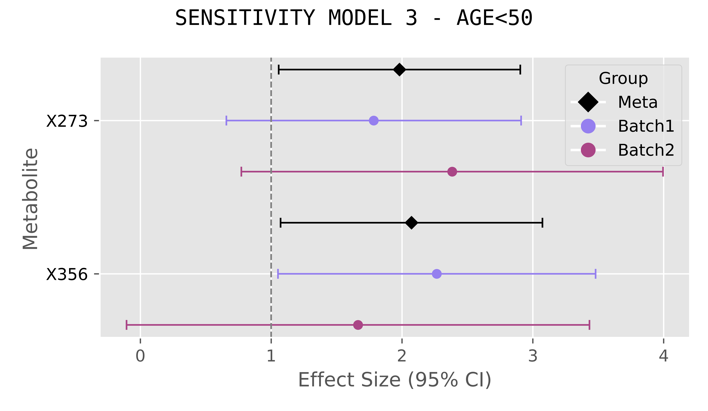

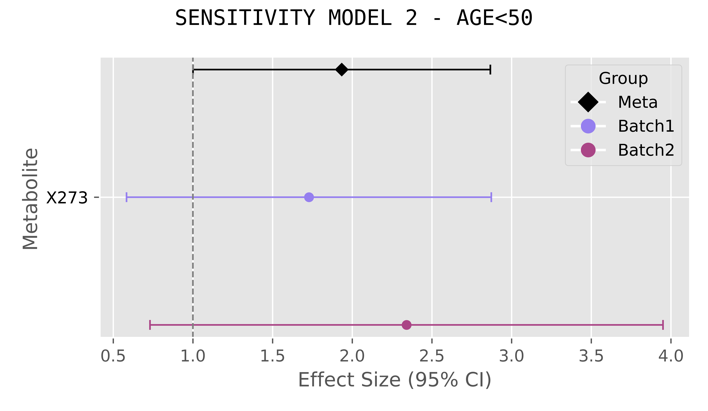

### Supplementary Figure 4. Alternate Clustering Approach to HDBSCAN

K-means clustering was provided as input PCA components that explain up to 80% of variance of the input serum metabolite matrix (853 metabolites x 5954 samples), with # of clusters chosen based on the Elbow Plot method assessing the sum of squared distances to nearest centroid (SSE), and silhouette scores. This resulted in 50 clusters, 175 PCA dimensions, with the achieved silhouette score being 0.12 (very low) suggesting this linear clustering approach was not able to discover separable, underlying structure in the input serum metabolite measures and time of blood draw. In this plot, panel (A) depicts the SSE (y-axis) for k chosen clusters (x-axis) and panel (B) depicts average silhouette scores (y-axis) for k chosen clusters (x-axis).

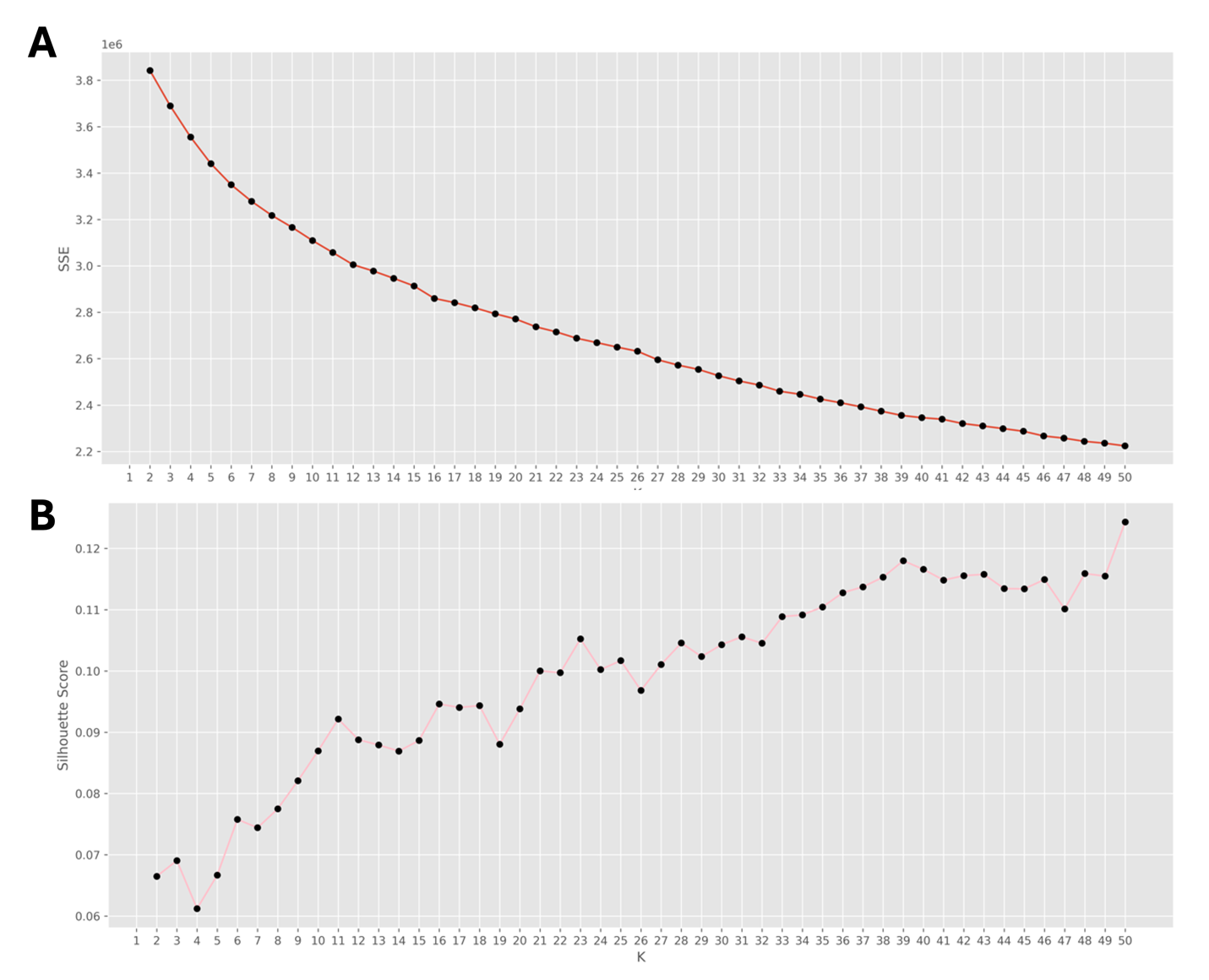

### Supplementary Figure 5. Serum Metabolite Cluster Characteristics

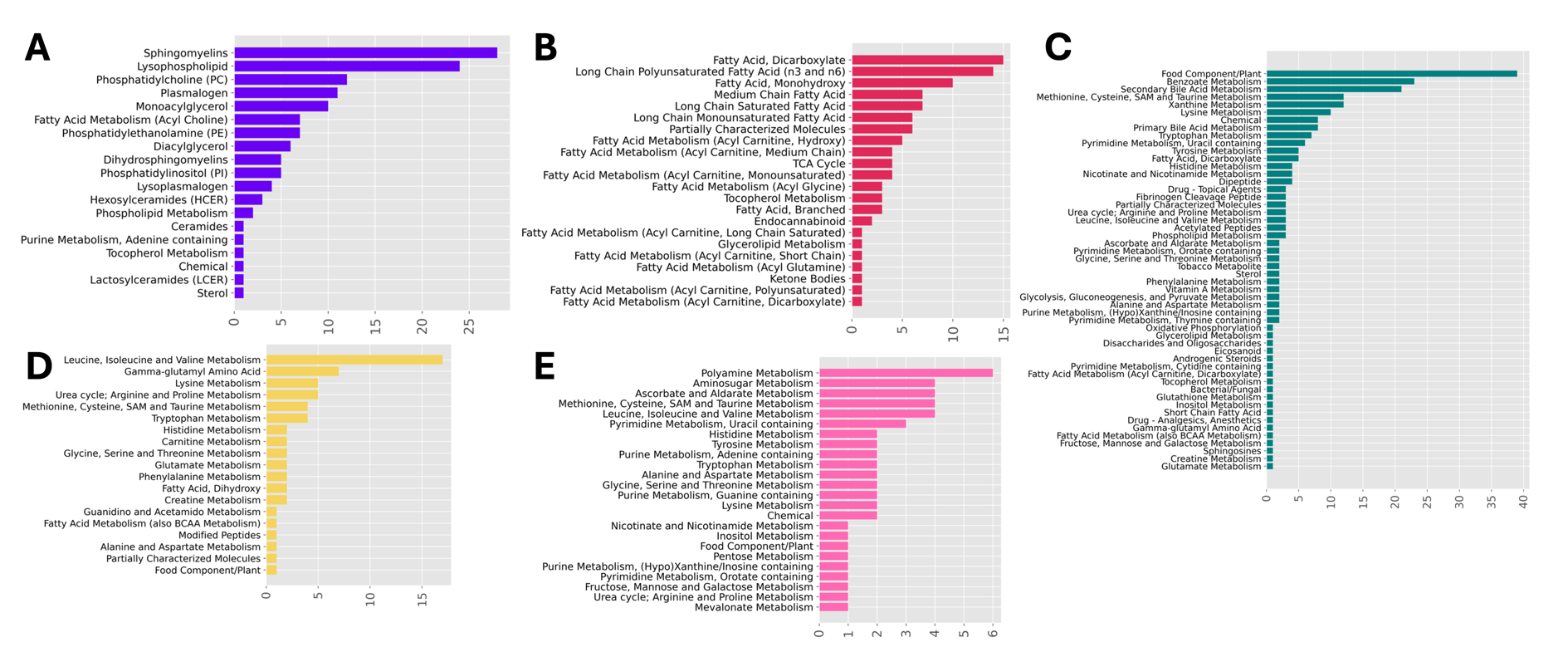
Depicted here are horizontal bar graphs illustrating the differing types of serum metabolites harbored by each of the five HDBSCAN metabolite clusters (A. Cluster 1, B. Cluster 2, C. Cluster 3, D. Cluster 4, E. Cluster 5). On the y-axis is the name of the metabolite category, and the x-axis the count (number of metabolites).

### Supplementary Figure 6. Summary of 673 Metabolite-Health Trait/Disorder Associations

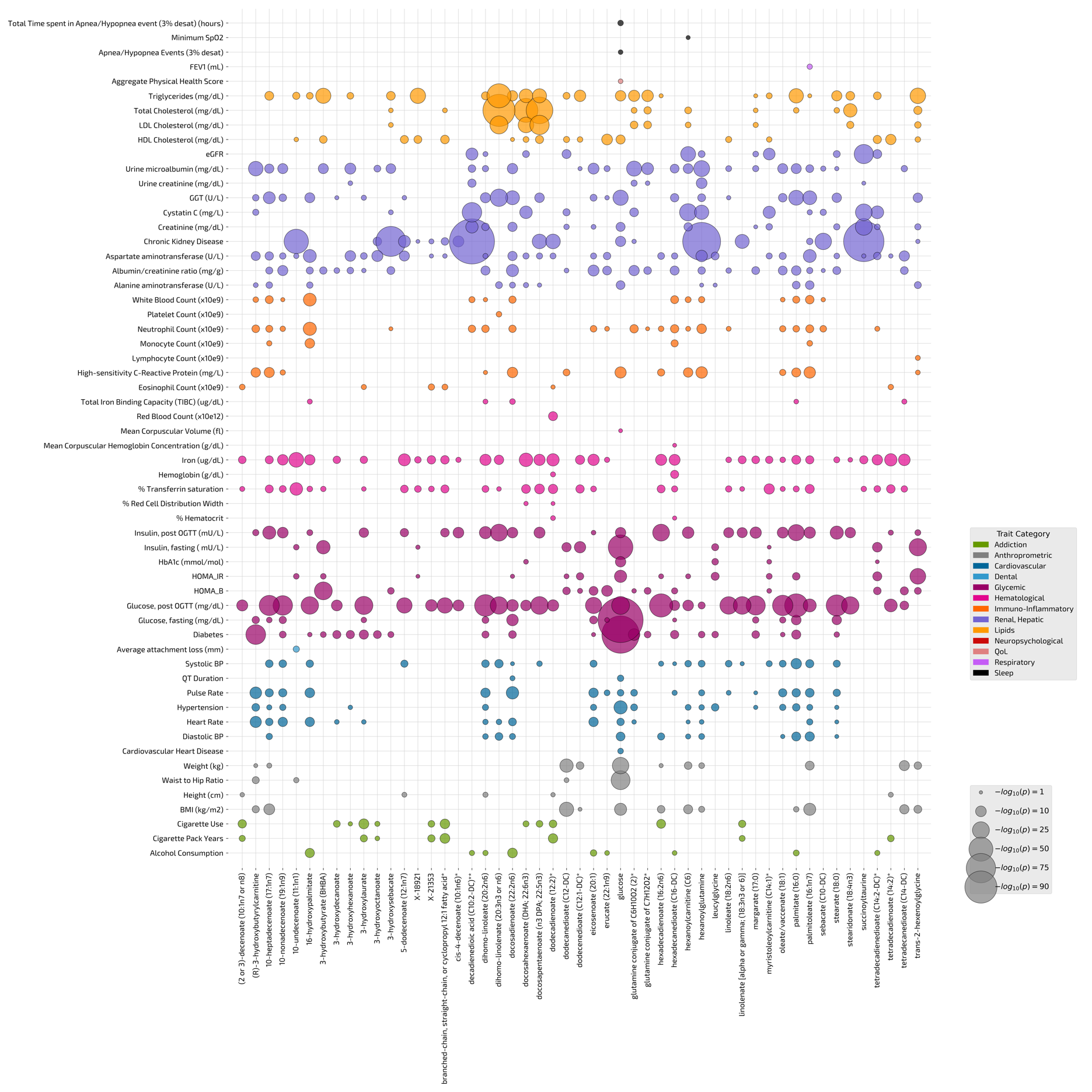
This figure depicts a bubble plot displaying all 673 metabolite-trait associations revealed by 51 fasting serum metabolites that exhibit association with time of blood draw, across 58 health traits/disorders (Supplementary Tables 18-19). Each row is a given health trait/disorder, and each column a particular serum metabolite. The colors of each scatter point are according to specific health trait categories, and the size according to strength of statistical association (-log10 transformed p-value). The health category colors, and scale of scatter point size with respect to p-value strength are specified in the right-hand side legend boxes.

### Supplementary Figure 7. Serum Metabolite Associations across 91 Health Traits

Displayed here are results from the metabolite-trait association analysis, for each of the 52 time of blood draw associated serum metabolites, across 91 health traits (Supplementary Tables 18-19). An upward triangle in each plot denotes positive effect direction, whereas an upside-down triangle a negative effect direction. Results with p<1.06e-05 (accounting for multiple tests across 52 metabolites and 91 traits) are considered statistically significant, and labeled by their names in the plot. The red horizontal line represents the significance threshold of 1.06e-05.

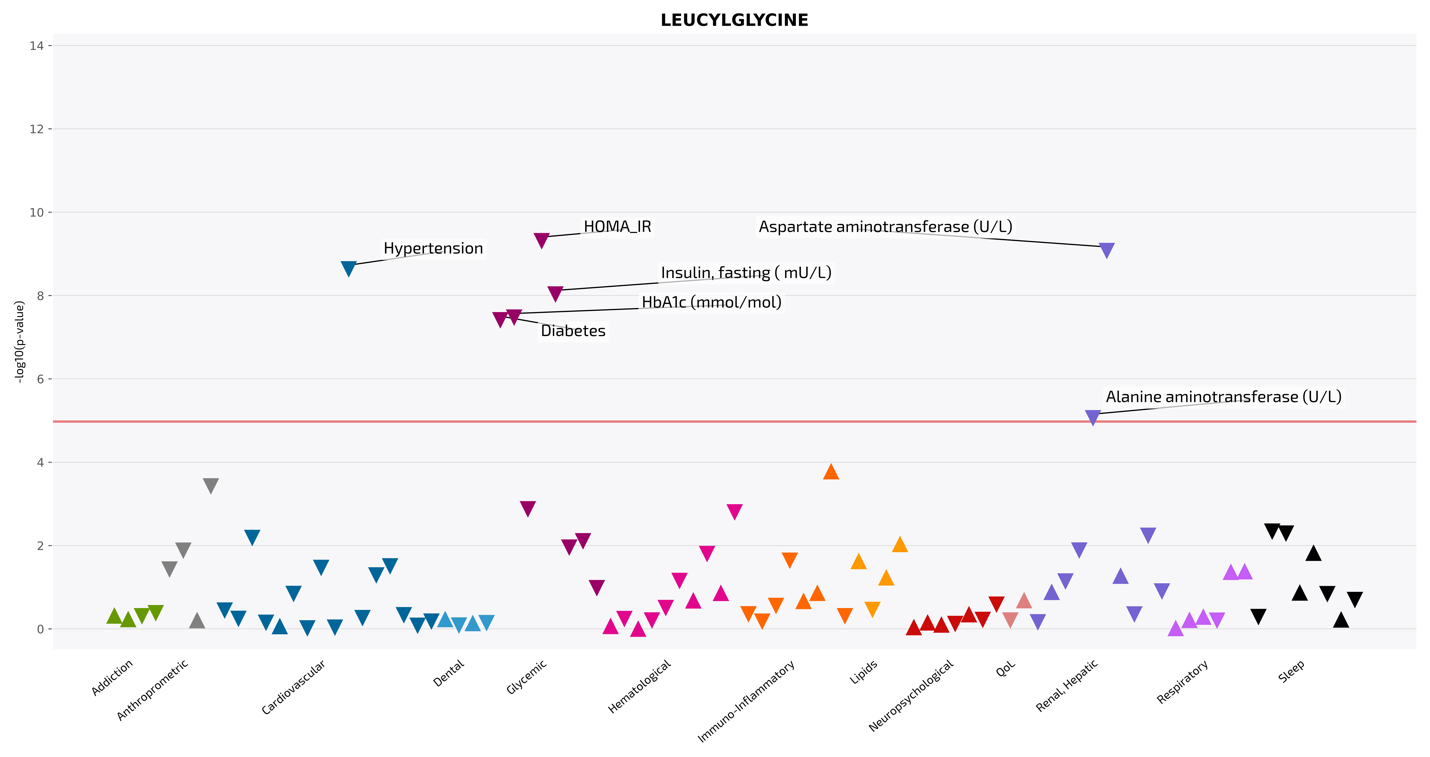

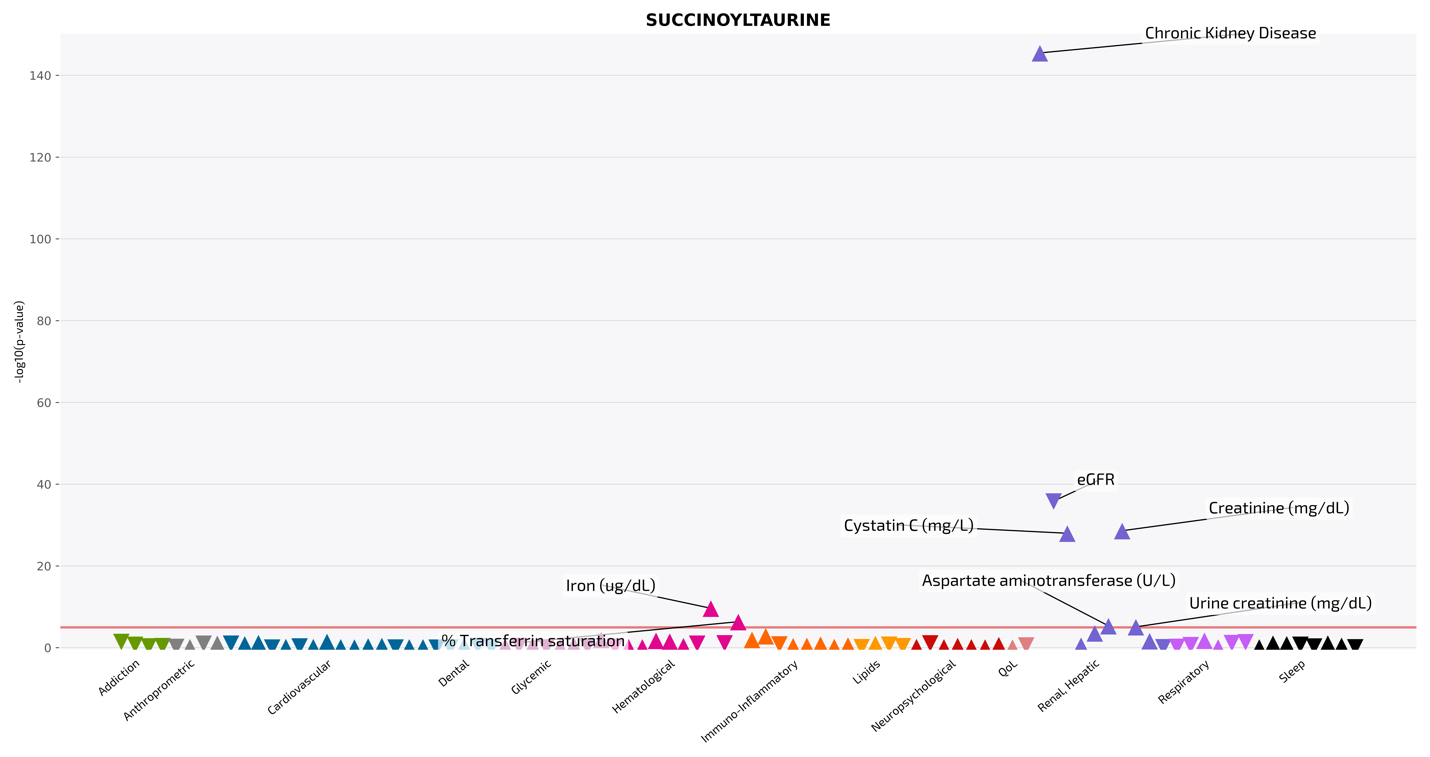

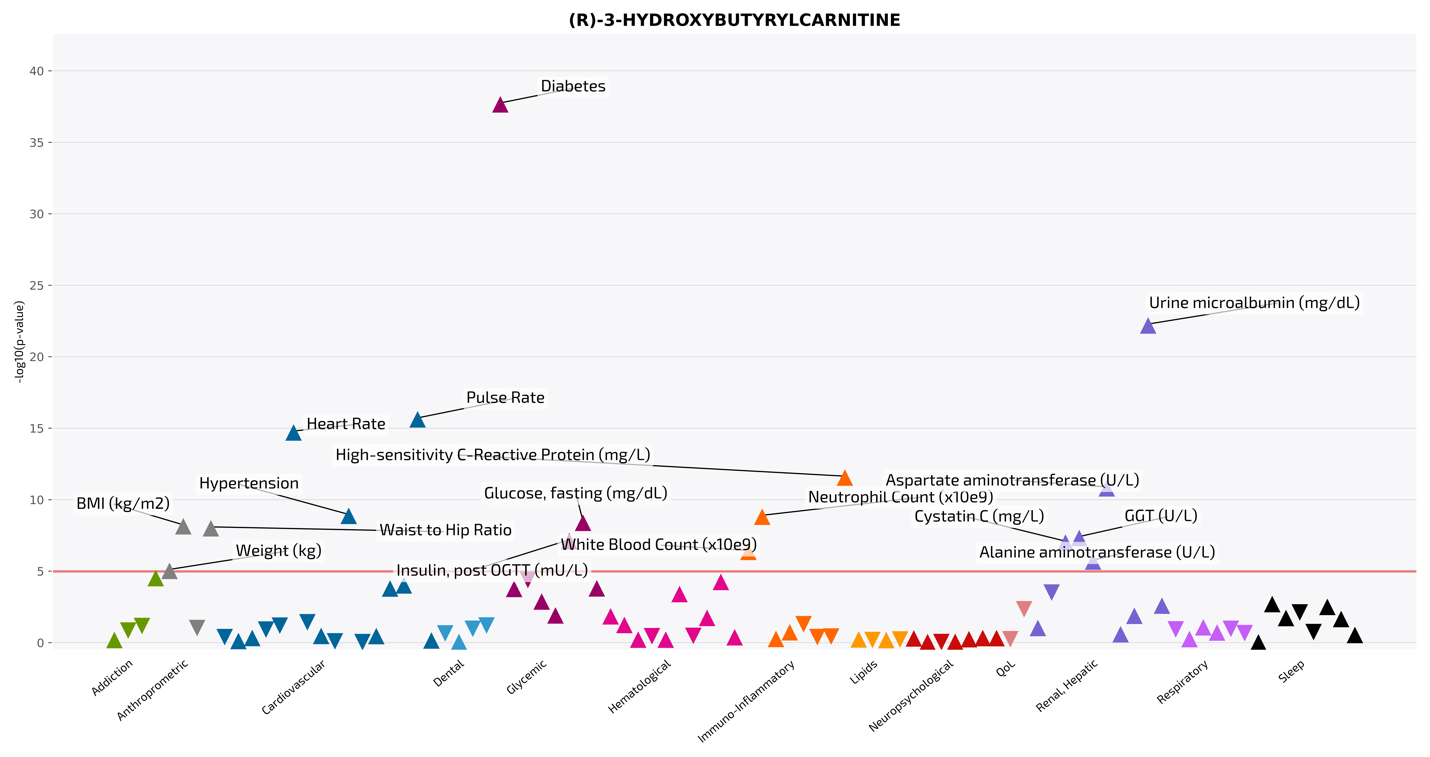

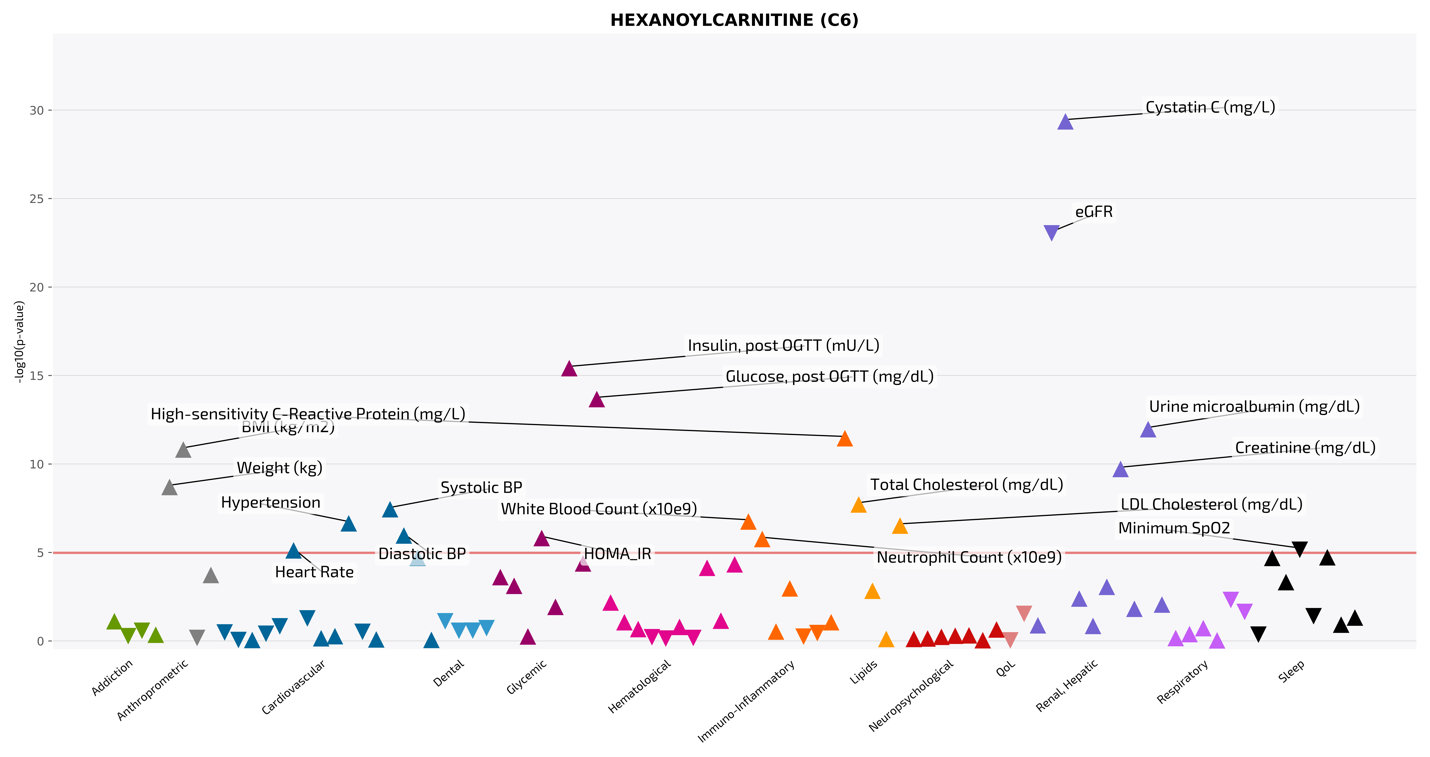

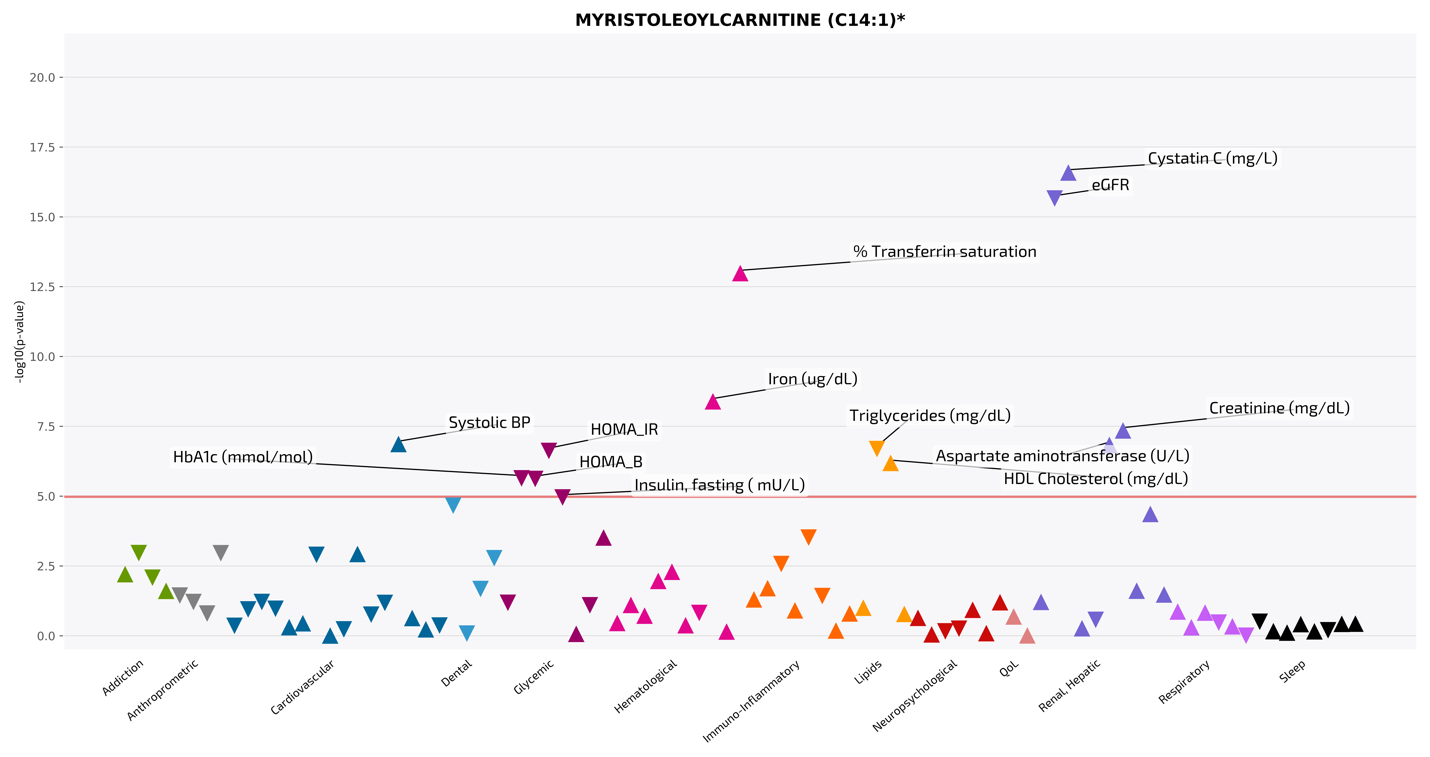

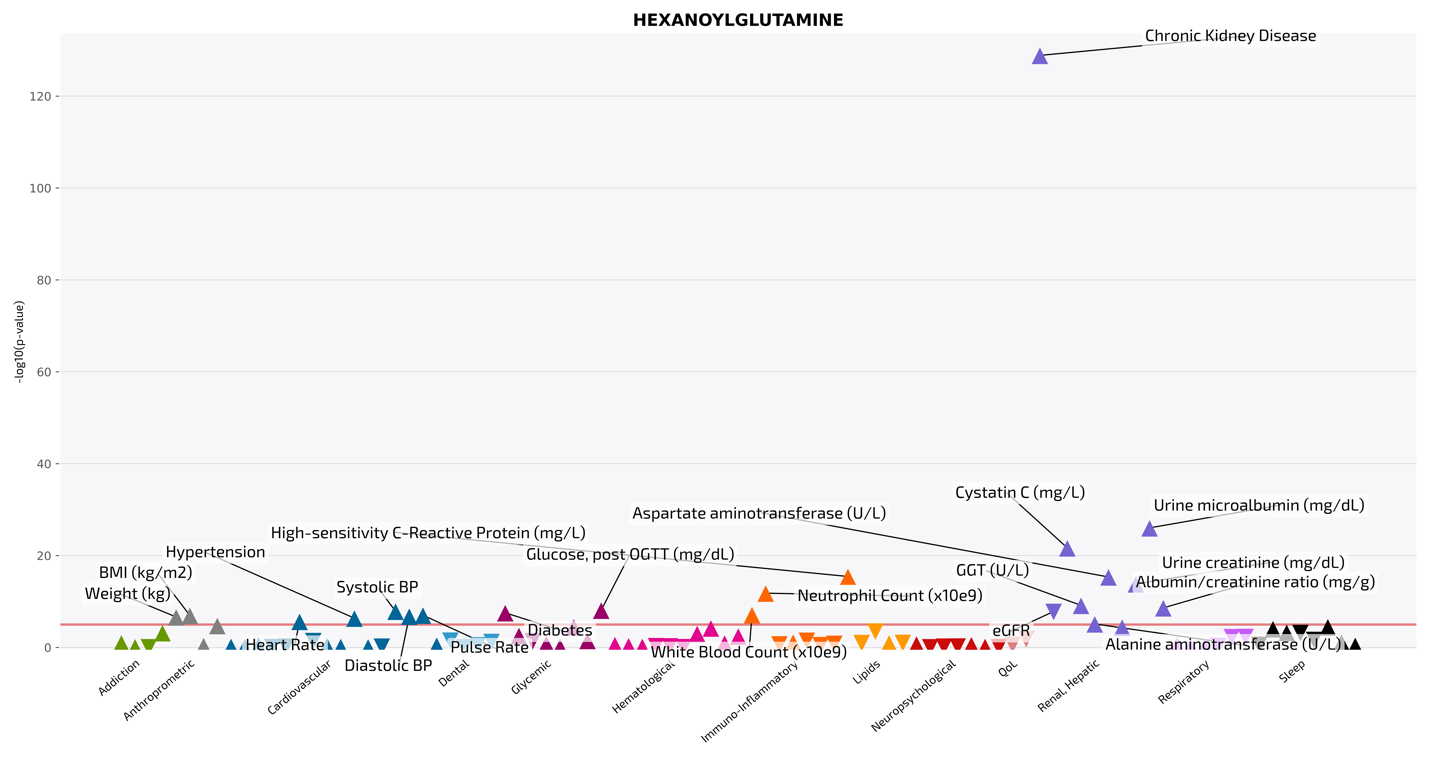

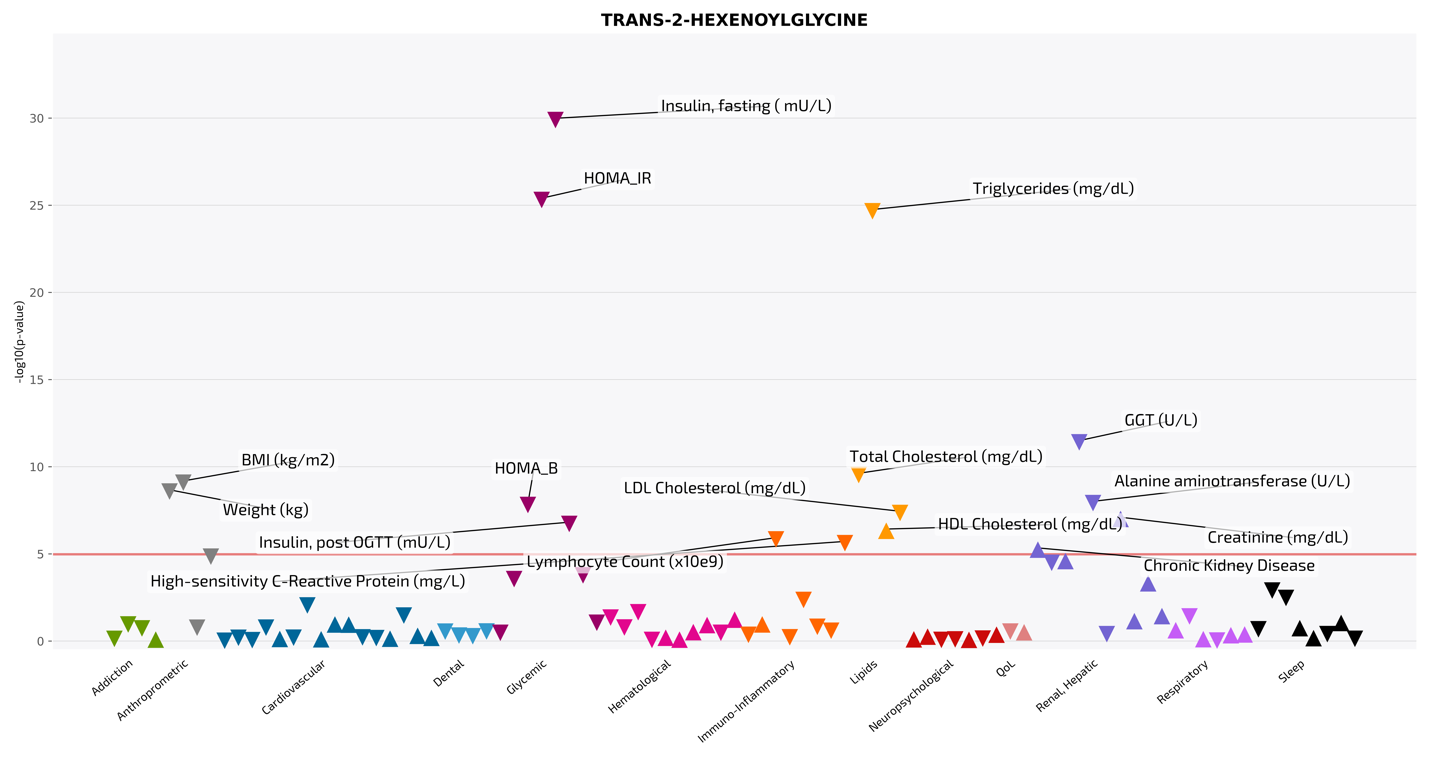

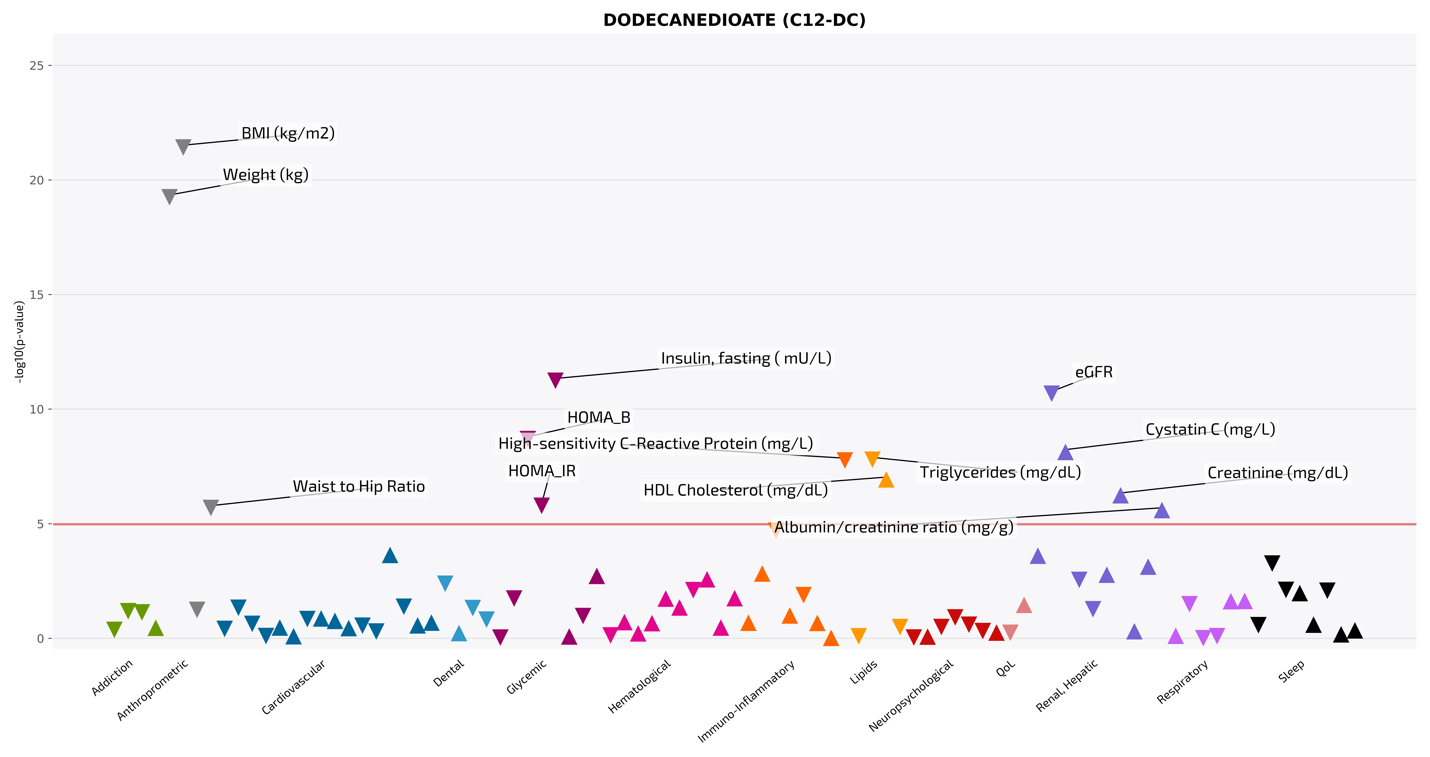

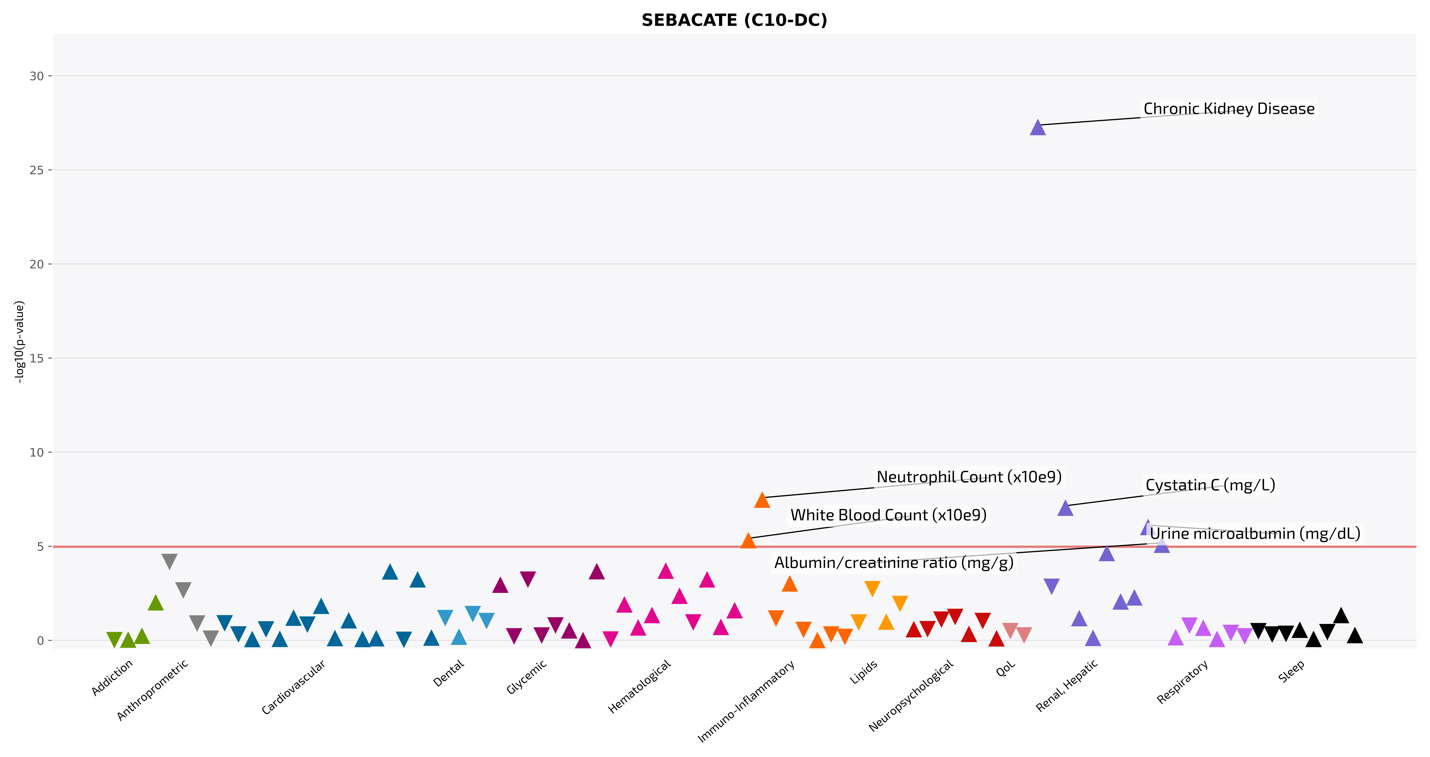

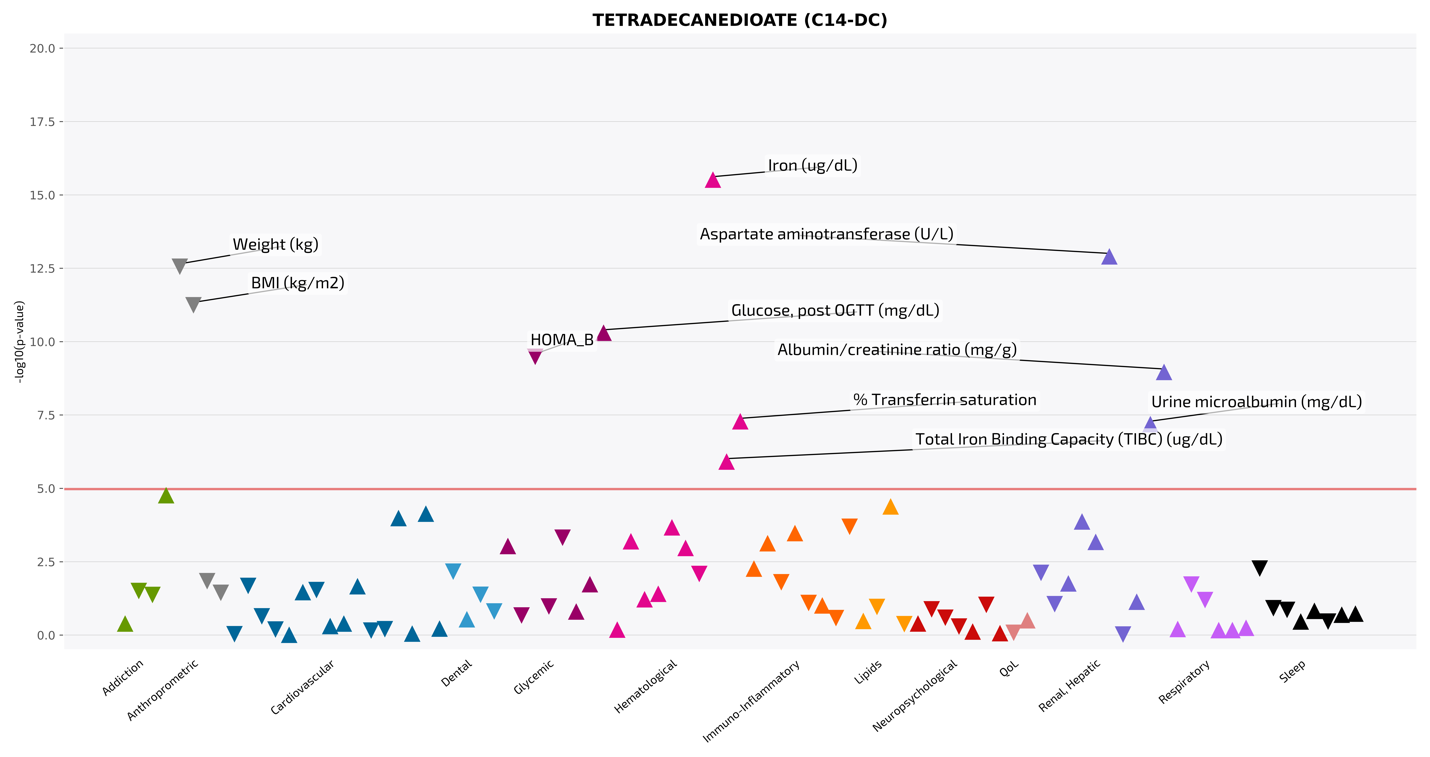

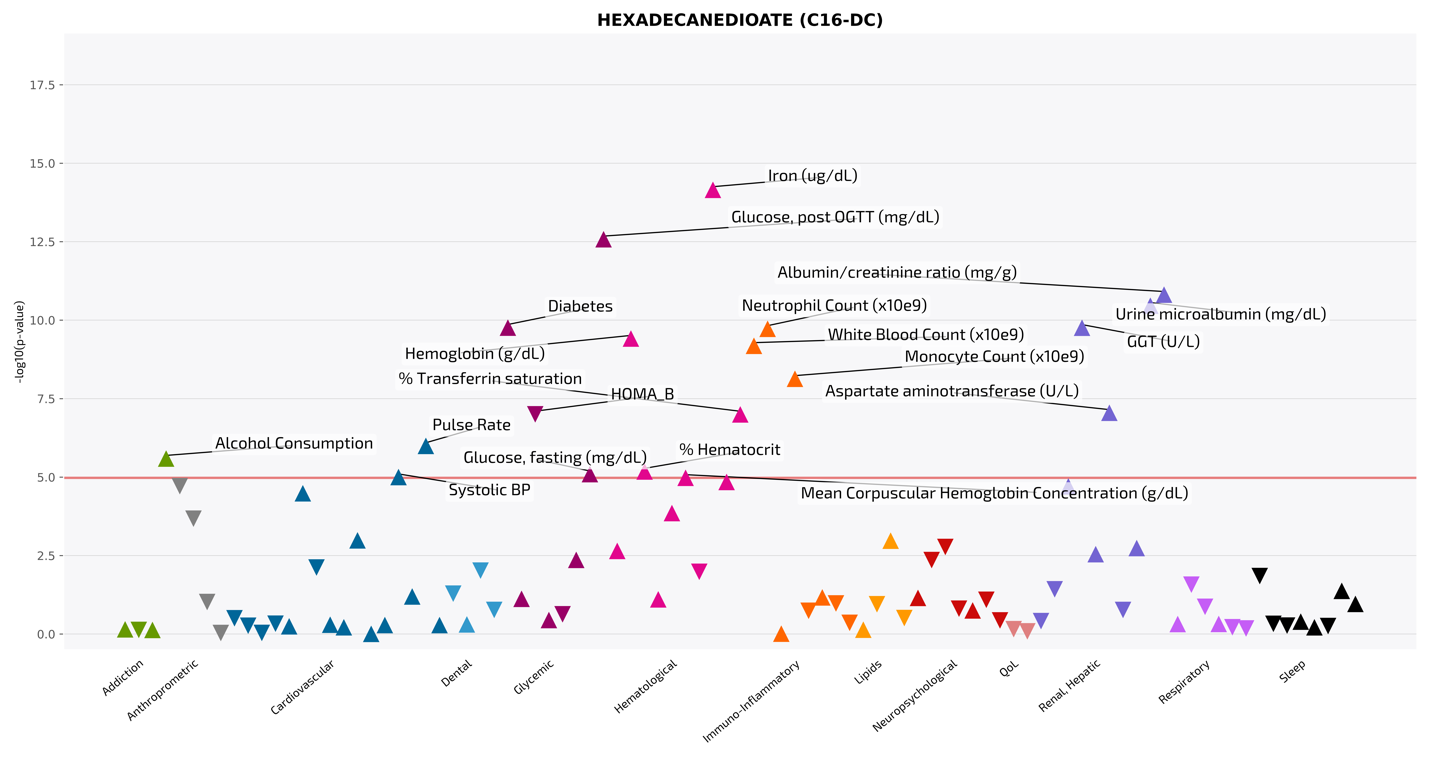

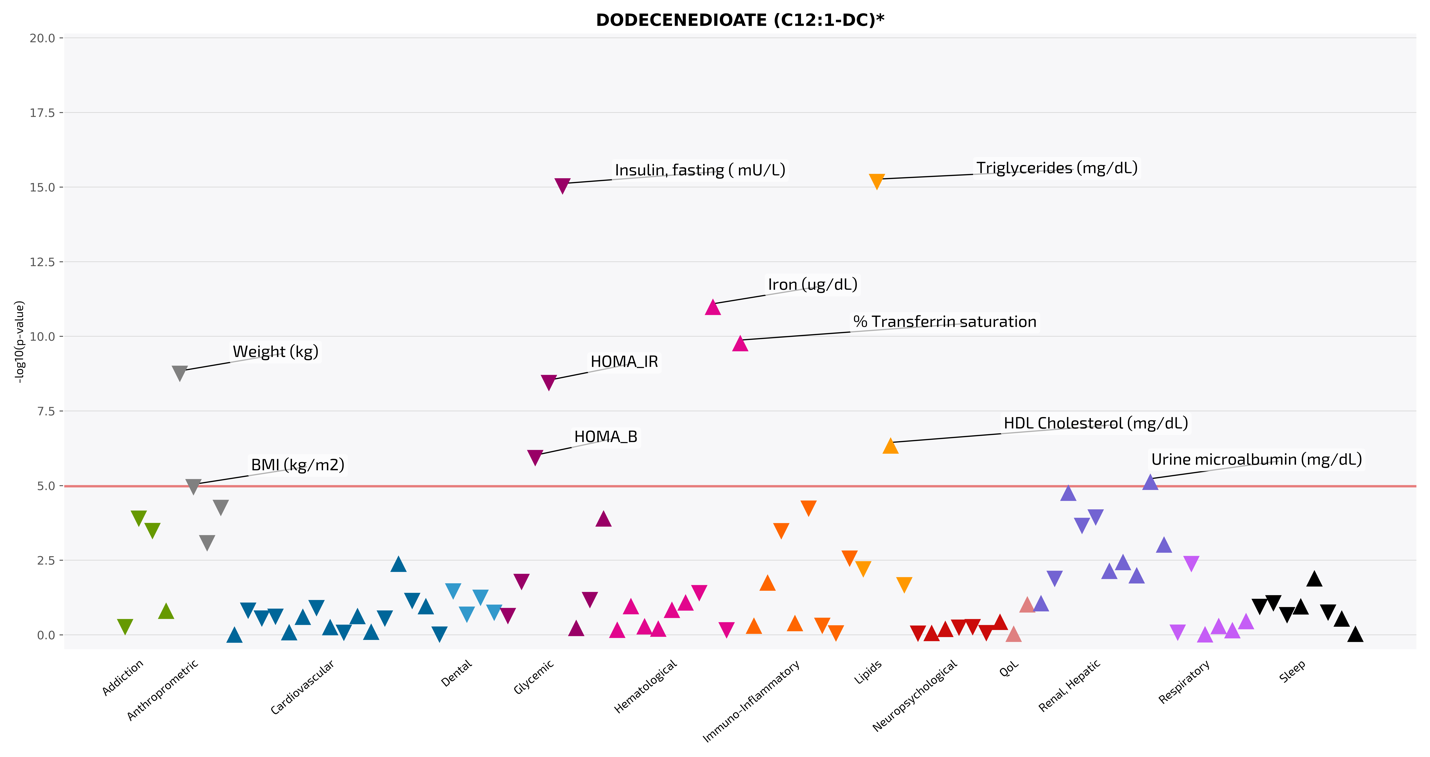

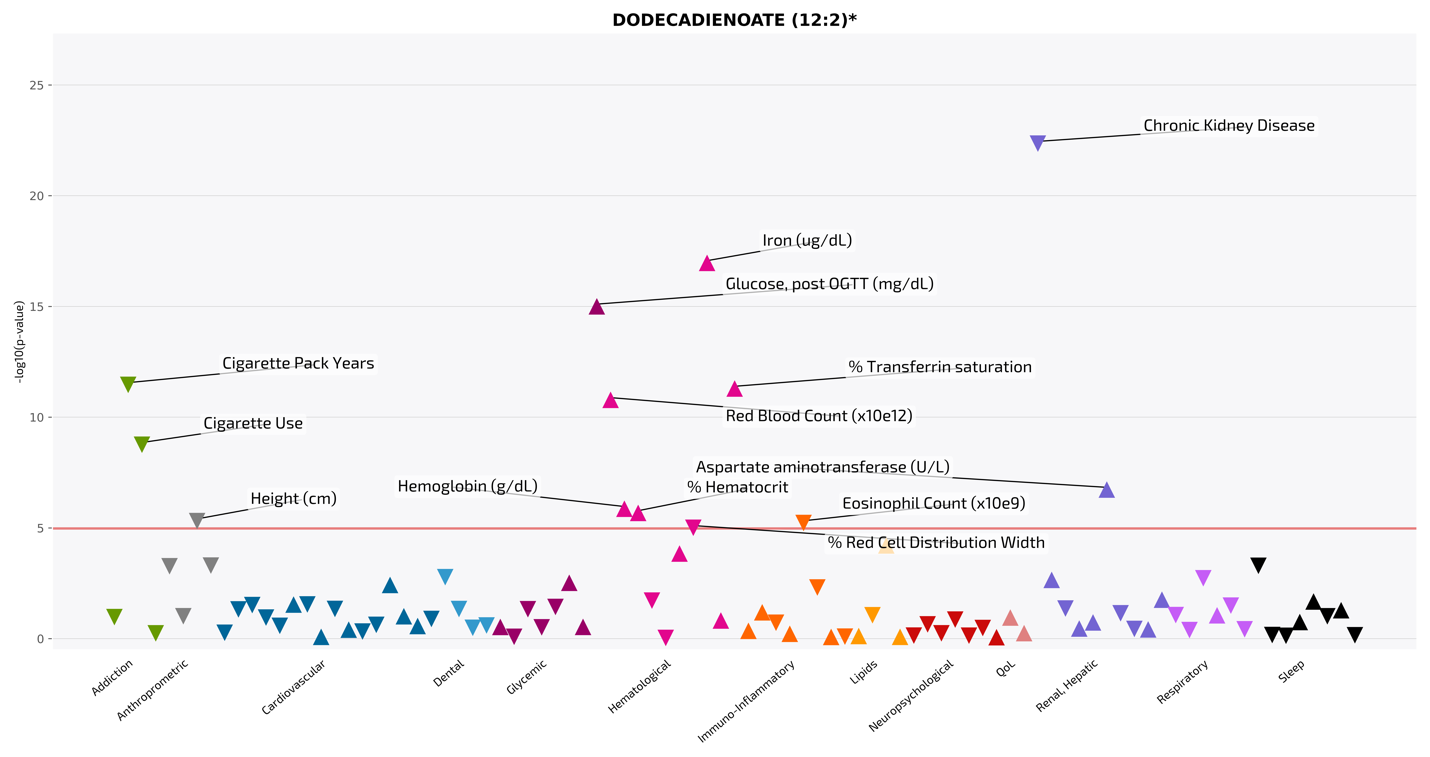

### Supplementary Figure 8. Comparing Models for Metabolite Associations with Health Traits

In panel (A), p-values from time of blood draw adjusted models (x-axis) are compared to p-values from models with no time of blood draw adjustment (y-axis) in a scatter plot. Depicted in panel (B) are effect size ratios, and in panel (C), difference in -log10 p-values, comparing models adjusting for blood draw time, and models not adjusting for blood draw time for the 673 metabolite-trait associations reported in Supplementary Tables 18-19.

### Supplementary Figure 9. Impact of Time of Blood Draw Adjustment on Metabolite-Trait Discovery

This forest plot displays 57 metabolite-trait associations (effect sizes, 95% confidence intervals) that exhibit >10-fold p-value difference and altered statistical significance with a given health trait/disorder, depending on model adjustment for time of blood draw. Black color depicts unadjusted models. Bright colors depict time of blood draw-adjusted models, where the color of the error bar and effect size scatter point are according to the health domain legend in the bottom center of the plot. Summary statistics associated with these 57 metabolite-trait associations are provided in Supplementary Table 23.

### Supplementary Figure 10. Comparing Models for Metabolite Associations with Health Traits

These forest plots compare effect size, 95% confidence intervals, and p-values for the 973 significant metabolite-trait associations identified across 51 time of blood draw associated serum metabolites from models adjusting for blood draw time, to models not adjusting for blood draw time (Supplementary Tables 18-19). Gray error bars and scatter are results from models not adjusting for blood draw time, and in color (according to trait category) models adjusting for blood draw time. P-values are provided on the left axis.

### Supplementary Figure 11. Single metabolite-outcome associations for a simulated circadian metabolite.

Panel (A) depicts bias (defined as the difference between the estimated and true β) in the naïve model from a single simulation, changing with metabolite circadian amplitude and the phase difference between metabolite and outcome rhythms. Panel (B) illustrates the distribution of estimated effects (β) from 1,000 simulations for three outcome scenarios: constant, in-phase with metabolite, and π/2 phase-shifted. Panel (C) illustrates the distribution of p-values across the same simulations and scenarios.
